# Genetic analyses of the QT interval and its components in over 250K individuals identifies new loci and pathways affecting ventricular depolarization and repolarization

**DOI:** 10.1101/2021.11.04.21265866

**Authors:** William J. Young, Najim Lahrouchi, Aaron Isaacs, ThuyVy Duong, Luisa Foco, Farah Ahmed, Jennifer A. Brody, Reem Salman, Raymond Noordam, Jan-Walter Benjamins, Jeffrey Haessler, Leo-Pekka Lyytikäinen, Linda Repetto, Maria Pina Concas, Marten E. van den Berg, Stefan Weiss, Antoine R. Baldassari, Traci M. Bartz, James P. Cook, Daniel S. Evans, Rebecca Freudling, Oliver Hines, Jonas L. Isaksen, Honghuang Lin, Hao Mei, Arden Moscati, Martina Müller-Nurasyid, Casia Nursyifa, Yong Qian, Anne Richmond, Carolina Roselli, Kathleen A. Ryan, Eduardo Tarazona-Santos, Sébastien Thériault, Stefan van Duijvenboden, Helen R. Warren, Jie Yao, Dania Raza, Stefanie Aeschbacher, Gustav Ahlberg, Alvaro Alonso, Laura Andreasen, Joshua C. Bis, Eric Boerwinkle, Archie Campbell, Eulalia Catamo, Massimiliano Cocca, Michael J. Cutler, Dawood Darbar, Alessandro De Grandi, Antonio De Luca, Jun Ding, Christina Ellervik, Patrick T. Ellinor, Stephan B. Felix, Philippe Froguel, Christian Fuchsberger, Martin Gögele, Claus Graff, Mariaelisa Graff, Xiuqing Guo, Torben Hansen, Susan R. Heckbert, Paul L. Huang, Heikki V. Huikuri, Nina Hutri-Kähönen, M.Arfan Ikram, Rebecca D. Jackson, Juhani Junttila, Maryam Kavousi, Jan A. Kors, Thiago P. Leal, Rozenn N. Lemaitre, Henry J. Lin, Lars Lind, Allan Linneberg, Simin Liu, Peter W. MacFarlane, Massimo Mangino, Thomas Meitinger, Massimo Mezzavilla, Pashupati P. Mishra, Rebecca N. Mitchell, Nina Mononen, May E. Montasser, Alanna C. Morrison, Matthias Nauck, Victor Nauffal, Pau Navarro, Kjell Nikus, Guillaume Pare, Kristen K. Patton, Giulia Pelliccione, Alan Pittman, David J. Porteous, Peter P. Pramstaller, Michael H. Preuss, Olli T. Raitakari, Alexander P. Reiner, Antonio Luiz P. Ribeiro, Kenneth M. Rice, Lorenz Risch, David Schlessinger, Ulrich Schotten, Claudia Schurmann, Xia Shen, M.Benjamin Shoemaker, Gianfranco Sinagra, Moritz F. Sinner, Elsayed Z. Soliman, Monika Stoll, Konstantin Strauch, Kirill Tarasov, Kent D. Taylor, Andrew Tinker, Stella Trompet, André Uitterlinden, Uwe Völker, Henry Völzke, Melanie Waldenberger, Lu-Chen Weng, Eric A. Whitsel, James G. Wilson, Christy L. Avery, David Conen, Adolfo Correa, Francesco Cucca, Marcus Dörr, Sina A. Gharib, Giorgia Girotto, Niels Grarup, Caroline Hayward, Yalda Jamshidi, Marjo-Riitta Järvelin, J.Wouter Jukema, Stefan Kääb, Mika Kähönen, Jørgen K. Kanters, Charles Kooperberg, Terho Lehtimäki, Maria Fernanda Lima-Costa, Yongmei Liu, Ruth J.F. Loos, Steven A. Lubitz, Dennis O. Mook-Kanamori, Andrew P. Morris, Jeffrey R. O’Connell, Morten Salling Olesen, Michele Orini, Sandosh Padmanabhan, Cristian Pattaro, Annette Peters, Bruce M. Psaty, Jerome I. Rotter, Bruno Stricker, Pim van der Harst, Cornelia M. van Duijn, Niek Verweij, James F. Wilson, Dan E. Arking, Julia Ramirez, Pier D. Lambiase, Nona Sotoodehnia, Borbala Mifsud, Christopher Newton-Cheh, Patricia B. Munroe

**Author notes:** jointly supervised this project (Nona Sotoodehnia, Borbala Mifsud, Christopher Newton-Cheh, Patricia B. Munroe).

## Abstract

The QT interval is an electrocardiographic measure representing the sum of ventricular depolarization (QRS duration) and repolarization (JT interval). Abnormalities of the QT interval are associated with potentially fatal ventricular arrhythmia. We conducted genome-wide multi-ancestry analyses in >250,000 individuals and identified 177, 156 and 121 independent loci for QT, JT and QRS, respectively, including a male-specific X-chromosome locus. Using gene-based rare-variant methods, we identified associations with Mendelian disease genes. Enrichments were observed in established pathways for QT and JT, with new genes indicated in insulin-receptor signalling and cardiac energy metabolism. In contrast, connective tissue components and processes for cell growth and extracellular matrix interactions were significantly enriched for QRS. We demonstrate polygenic risk score associations with atrial fibrillation, conduction disease and sudden cardiac death. Prioritization of druggable genes highlighted potential therapeutic targets for arrhythmia. Together, these results substantially advance our understanding of the genetic architecture of ventricular depolarization and repolarization.

## Main

The electrocardiogram (ECG) is a non-invasive tool that captures cardiac electrical activity^1^. The QT interval (QT) represents the sum of ECG measures for ventricular depolarization (QRS duration; QRS) and repolarization (JT interval; JT) (Fig. 1a). The QT is used to diagnose congenital long or short QT syndromes and acquired QT-prolongation, which are associated with an increased risk for ventricular arrhythmia and sudden cardiac death (SCD)^2–5^. Susceptibility to congenital long QT syndrome (LQTS) is mediated by rare and common variation at 15 genes including *KCNQ1*, *KCNH2* and *SCN5A*^6, 7^. However, 25-30% of LQTS cases have a negative genetic test and LQTS genes do not adequately explain the heritability of the QT in the general population, or predisposition to QT-prolongation from precipitating factors such as medication^8, 9^.

**Figure 1:**
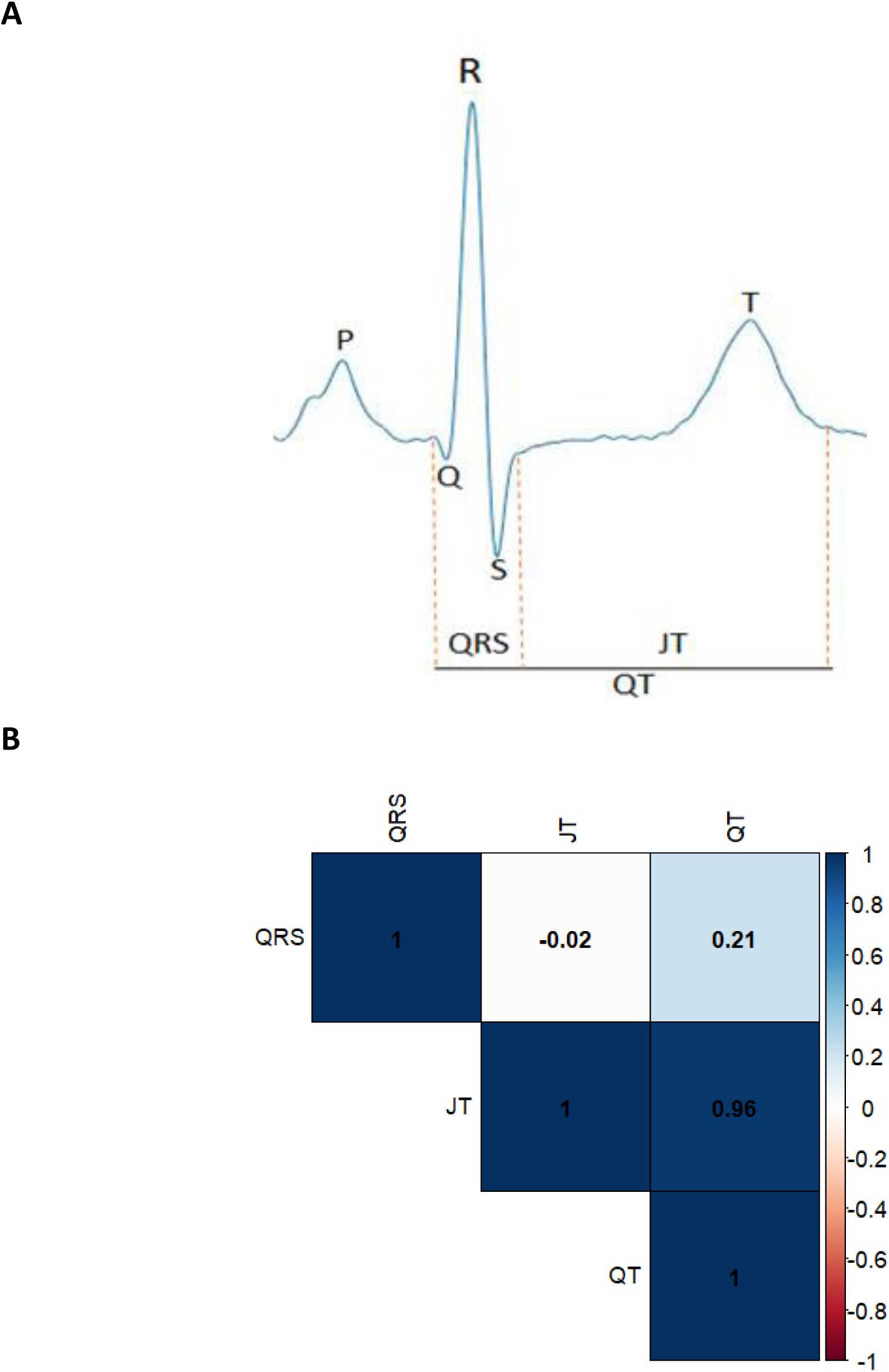
Relationship and phenotypic correlations of QT, JT and QRS. A) Representation of each ECG measure on a single cardiac beat. B) Phenotypic correlations calculated in ∼51K UK Biobank individuals of European ancestry. Spearman’s rank (r_s_) correlation coefficients are reported.

QT and JT phenotypes are highly correlated, whereas QRS has a modest positive and a negligible negative correlation with QT and JT respectively (Fig. 1b)^10^. Therefore, the investigation of QT and its individual components has potential to identify both shared and specific biological mechanisms for ventricular depolarization and repolarization. In genome-wide association studies (GWAS) for QT and JT, common variants at genes regulating cardiac ion channels, calcium-handling proteins and myocyte structure have been reported^11, 12^. GWAS for QRS have highlighted genes for sodium channels, kinase inhibitors and transcription factors in cardiac embryonic development^13, 14^. However, a large proportion of the heritability remains unexplained (∼67% for QT, ∼83% for QRS), and our limited understanding of the underlying biological networks restricts the potential translational opportunities^15–17^.

We have performed the largest multi-ancestry GWAS meta-analysis to date for QT, JT and QRS in over 250,000 individuals, to discover additional candidate genes and pathways relevant to ventricular depolarization and repolarization, identify new therapeutic targets, and test the association of polygenic risk scores (PRSs) with cardiovascular disease.

## Results

### Meta-analysis of GWAS

Thirty-five studies contributed to our primary multi-ancestry GWAS meta-analysis for QT, JT, and QRS, comprising a maximum total of 252,977 individuals of European (84%), Hispanic (7.7%), African (6.7%), South and South-East Asian (<1%) ancestries (Supplementary Tables 1–3, Supplementary Note 1). The meta-analysis workflow is summarized in Fig. 2. No evidence of inflation of test statistics was observed (Supplementary Fig. 1 and 2).

**Figure 2:**
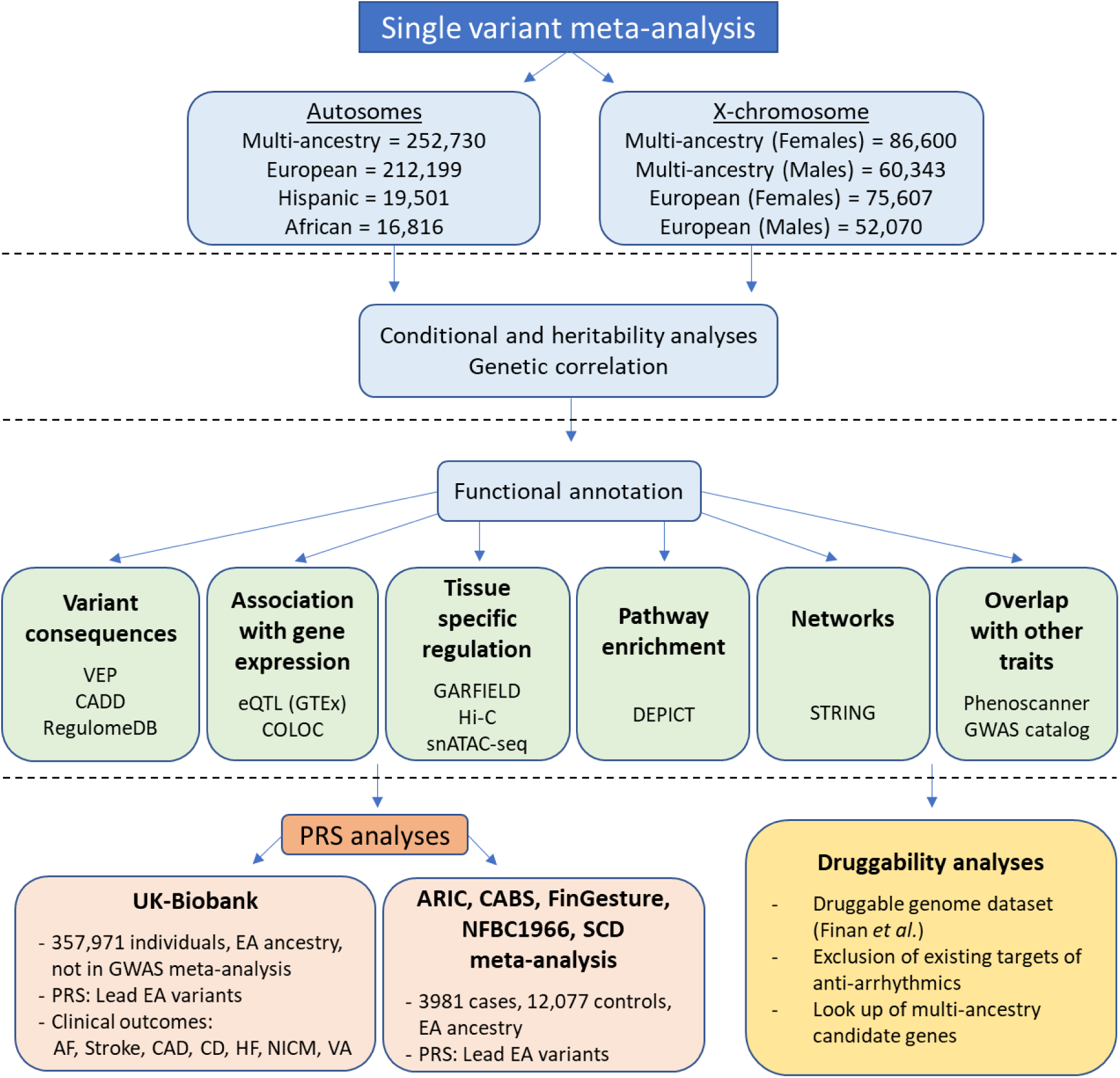
Workflow for single variant analyses of QT, JT and QRS. Workflow for single variant meta-analysis and downstream bioinformatics. VEP (Variant Effect Predictor), CADD (Combined Annotation Dependent Depletion), eQTL (expressive Quantitative Trait Locus), GTEx (Genotype-Tissue Expression project), COLOC (Colocalization), GARFIELD (GWAS Analysis of Regulatory and Functional Information Enrichment with LD correction), DEPICT (Data-driven Expression-Prioritized Integration for Complex Traits), STRING (Search Tool for Retrieval of Interacting Genes/Proteins), GWAS (Genome-Wide Association Study), EA (European Ancestry), PRS (Polygenic Risk Score), AF (Atrial Fibrillation), CAD (Coronary Artery Disease), CD (Conduction Disease), HF (Heart Failure), NICM (Non-Ischaemic Cardiomyopathy), VA (Ventricular Arrhythmia), SCD (Sudden Cardiac Death)

### QT GWAS meta-analysis

We discovered 176 genome-wide significant (GWS; *P*-value (*P*) < 5×10^-8^) lead variants at independent autosomal loci (114 unreported) associated with QT in the multi-ancestry meta-analysis (Table 1, Supplementary Fig. 3a). Of the previously reported loci for QT or JT (grouped as the phenotypic correlation is high), there was support for association at 62/66 (93.9%) loci (*P* < 5×10^-8^). There was weaker support for association at 3 loci (*NRAP*, *MYH6*, *NACA*, *P* < 1.29×10^-4^) but not for *SUCLA2* (*P* > 0.05) (Supplementary Table 4). Ancestry-specific analyses identified additional unreported loci in European (11), Hispanic (1), and African (1) individuals (Supplementary Table 5). All European and African ancestry-specific lead variants were supported in the multi-ancestry analysis (*P* < 5 x 10^-5^). The Hispanic ancestry locus was not supported in the multi-ancestry analysis (*P* = 0.07), however, the lead variant at this locus was rare (MAF = 0.002) and monomorphic in Europeans.

**Table 1:**
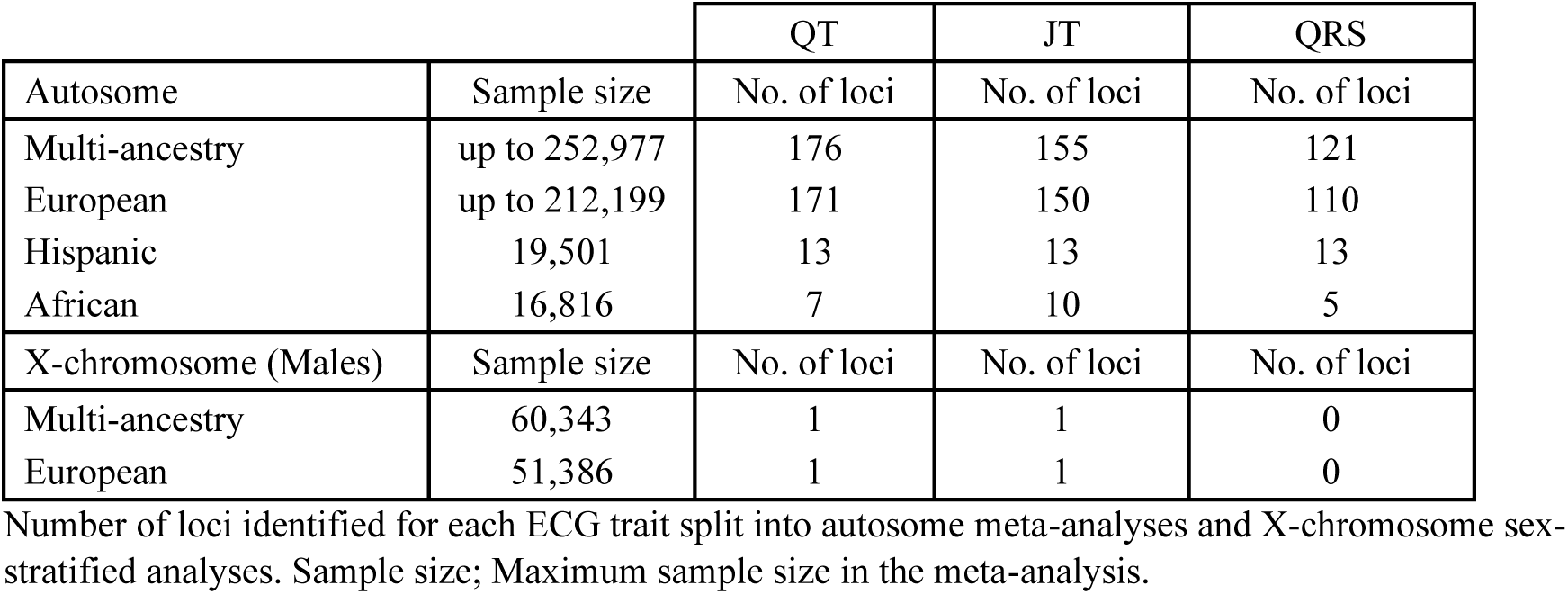
Number of loci identified in each QT, JT and QRS meta-analysis.

To identify additional signals, we performed joint and conditional analyses with Genome-wide Complex Trait Analysis (GCTA)^18^ in 52,230 individuals of European ancestry from the UK Biobank (UKB) using European ancestry meta-analysis summary statistics. These analyses identified an additional 65 conditionally independent variants at 38 loci at *P_joint_* < 5×10^-8^ (Supplementary Table 6).

### JT and QRS GWAS meta-analyses

In JT and QRS GWAS, we identified 155 and 121 lead variants at independent autosomal loci (96 and 77 unreported) in multi-ancestry meta-analyses respectively (Table 1, Supplementary Fig. 3, Supplementary Tables 7 and 8). Ancestry-specific analyses identified additional unreported loci (N=18) in European (4 JT, 6 QRS), African (4 JT, 2 QRS) and Hispanic (1 JT, 1 QRS) ancestries. Of these, 13 lead variants had evidence for support in the multi-ancestry analysis (*P* < 5 x 10^-5^). Joint and conditional analyses identified an additional 56 and 29 conditionally independent variants at 32 JT and 18 QRS loci respectively (Supplementary Table 6).

### X-chromosome meta-analyses

X-chromosome analyses (multi-ancestry sample size: 86,600 and 60,343 for separate female and male analyses respectively) identified one locus in males for both multi- and European ancestry meta-analyses, for QT and JT (Table 1, Supplementary Table 5 and 7). There were no GWS findings for QRS or female X-chromosome analyses, and no suggestive evidence of association on a lookup of the lead QT/JT variant (rs55891214) in these analyses (*P* > 0.05). rs55891214 is highly correlated (r^2^ > 0.9) with lead variants reported in GWAS for serum testosterone, estradiol levels, male-pattern baldness, and heel bone mineral density^19–22^. The nearest gene, *FAM9B*, is exclusively expressed in the testis, and together these findings suggest the association may be driven by serum testosterone levels^19, 23^.

### Overlap of genetic contributions and heritability of QT, JT, and QRS

There was substantial overlap of multi-ancestry JT and QT GWAS loci (130/200, 65%) but less between QRS and QT (53/243, 21.8%) (overlap: r^2^ > 0.1 between lead variants or within ± 500 kb). For QRS and JT, there was overlap at 34 loci, where a lead variant was genome-wide significant in both analyses (Supplementary Table 9). Predominantly discordant (27/34, 79.4%) directions of effect were observed at these lead variants (Fig. 3). Across all loci for QT, JT and QRS, overlap was observed with previously reported loci for PR interval (51 (29.1%), 46 (29.7%), 42 (34.7%), respectively) and resting heart rate (58 (33.1%), 57 (36.8%), 46 (38.0%)), demonstrating shared genetic contributions. 13 loci were common to all 5 ECG measures (Supplementary Table 10), highlighting these loci as integral genetic determinants of global cardiac electrophysiology. Estimated genetic correlations were calculated using LD Score Regression (LDSC)^24, 25^. A strong positive correlation was observed between QT and JT (r_g_ = 0.91, *P* < 0.001) and a weak positive correlation between QT and QRS (r_g_ = 0.17, *P* = 0.05) (Supplementary Fig. 4). In contrast, a negative genetic correlation was observed between JT and QRS (r_g_ = -0.25, *P* = 0.003).

**Figure 3:**
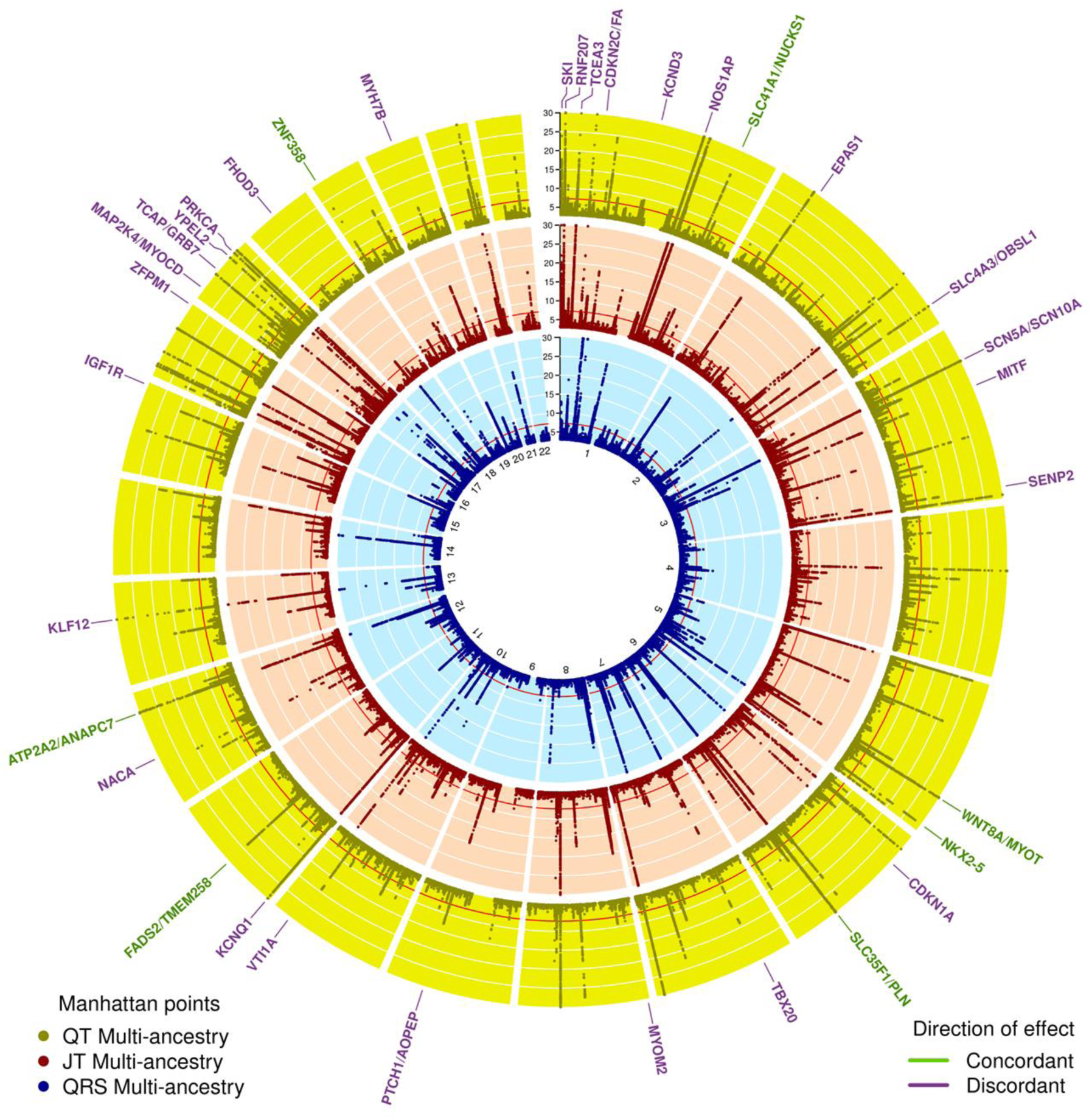
Circular Manhattan plot for QT, JT and QRS multi-ancestry meta-analyses. Circular Manhattan plots for QT (outer), JT (middle) and QRS (inner) multi-ancestry meta-analysis. The Y axis has been restricted to -log10 P-value < 30. Overlapping JT and QRS loci are labelled with the most likely candidate at the locus. Direction of effect was compared by comparing the lead JT variant beta with the corresponding direction of effect of the same variant in the QRS GWAS meta-analysis. This plot was produced using the R package Circlize version 0.4.10. Gu, Z. (2014) circlize implements and enhances circular visualization in R. Bioinformatics. 10.1093/bioinformatics/btu393

SNP-based heritability estimations in Europeans from UKB for QT, JT and QRS were 29.3%, 29.5% and 15.0% (standard error [SE]: 1%) respectively. The percentage of overall variance explained by all lead and conditionally independent variants from the European meta-analysis was 14.6%, 15.9% and 6.3%. Therefore, these variants explain 49.8%, 53.9% and 42.0% of the SNP-based heritability of QT, JT and QRS.

### Gene-based meta-analysis

To investigate whether rare variants (MAF< 0.01) in aggregate modulate ECG traits, we conducted gene-based meta-analyses of rare variants predicted by Variant Effect Predictor (VEP)^26^ to have high or moderate impact on protein function, using Sequence Kernel Association Testing (SKAT)^27^. These analyses discovered 13, 16 and 3 genes for QT, JT and QRS respectively (*P* <2.5×10^- 6^; Bonferroni adjusted for ∼20,000 genes). These genes were brought forward for conditional analyses, and 7, 7 and 2 genes remained associated with QT, JT and QRS respectively after conditioning on the rare variant with the lowest *P*-value at each gene (*P* < 0.05/number of genes) (Table 2). These results indicate that the gene-based associations were not a consequence of a single variant with a strong effect. A burden of rare coding variants at three Mendelian long-QT syndrome (LQTS) genes (*KCNQ1*, *KCNH2* and *SCN5A*) was associated with QT and/or JT. *SCN5A* was also associated with QRS. *MYH7* and *TNNI3K* were associated with JT. *TNNI3K*, which was not associated using single variant analysis, encodes a cardiomyocyte-specific kinase previously linked to familial cardiac arrhythmia and dilated cardiomyopathy (OMIM: 613932)^28, 29^.

**Table 2:** Significant genes from gene-based meta-analysis for each ECG trait following conditional analysis.

|  | Gene | N | P-value<br>Unconditional | No. of<br>variants | Beta | SD | Conditioned<br>variant | P-value<br>after<br>conditioning |
| --- | --- | --- | --- | --- | --- | --- | --- | --- |
| QT | *KCNH2* | 180,961 | 9.11E-12 | 43 | 0.15 | 0.022 | 7:150654525-G-A | 1.68E-07 |
|  | *NOS3* | 183,747 | 1.04E-10 | 68 | 0.03 | 0.012 | 7:150698349-G-A | 1.65E-05 |
|  | *KCNQ1* | 158,377 | 1.23E-11 | 36 | 0.06 | 0.047 | 11:2790111-C-T | 1.75E-05 |
|  | *RNF207* | 168,015 | 9.46E-16 | 39 | -0.15 | 0.020 | 1:6279316-C-T | 3.89E-05 |
|  | *OLFML2B* | 183,747 | 5.30E-12 | 46 | 0.06 | 0.012 | 1:161970046-A-G | 6.85E-04 |
|  | *TSSC4* | 189,264 | 5.16E-18 | 25 | -0.10 | 0.014 | 11:2424684-A-C | 1.97E-03 |
|  | *MYH7* | 173,501 | 3.13E-07 | 38 | 0.05 | 0.025 | 14:23892910-A-G | 3.58E-03 |
| JT | *SCN5A* | 181,936 | 2.53E-15 | 90 | -0.05 | 0.013 | 3:38591853-A-G | 1.03E-05 |
|  | *NOS3* | 183,468 | 5.13E-12 | 68 | 0.04 | 0.012 | 7:150698349-G-A | 1.60E-05 |
|  | *RNF207* | 167,737 | 1.33E-16 | 39 | -0.16 | 0.020 | 1:6279316-C-T | 5.55E-05 |
|  | *MYH7* | 173,223 | 1.48E-08 | 38 | 0.06 | 0.025 | 14:23892910-A-G | 1.47E-04 |
|  | *KCNQ1* | 158,099 | 5.01E-11 | 36 | 0.10 | 0.047 | 11:2790111-C-T | 3.09E-04 |
|  | *TNNI3K\** | 181,936 | 3.48E-08 | 56 | 0.03 | 0.016 | 1:74929170-T-C | 2.79E-03 |
|  | *OLFML2B* | 183,468 | 1.35E-12 | 46 | 0.07 | 0.012 | 1:161970046-A-G | 2.91E-03 |
| QRS | *SCN5A* | 181,930 | 3.34E-11 | 90 | 0.04 | 0.013 | 3:38591853-A-G | 7.89E-06 |
|  | *DLEC1* | 188,621 | 1.05E-06 | 117 | 0.05 | 0.012 | 3:38163900-C-T | 2.91E-04 |

N, total sample size at gene-based meta-analysis for each gene; P-value unconditional, Gene-based P-value for association with the ECG trait; No. of variants, Number of rare variants included in burden testing; SD, Standard Deviation; Conditioned variant, Variant with the smallest P-value used for conditional analysis to test whether the association was driven by a single variant; P-value after conditioning, Gene-based P-value after conditioning on the variant with the smallest P-value. Thresholds: Unconditional P-value significant if $< 2.5 \times 10^{-6}$ (Bonferroni corrected for ~20,000 genes). P-value after conditioning significant if $< 0.05/\text{number of genes}$ brought forward for each ECG trait. \*Gene within a locus previously unreported for these ECG measures using single-variant GWAS methods.

As several genes mapped to loci implicated in single variant GWAS analyses, we explored the relationship between these rare gene-based signals and common (MAF > 0.05) or low frequency (0.01≤ MAF ≤0.05) variants. Analyses were repeated in 76,202 individuals from UKB conditioning on independent significant variants identified in the corresponding European GWAS meta-analysis, and residing within the same locus as the gene. These conditional analyses showed that associations for *KCNQ1*, *KCNH2*, and *RNF207* with QT and/or JT were independent of flanking variants identified by GWAS (Supplementary Table 11). Because conditional analyses required a large sample with a shared set of variants, we lacked adequate power to definitively determine the independence of *DLEC1* from nearby common variants. For *MYH7* and *TNNI3K*, there were no GWS variants at the locus in our single-variant meta-analysis.

### Biological annotation of GWAS loci

#### Variant-level functional annotation

Most multi-ancestry QT lead variants, 160/176 (90.9%), were common, 13 were low frequency and 3 were rare (*RUFY1*, *DNAJB5*, *CACNB2*; Supplementary Table 5). At 25 loci, a lead variant or proxy (r^2^ >0.8) was annotated with VEP, as missense (N = 24) or stop-gain (N = 1) (Supplementary Table 12a). Ten of these (40%) were predicted by SIFT or PolyPhen-2 to be “deleterious” or “damaging”. These included: rs1805128, a *KCNE1* polymorphism D85N (c.253G>A), which is a recognized modifier of Long QT syndrome^30^, and a missense variant in *NEXN* (rs1166698). *NEXN* mutations are associated with cardiomyopathies (OMIM: 613121)^31, 32^. At 16 loci, a lead variant or proxy had a Combined Annotation Dependent Depletion (CADD) score ≥20 and was thus predicted to be among the top 1% most deleterious in the genome (Supplementary Table 12b). Of these, the variants at 10 loci were non-coding.

A similar proportion of lead variants for JT (91%) and QRS (95.9%) were common. Missense variants were identified in 21 JT and 11 QRS loci (Supplementary Table 12a) and predicted to be “deleterious” or “damaging” at 8 JT and 7 QRS loci. At 18 JT and 13 QRS loci, the lead/proxy variant had a CADD score ≥20 (Supplementary Table 12b). Of these, the variant was non-coding in 13 JT and 7 QRS loci.

#### Association with gene expression levels in cardiac tissue

Using data from the Genotype-Tissue Expression (GTEx) project^33^, we identified 39 (22.2%) multi-ancestry QT loci where a variant was a cis-eQTL in left ventricle (LV) or right atrial appendage (RAA) tissue samples (Supplementary Table 13). There was strong support for pairwise colocalization (posterior probability >0.75) at 17 loci for the LV and 14 for the RAA.

At 37 (23.9%) JT and 18 (14.9%) QRS multi-ancestry loci the lead variant or proxy was a significant cis-eQTL in LV or RAA tissue (Supplementary Table 13). There was support for pairwise colocalization at 12 JT and 7 QRS loci in LV tissue and 12 JT and 8 QRS loci in RAA. Comparing QT/JT and QRS, discordant directions of effect were identified at 3 overlapping loci (*KLF12* [RAA], *PRKCA* and *TCEA3* [RAA and LV]), suggesting that differences at a variant level may translate to opposing effects on tissue-specific gene expression (Fig. 4).

**Figure 4:**
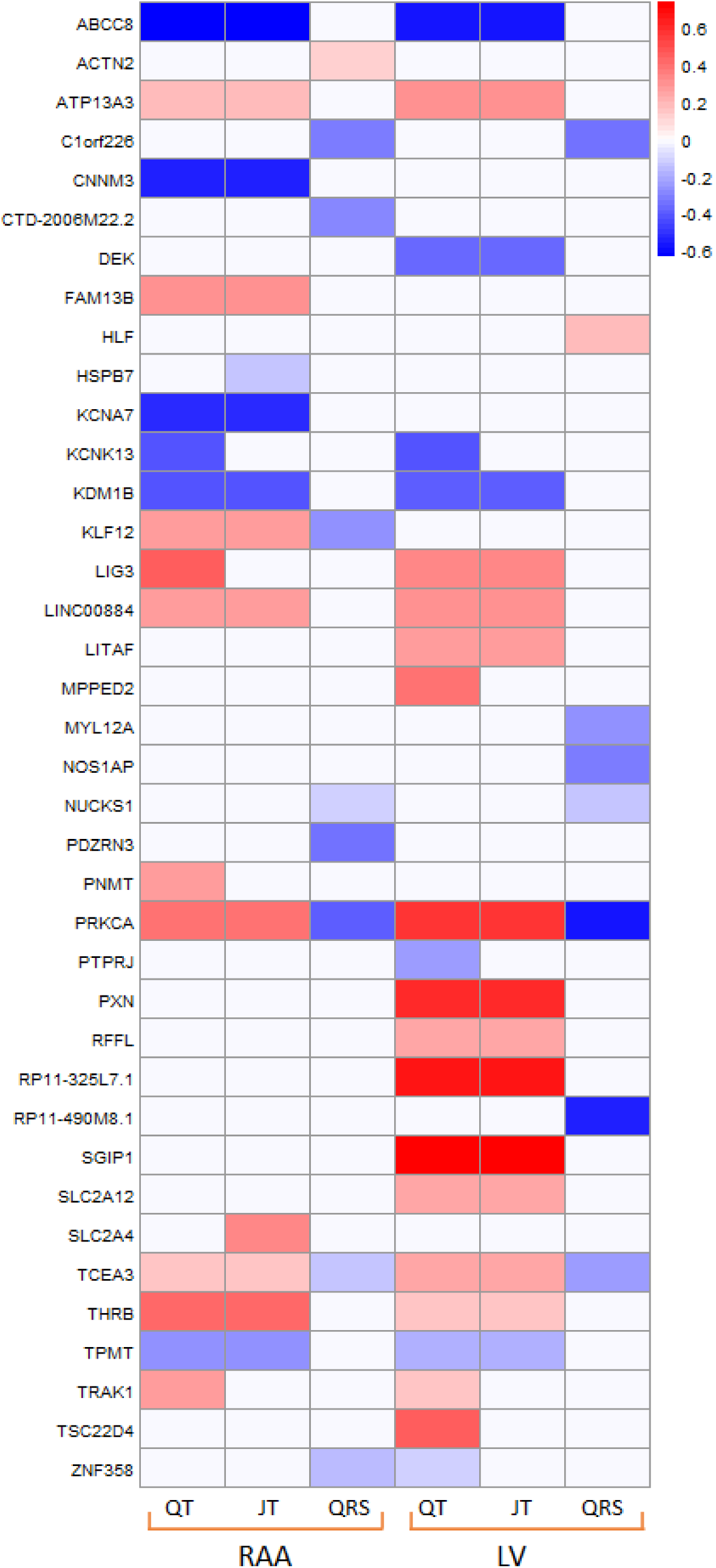
Comparison of co-localised eQTL signals for QT, JT and QRS in right atrial appendage and left ventricle tissues. Colocalization analyses performed using data from GTEx (version 8), (methods). The direction of effect has been aligned to the ECG trait prolonging allele. Y axis: Transcripts. RAA: Right atrial appendage, LV: Left ventricle

#### Tissue- and cell-type specific effects of variants through regulatory elements

Potential target genes of regulatory variants were identified using two long-range chromatin interaction datasets (40 kb-resolution Hi-C and ∼4 kb-resolution promoter-capture Hi-C) from left and right ventricular tissue^34, 35^. Promoter interactions were identified at 39 (22.2%) QT loci (Supplementary Tables 14a and 14b). Evaluation of cardiac cell-type specific effects, using single nucleus Assay for Transposase-Accessible Chromatin using sequencing (snATAC-seq) data^36^, identified significant enrichment at open chromatin regions for QT in atrial and ventricular cardiomyocytes (Supplementary Fig. 5).

Promoter interactions were identified at 46 (29.7%) JT and 28 (23.1%) QRS loci (Supplementary Tables 14a and 14b). Cardiac cell-type specific enrichment was significant in atrial and ventricular cardiomyocytes for both JT and QRS (Supplementary Fig. 5). There was also enrichment in adipocytes for JT.

Tissue-specific enrichment of variants in DNaseI hypersensitivity sites, using GWAS Analysis of Regulatory and Functional Information Enrichment with LD correction (GARFIELD, v2)^37^ identified strongest enrichment in fetal heart tissue for QT, JT and QRS (Supplementary Fig. 6).

#### Gene-set tissue/cell-type enrichment and pathway analyses

Candidate genes were prioritized to common functional pathways using reconstituted gene-sets in Data-driven Expression-Prioritization Integration for Complex Traits (DEPICT) software^38^. These gene-sets were highly expressed (FDR<0.01) in cardiac tissues for all three traits (Supplementary Table 15). Additionally, for QRS, connective tissue cell-types including cardiac valve, chondrocytes, and joint/skeletal tissues, were identified (Supplementary Fig. 7).

The most significant (FDR<0.01) gene-ontology (GO) biological processes for QT could be grouped into broad categories, including cardiac and muscle cell differentiation/development, and regulation of gene expression (Supplementary Fig. 8). In addition, and previously not reported for QT, response to insulin and insulin receptor signalling processes were identified (Fig. 5). In the Reactome database, top pathways were related to signal transduction, including protein kinase B (PKB) mediated events, mechanistic target of rapamycin (mTOR) signalling, the phosphoinositide 3-kinases (PI3K) cascade and the insulin receptor signalling (IRS) cascade (Supplementary Table 16). The top 10 mouse phenotypes enriched for these gene-sets included abnormal myocardial layer/trabeculae morphology, decreased embryo size/growth retardation, and increased heart weight.

**Figure 5:**
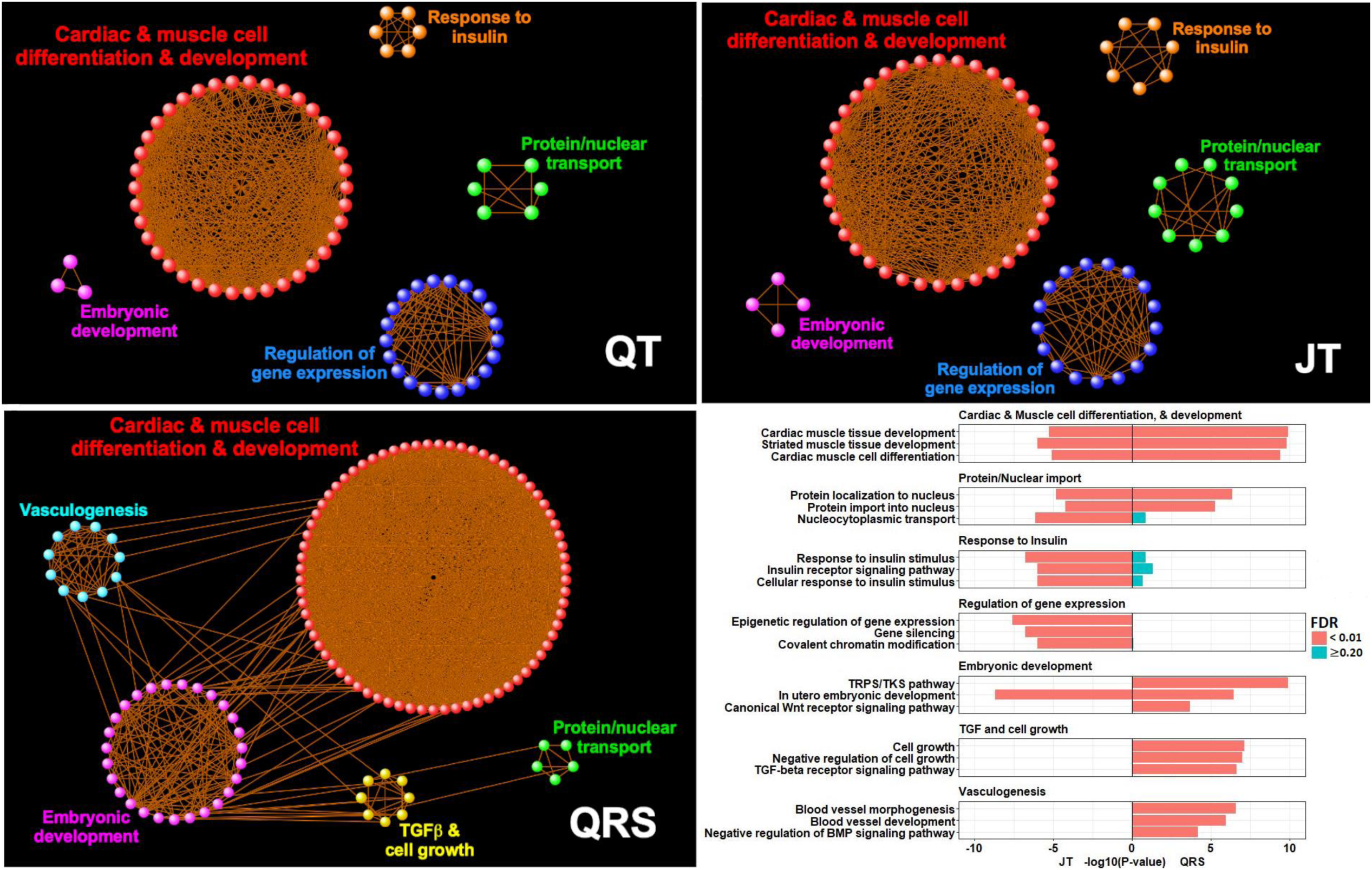
Enrichment network visualisation of DEPICT GO biological processes. First three panels (QT, JT and QRS) were created using Cytoscape (v3.8.2) and display common themes among pathways. Modules were created by connecting significant GO biological processes (FDR < 0.01) with overlapping gene members. Orphan pathways or those with less than three edges were excluded. The final panel shows a bar graph with example GO process members (y-axis) for JT and QRS from each “common theme”, along with their enrichment *P*-values (x-axis) and color coded by FDR. TGF-beta: Transforming growth factor beta, TRPS/TKS: transmembrane receptor protein serine/threonine kinase, BMP: Bone morphogenic protein.

GO biological processes significant for JT (and QT), but not QRS, included regulation of gene expression, histone and chromatin modification, insulin receptor signalling and response to insulin stimulus (Fig. 5, Supplementary Table 16). Cellular growth, the transmembrane receptor protein serine (TRPS)/threonine kinase (TK) signalling pathway and vasculogenesis were enriched only for QRS. Reactome pathways significant for JT (and QT) but not QRS included regulation of lipid metabolism by peroxisome proliferator and its activated receptor effect on gene expression, along with insulin receptor signalling and related events (Supplementary Fig. 9). The top Reactome pathway for QRS, not significant for JT (or QT), was extracellular matrix interactions^39^. A summary of all bioinformatic analyses for previously unreported loci is in Supplementary Table 17-19.

### Identification of potential drug targets for therapeutic opportunities

To identify potential new drug targets for arrhythmia, we interrogated the druggable gene set database published by Finan et al^40^. We examined all 200, 173 and 155 plausible candidate genes from the QT, JT and QRS multi-ancestry meta-analyses respectively, including known and previously unreported loci (Supplementary Tables 4, 17-19). 53 (QT), 46 (JT), and 31 (QRS) genes were identified as potential drug targets, that are not current targets of anti-arrhythmic drugs (Supplementary Table 20). Of these, 21 QT, 17 JT, and 10 QRS genes were classed as Tier 1, encoding proteins that are targets of drugs either approved or in clinical development. Genes from significant signals in cardiac tissue-specific eQTL co-localization and Hi-C analyses may be favoured for prioritization. Of the 53 potential gene therapeutic targets for QT, *ABCC8*, *KCNA7*, *KCNK13*, *PRKCA* and *THRB* had support for co-localization in eQTL analyses and *NFKB1*, *MITF*, *PLK2* and *CASQ2* were significant Hi-C findings. *CASQ2* and *PLK2* were previously investigated as potential targets of gene transfer technology^41, 42^.

### Association of genetically determined QT, JT, and QRS with cardiovascular disease and sudden cardiac death

PRSs were constructed using European-ancestry lead variants. Each PRS was tested for association with the directly measured ECG trait in 4,214 individuals from UKB not included in the GWAS. Associations observed for each PRS were (β [95% CI]): 6.4ms (5.7–7.1) for QT; 6.4ms (5.7– 7.1) for JT; and 2.2ms (1.7–2.7) for QRS (ms per standard deviation [SD] increase in the PRS).

In ∼357K unrelated individuals of European ancestry from UKB not included in the GWAS meta-analysis, each PRS was tested for association with prevalent cardiovascular disease cases including atrial fibrillation (AF), “atrioventricular block (AVB) or pacemaker implantation (PPM)”, “bundle branch block (BBB) or fascicular block”, and heart failure (Supplementary Table 21, Supplementary Note 2, Fig. 6). A Bonferroni threshold (0.05/number of conditions tested) was used to indicate significance (*P* < 6.3 x 10^-3^). Genetically determined QT and JT were associated with decreased risk for “AVB or PPM implantation” (odds ratio [OR] (95% CI) per SD: 0.94 [0.924–0.963] and 0.94 [0.918–0.956] respectively). In contrast, genetically determined QRS was associated with increased risk for “BBB or fascicular block” (1.07 [1.037–1.105]). Genetically determined QT and QRS were associated with decreased risk for AF (0.97 [0.954–0.979]) and 0.93 [0.921–0.945] respectively). Including the QRS PRS as a covariate in the QT PRS model did not substantially change the points estimate (OR: 0.97 [0.958-0.983]), indicating the relationship with AF was not driven by overlap with the genetic contribution for QRS.

**Figure 6:**
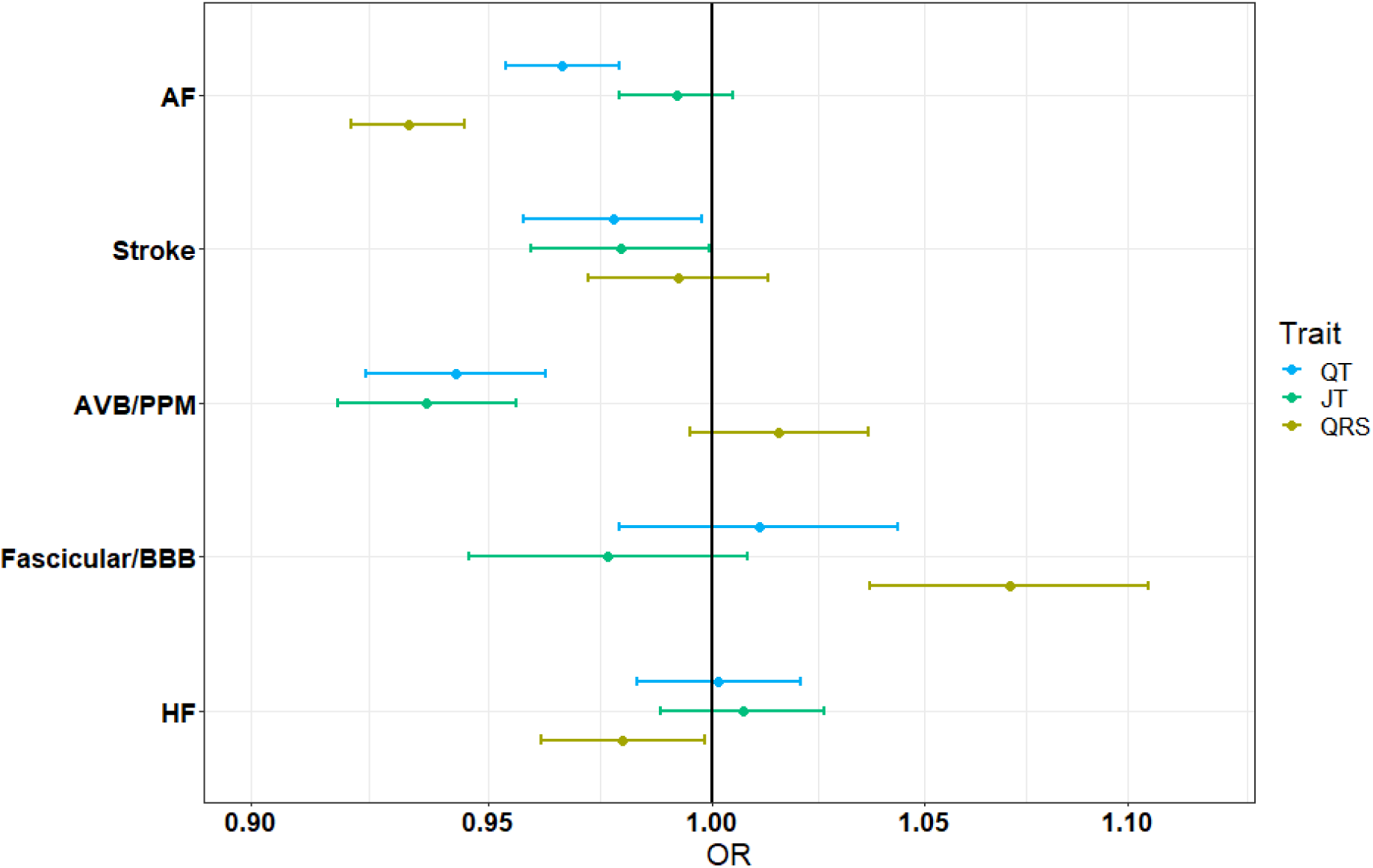
Odds ratios and confidence intervals for ECG PRS with clinical outcomes in UK Biobank. Odds ratios (OR) and 95% confidence intervals (x-axis) for association of each ECG polygenic risk score (PRS) with prevalent cases in UK Biobank. Associations are reported as risk per standard deviation increase in the PRS. A total of 371,951 individuals of European ancestry were included in this analysis. AF (Atrial Fibrillation), AVB (Atrioventricular block), PPM (Pacemaker), BBB (Bundle branch block), HF (Heart Failure).

The PRSs were also tested for association with SCD in the Atherosclerosis Risk in Communities (ARIC) study, Cardiac Arrest Blood Study (CABS), Finnish Genetic Study for Arrhythmic Events (FinGesture) and Northern Finland Birth Cohort of 1966 (NFBC1966) cohorts (Supplementary note 3). To allow the meta-analysis of these studies, results are reported as risk for SCD, per unit increase in the average ms per allele. The distribution of the PRS (mean [SD]) was: 0.321 (0.02) for QT; 0.361 (0.02) for JT; 0.134 (0.008) for QRS. Therefore, findings are reported as log OR (95% CI). The lead variant at the *NOS1AP* locus for QT and JT (rs12042862, T-allele) was associated with increased risk for SCD (0.11 [0.036–0.190], *P*= 0.004), as previously reported^43^. There was no association observed between each PRS and SCD in the full sample (Supplementary Table 22). As the incidence of SCD is different between men and women, we performed sex-stratified analyses^44^. Increasing QT PRS was associated with SCD in females (8.2 [3.05-13.35], *P* = 1.8×10^-3^) with a concordant direction of effect across all studies. Sensitivity analyses in FinGesture suggested that the association between the QT PRS and SCD in women, may be driven by non-ischaemic aetiologies compared with ischaemic (*P*= 0.004 vs. 0.926 respectively).

## Discussion

Our large-scale GWAS meta-analyses for QT and its components, QRS and JT, substantially advance our understanding of the genetic architecture of ventricular depolarization and repolarization. We more than double the number of autosomal loci associated with each trait and identify sex-specific effects at an X-chromosome locus (*FAM9B*). In addition to established processes, we report loci involved in energy metabolism and response to insulin, which have greater enrichment for ventricular repolarization. Extracellular matrix interactions, cell growth and connective tissue components are significantly enriched among QRS-associated genes, and therefore ventricular depolarization. We identify Mendelian genes for which a burden of rare variants are associated with these ECG measures (e.g. *KCNQ1*, *KCNH2, TNNI3K*). We also highlight potential new therapeutic targets and together with the association of PRSs with AF, conduction disease and SCD, these indicate possible translational opportunities of our findings.

Previous knowledge of ventricular repolarization has centered on the role of cardiac ion channels, predominantly from the investigation of inherited arrhythmic syndromes^3^. However, ventricular repolarization is complex and influenced by multiple processes, as suggested by previous GWAS and now advanced in our present study^11, 12^. Our analyses have identified additional candidate genes involved in cardiomyocyte differentiation, tissue development, cardiac contraction and regulation of gene expression. In this study we also report pathways related to insulin receptor and mTOR signalling, along with genes that implicate cardiac energy metabolism. Ion homeostasis is an energy-consuming process and mismatch in the supply and utilization of adenosine triphosphate (ATP) can lead to electrical and mechanical instability^45^. Appropriate cardiac energy utilization is therefore, necessary to maintain normal physiological activity. Candidate genes common to QT and JT within insulin related pathways include *SLC2A4*, *PIGQ* and *ABCC8*. *SLC2A4* (alias *GLUT4*), is a glucose transporter in cardiomyocytes with effects on cardiac contractility, development of hypertrophy, and susceptibility to atrial and ventricular arrhythmias^46–48^. *PIGQ* is involved in the biosynthesis of glycosylphosphatidylinositol-anchored proteins, which are involved in membrane protein transportation and cell surface protection^49^. Mutations involving these proteins cause cardiac related glycosylation disorders, including congenital defects and arrhythmia^50^. *ABCC8* (alias sulfonylurea receptor 1) modulates ATP-sensitive potassium channels and insulin release. It is a crucial component of sarcolemma K^+^ ATP channels in mouse atrial myocytes, however a similar role has not been identified in human cardiomyocytes^51, 52^. In humans, the role of *ABCC8* is predominantly within pancreatic beta cells, and therefore may indicate indirect effects on ventricular repolarization through insulin secretion^53^. Insulin is considered cardioprotective and receptors for insulin signalling are highly expressed in the heart^54^. Type 1 diabetic mice have altered ion channel kinetics in ventricular and atrial myocytes, that increase the risk of arrhythmia (including AF) and can be reversed with insulin therapy^55–57^.

Sex-differences in ventricular repolarization are well recognised and incorporated into clinical definitions for QT-prolongation^58^. Our study identified an X-chromosome locus (*FAM9B*), which may contribute to these differences through serum testosterone levels. In rat cardiomyocytes, testosterone upregulates *KCNQ1* expression with a long-term effect on QT shortening^59^. Androgen receptors are expressed in the atrial and ventricular myocardium in multiple species including humans of both sexes^60, 61^. During puberty in males, appropriate shortening of QT is driven by increasing testosterone, while the comparatively gradual development of a relatively hypogonadal state in post-pubertal males, may explain senescent increases^58, 62^. Prolonged ventricular repolarisation is also a feature of several human diseases that share androgen deficiency as a common characteristic^63–66^. In addition to testosterone, a role for other hormones in ventricular repolarization is supported by the association of variants with QT and JT at the *THRB* locus, a nuclear hormone receptor for triiodothyronine^67^.

Loci for QRS in comparison (and therefore ventricular depolarization), have greater enrichment in processes for vasculogenesis, cell growth and embryonic development (Fig. 5). These include candidate genes encoding transcription factors (or their regulators) with roles in cellular proliferation and cardiac conduction system development (*ID2, PRDM6* and *PALLD*)^68–71^. Gene-sets were also enriched in connective tissues and cell-types. *PDE1A* is an example of one of these genes. It encodes a cyclic nucleotide phosphodiesterase and regulates cardiac fibroblast activation and fibrosis formation^72^. Myocardial fibrosis impairs cardiac electrical conduction, which increases the risk for arrhythmogenesis and is an important pathophysiological process in ventricular remodelling and cardiomyopathies^39, 73^.

In this study, we observed predominantly discordant directions of effect comparing overlapping QRS and JT loci. Despite the strong phenotypic and genetic correlation between QT and JT, the genetic contribution to the QT interval represents the combined effects of variants associated with QRS and JT. Therefore, overlap and shared biology is also observed across QT and QRS loci, along with a weak positive genetic correlation. These findings could inform drug development for arrhythmia, as genes or their encoded proteins could be targeted for their specific effects on either ventricular depolarization or repolarization.

Inherited arrhythmic syndromes highlight the importance of rare variation on ECG traits and arrhythmic risk. However, our understanding of the relationship with common variation in the general population has previously been limited. A recent study suggests the phenotypic effects of rare variants on QT are modulated by polygenic variation in the general population^74^. Our gene-based meta-analyses identified a burden of rare coding variants associated with QT, JT and QRS, in genes typically linked with inherited channelopathies (*KCNQ1*, *KCNH2*) and cardiomyopathies (*MYH7*, *TNNI3K*). Therefore, our findings support a continuum between the genetic architectures of polygenic traits and disorders that are classically considered monogenic, and highlight the utility of employing a rare variant gene-based approach in large, unselected populations.

In PRS analyses, we observed decreased risk of AF with increasing QT and QRS PRSs. This is an opposite direction of effect compared with epidemiological studies using directly measured ECG intervals where an increase in QRS or QT was associated with an increased risk of AF^75–77^. However, this relationship may be J-shaped as reported in a large study of over 280K individuals, and an increased risk of AF is also observed in patients with short QT syndrome compared to the general population^78, 79^. In addition, class-III anti-arrhythmics, used for the chemical cardioversion of AF and maintenance of sinus rhythm, inhibit hERG K+ currents that both increase the atrial refractory period (thereby contributing to a protective effect) and prolong the QT interval^80, 81^. However, our findings along with the association with conduction disease, may also reflect different biological information captured in the variance explained by the PRS, compared to the directly measured ECG trait; the latter being susceptible to modification by other factors such as coronary artery disease.

In summary, by analyzing the largest available sample size to date, we have substantially advanced the delineation of shared and distinct mechanisms influencing ventricular depolarization and repolarization. This work will inform functional follow-up and the prioritization of potential therapeutic targets for arrhythmia.

## Online methods

### Study cohorts

A total of 35 studies (involving 53 ancestry-specific sub-studies), including members of the Cohorts for Heart and Aging Research in Genomic Epidemiology (CHARGE) consortium^82^ contributed to this study (Supplementary Table 1). A maximum total of 252,977 individuals of European (84%), Hispanic (7.7%), African (6.7%), South and South-East Asian (<1%) ancestries were included. All participating institutions approved this project and informed consent was obtained for all individuals at the study level. Cohorts were predominantly population- or community-based with a small number of studies ascertained on a specific case status. Affymetrix or Illumina arrays were typically used for genotyping. Study-specific genotype quality control filters prior to imputation, including call rate, Hardy-Weinberg equilibrium (HWE) *P*-value, and minor allele frequency, are provided in Supplementary Table 2. All GWAS summary data utilized NCBI build 37. Most studies imputed ungenotyped rare and common variants using 1000G reference panels (40/53 sub-studies); the remainder used the Haplotype Reference Consortium (HRC) panel (r1.1 2016), (Supplementary Table 2)^83, 84^.

Fourteen studies (including 31 sub-studies), imputed genotype data for X chromosome analysis. The pooled multi-ancestry sample size for X chromosome analyses was 86,600 (75,607 European, 7,040 Hispanic, 1,943 African, 709 South Asian, 590 South East Asian and 711 mixed) for females and 60,343 individuals (52,070 European, 5,182 Hispanic, 1,518 African, 806 South Asian, 479 South East Asian and 379 mixed) for males.

### Cohort-level single variant association analyses

Single variant genome wide association studies were performed by each participating cohort for QT, JT and QRS. For each trait of interest, additive genetic models were implemented for two phenotypes: 1) the raw phenotype (on the millisecond (ms) scale)) and 2) the rank-based inverse normal transformation of each phenotype (on the standard deviation scale due to non-normal distributions of these traits). Per study summary statistics for each ECG measure and covariate are provided in Supplementary Table 3. Individuals were excluded at the study level for: prevalent myocardial infarction or heart failure, pregnancy at the time of recruitment, implantation of a pacemaker or implantable cardiac defibrillator, QRS duration >120ms, or right or left bundle branch block or atrial fibrillation on ECG. Additionally, if the data were available, individuals using digitalis, class I or III anti-arrhythmics or QT prolonging medication were excluded. These exclusions were chosen to reduce the risk of confounding in our analyses of ECG parameters. Cohorts including related individuals used appropriate software to account for this e.g BOLT linear mixed model software (BOLT-LMM)^85^ (which fits a linear mixed model on hard-called genotyped single nucleotide polymorphisms (SNPs)) or other software incorporating a kinship matrix or pedigree^86–88^. An imputation quality cut-off of Rsq >0.3 (or similar in IMPUTE) was applied in all cohorts to ensure high quality variants were included in the meta-analysis.

Covariates were chosen for their known association with each ECG measure and included age (years), sex (except in sex-stratified X chromosome analyses), RR interval (ms), height and body-mass index (BMI, kg/m^2^). Genetic principal components (PCs) were included to account for cryptic population stratification except in cohorts with pedigree data available or when analyses were performed using linear mixed models. Cohorts comprised of multiple ancestries performed separate analyses for each ancestry to control for underlying population stratification. Additional cohort-specific covariates were included when deemed appropriate locally, for example recruitment site or genotyping array. Autosome and X chromosome analyses were performed separately. For X chromosome analyses, male genotypes were coded as 0 or 2 and sex-stratified analyses were performed to account for random X chromosome inactivation in females. Pseudoautosomal regions were excluded from the analyses due to the high risk of genotyping errors in these regions.

### Cohort level generation of covariance matrices for gene-based testing

To test for associations due to a burden of rare (MAF≤ 0.01) variants with functional consequences within protein-coding genes, additional analyses were performed by participating cohorts using Rare variant test (Rvtests) (version 2.0.6 or later)^89^. Rvtests generates summary score statistics per variant using a separate matrix file containing the covariances between markers in a specified sliding window. To avoid associations driven by extreme outliers, these analyses were performed using only rank-based inverse normal transformed values for QT, JT and QRS. To reduce computational demand, only variants with a MAF< 1% and Rsq >0.3 were included and LD windows of 500kb were specified for construction of the genotype-covariance matrices. Rvtests uses a genomic kinship matrix to account for relatedness in each cohort and PCs were again included to adjust for residual population stratification. Analyses were only performed using individuals of European ancestry due to the potential of population differences in allele frequencies in rare variants. Only autosomes were included in these analyses.

### Central quality control of study-level data

Quality control of all study-level GWAS summary statistics was performed centrally using the EasyQC R package (version 9.2)^90^. In brief, variants were aligned to either the 1000G or HRC reference panel and allele frequency plots for each study were compared against the reference. Quantile - quantile (QQ) and *P*-value – Z-score statistics (P-Z) plots were visually inspected. Variants with invalid beta estimates, standard errors or *P*-values were removed. Per-study summary statistics were generated including standard error and beta estimate ranges. Genomic-control inflation factors (lambdas) were calculated to identify systematic inflation of test statistics, which can result from a variety of factors, including population stratification, and lead to a large number of true positive findings^91^.

### GWAS Meta-Analysis

The meta-analysis workflow is summarized in Figure 2. The primary analyses were pre-specified to be the multi-ancestry rank-based inverse normal transformed meta-analysis for each ECG trait, to avoid unstable normal approximation test statistics for low-frequency variants or outlier trait values. Ancestry-specific secondary meta-analyses were also performed for European, African and Hispanic ancestries. Due to the lack of suitablysized replication datasets, we undertook a one-stage, single-discovery design. Additionally, to enable estimation of clinically recognizable effect sizes (inverse normal transformation produces results on a standard deviation scale), a meta-analysis for each trait and ancestry was also performed using the raw phenotype on the millisecond (ms) scale. All meta-analyses were conducted using an inverse variance-weighted, fixed effects model using METAL (version released 2011-03-25) and performed independently across two sites and checked for consistency^92^. Genomic control was applied during meta-analysis to studies in which the inflation factor (λ) was > 1.0. Summary genome-wide association (“Manhattan”) plots, QQ plots and lambdas for the entire meta-analysis were produced for each trait. For all subsequent analyses, only variants present in >50% of the total sample size of the meta-analysis were included. Genome-wide significance (GWS) was defined as *P*≤ 5 x 10^-08^. The 1000 Genome reference panel was used for calculating correlations between variants in downstream analyses, including all individuals for multi-ancestry summary statistics and individuals from relevant populations for European, African and Hispanic meta-analyses. Where pre-computed LD scores were required, or correlations calculated within tools that did not permit modification of individuals included in the reference panel, the European ancestry meta-analysis was used in place of the multi-ancestry recognising a substantial proportion of the multi-ancestry meta-analysis included individuals of European descent. When this is the case, it is explicitly stated in the methodology and results.

### Definition of known and novel loci

To identify novel associations in our results, we first defined boundaries for previously published loci (Supplementary Table 4) using the following process in PLINK(v1.9)^93^. Reported genome-wide significant (*P*≤ 5 x 10^-08^) lead variants from each published GWAS were extracted and correlations calculated using the 1000 Genome phase 3 reference panel (Nov 2014)^83^ in a 4Mb region centred on each variant. The locus start was defined as minus 50kb from the most upstream variant that was in r^2^ > 0.1 with the lead variant and the locus end as plus 50kb from the most downstream variant which was in r^2^ > 0.1. Overlapping boundaries were subsequently merged. This window was declared the genomic boundary for the locus or a minimum physical distance window of ±500kb around the reported variant – whichever was larger. For the purpose of these analyses, as QT and JT phenotypes are highly correlated phenotypes (Figure 1), previously reported JT and QT loci were pooled. A separate list was compiled for previously described QRS duration loci. Novel loci in our meta-analyses were identified by applying the same approach to variants meeting the GWS threshold in our results. Variants within loci boundaries not overlapping with known loci were declared novel associations. Subsequently, to evaluate for evidence of heterogeneity at each locus, a forest plot was produced for the lead variant using the R-package Metaviz (version 0.3.0) and manually inspected along with the I^2^ heterogeneity index for that variant. Finally, Locus-Zoom plots were generated for each locus to visually inspect correlations (r^2^) between lead variants and surrounding markers and their associated *P*-values^94^.

### Conditional and heritability analyses

We sought to determine whether any variants at a given locus were conditionally independent (i.e. independent signals of association). Conditional analyses were performed for loci that achieved genome-wide significance in the European-ancestry analysis using 52,230 individuals of European ancestry from UK Biobank using Genome-wide Complex Trait Analysis (GCTA, v1.26.0)^18^. Related pairs up to the 2^nd^-degree (kinship coefficient< 0.0884) were excluded. Due to insufficient reference sample size and an inability to effectively reproduce the ancestral mix, conditional analyses were not performed for other ancestries or the multi-ancestry meta-analyses. Stringent thresholds of *P*_Joint_ < 5×10^-08^ and minimal correlation (r^2^< 0.1) with the lead variant were used to declare a variant “conditionally independent”. Using the same dataset, heritability estimates for each trait in European samples were obtained using BOLT-Restricted Maximum Likelihood (REML, v2.3.2), which applies variance components analysis using modelled directly genotyped SNPs to calculate SNP-based heritability^85^. The percent variance explained (PVE) by lead and conditionally independent variants was calculated as^95^;

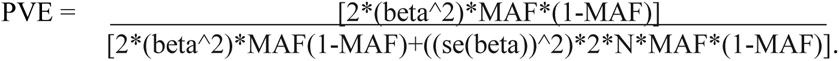

The total PVE by all lead and conditionally independent variants was calculated as the sum of each variant’s PVE. The heritability explained was the total PVE divided by the heritability of the trait.

### LD score regression

To calculate the genetic correlation between ECG traits, LD score regression was performed using LD SCore (LDSC) software (v1.0.1).^24^. European meta-analysis summary statistics were filtered to include only variants present in the International HapMap Project (∼1.1 million variants), along with pre-computed LD scores using the 1000G reference panel provided by LDSC^24^. LDSC uses the LD scores as regression weights and subsequently calculates the genetic correlation using intersecting SNPs from each meta-analysis^25^.

### Gene-based testing meta-analysis

Gene-based meta-analysis was performed using the R package rareMETALs (v.7.1)^96^. Analyses were restricted to up to 192,780 individuals of European descent (from 37 studies) due to potential differences in the allele frequency of rare (MAF≤ 0.01) variants between populations. QC of study-level data was performed as previously described. Variants from all studies, however, were subsequently filtered to only include those predicted by VEP (Ensembl release 99) to have high or moderate impact (and thus be protein-altering). Score and covariance files used as input for gene-based meta-analyses in rareMETALS were generated using Rvtests as described above^89^. Gene-based meta-analysis was subsequently performed for inverse-normal transformed QT (N=192,780), JT (N=192,501) and QRS (N=192,495) using Sequence Kernel Association Testing (SKAT)^27^, which considers the joint effects of multiple variants on the phenotype, while taking into account the effect size and direction of effect of each variant. Power under SKAT is maximal for genetic variation in a gene that causally increases and decreases a quantitative trait, but is less powered to detect genetic effects that all influence a quantitative trait in one direction. Gene-based meta-analysis was conducted for 18,751 genes that had more than one rare (MAF ≤ 0.01) variant annotated as high or moderate impact. A gene-based test was considered significant if *P*< 2.5 x 10^-06^ (Bonferroni correction for ∼20,000 tested genes).

To follow-up on gene associations passing Bonferroni correction, additional conditional analyses were performed for each of the three traits. First, to confirm that the gene-based association is not solely driven by one rare variant, conditional gene-based meta-analyses were repeated while conditioning on the most significant variant in the gene. Genes were considered significant if *P_conditional_*< 0.05/number of genes tested for the trait (13 genes for QT, 16 genes for JT and 3 genes for QRS), upon gene-based analysis conditioned on the most significant variant. Second, conditional analyses were restricted to 76,202 individuals from the UK Biobank to ensure a common set of variants were able to be examined. To confirm that the gene-based association was not solely driven by flanking low-frequency and common (MAF >0.01) QT/JT/QRS variants at the locus in which the gene is located regardless of their annotation with VEP, (P< 5 x 10^-08^ and r^2^< 0.1 or ±500kb from the lead variant in the respective GWAS meta-analysis) analyses were repeated while conditioning on all independent variants at the locus. Furthermore, additional conditional analyses were conducted for associated genes that reside in the same locus. This was done to ensure that these are independent gene-associations in the same locus and not attributable to rare variants in the other gene in the locus.

### Biological annotation of GWAS loci

#### Identification of variant consequences

To identify variants with potential functional consequences, we annotated lead and conditionally independent variants, and their proxies (r^2^ >0.8), using VEP (Ensembl release 99)^26^ to extract information on the impact of a variant on a transcript or protein including their deleteriousness scores using the Sorting Intolerant From Tolerant algorithm (SIFT, version 5.2.2)^97^ and PolyPhen-2 (Version 2.2.2^98^). Additionally, Combined Annotation Dependent Depletion (CADD)^99^ and RegulomeDB^100^ rank scores for these variants were extracted. CADD scores correlate with pathogenicity of both coding and non-coding variants and rate a variant according to its deleteriousness within the genome^99^. RegulomeDB annotates variants with known and predicted regulatory elements in intergenic regions including regions of DNase hypersensitivity, transcription factors binding sites, and promotor regions utilising publicly available datasets^100^.

#### Association with tissue-specific gene expression

To evaluate correlations between GWAS variants and tissue-specific gene expression, data from the Genotype-Tissue Expression project (GTEx, v8)^33, 101–103^ were extracted for tissues relevant to cardiac electrophysiology including cardiac, vascular (coronary artery and aorta) and brain (as autonomic regulation influence ECG traits). First, lead and conditionally independent variants and their proxies (r^2^ >0.8), were checked to determine whether they overlapped with the lead variant at an expression quantitative trait locus (eQTL) for each tissue. Additionally, to determine whether the same variant may be causal in both our GWAS meta-analysis and the original eQTL study, colocalization analyses were performed using the R package COLOC, which uses Bayesian methods to determine the correlation between variants from the two datasets^104^. A posterior probability of >75% was used to determine significance.

#### Tissue- and cell-type specific regulatory elements

To identify tissue-specific enrichment of variants in DNaseI hypersensitivity sites, we used GWAS Analysis of Regulatory and Functional Information Enrichment with LD correction (GARFIELD, v2)^37^. GARFIELD performs greedy pruning of GWAS SNPs (r^2^ > 0.1) and annotates them based on overlapping functional information to assess the enrichment of association signals with features extracted from the ENCODE, GENCODE and Roadmap Epigenomics projects^37^. Odds ratios are quantified and assessed using a generalised linear model framework while matching for MAF, distance to the nearest transcription start site, and number of proxies (r^2^ >0.8).

We sought to identify potential target genes of regulatory variants using long-range chromatin interaction (Hi-C) data analyzed using FUMA GWAS (Functional Mapping and Annotation of Genome-Wide Association Studies) software (v1.3.6)^105^. Within FUMA, pre-processed significant loops computed by Fit-Hi-C pipelines filtered at an FDR < 0.05 and overlap with lead and conditionally independent variants and their proxies (r^2^ >0.8) was identified^34^. Additionally, we utilised recently published promoter capture Hi-C data which uses loops called from Knight-Ruiz normalised 5kb, 10kb and 25kb resolution data^35^. Promoter interactions in left and right ventricular tissue for potential regulatory variants were extracted and variants with the highest regulatory potential were determined using a RegulomeDB score cut-off of ≤3b^100^.

To identify cardiac cell-type specific functional effects of non-coding variants, we integrated GWAS variants with cell-type chromatin marks from single nucleus Assay for Transposase-Accessible Chromatin using sequencing (snATAC-seq) data^36^. These data contain open/accessible chromatin information for nine cell-types obtained from the heart, including atrial and ventricular cardiomyocyte, smooth muscle, endothelial, adipocyte, macrophage, fibroblast, lymphocyte and nervous cells. Haplotype blocks were created for each lead and conditionally independent variant including variants with r^2^ >0.1 within a 2Mb radius. Subsequently, using a SNP enrichment method, CHEERS (Chromatin Element Enrichment Ranking by Specificity)^106^, the peaks with the lowest 10^th^ percentile of total read counts from the snATAC-seq data were removed, the peak counts were subsequently quantile normalised, and the Euclidean distance calculated^106^. A one-sided *P*-value for enrichment of variants in estimated haplotype blocks within cell-type specific ATAC-seq peaks was calculated and a Bonferroni-corrected threshold used to declare significance (0.05/number of cell-types).

#### Candidate gene prioritisation and pathway enrichment

To prioritise candidate genes at each locus, we used Data-driven Expression-Prioritized Integration for Complex Traits (DEPICT, v3). DEPICT prioritizes the most likely causal genes at associated loci according to common functional pathways using reconstituted gene-sets containing a membership probability for each gene in the genome^38^. Additionally, it highlights enriched pathways and tissues/cell types where genes from associated loci are highly expressed. DEPICT uses a clumping method (r^2^ = 0.1, window size = 250kb, *P*= 5 x 10^-08^) to identify uncorrelated variants from each meta-analysis using 1000G reference data after excluding the major histocompatibility complex region on chromosome 6. Gene-set enrichment analysis was conducted based on 14,461 predefined reconstituted gene sets from various databases and data types, including Gene Ontology (GO), Kyoto Encyclopedia of Genes and Genomes (KEGG), REACTOME, phenotypic gene sets derived from the Mouse genetics initiative, and molecular pathways derived from protein–protein interactions. Finally, tissue and cell type enrichment analyses were performed based on expression information in any of the 209 Medical Subject Heading (MeSH) annotations for the 37,427 human Affymetrix HGU133a2.0 platform microarray probes. Cytoscape (version 3.8.2, https://cytoscape.org/)107 was used to visualise significantly enriched (FDR< 0.01) DEPICT GO biological processes for each ECG trait. Processes were connected by overlap of significantly enriched genes (minimum number of 25% of member genes). Orphan pathways or those with less than three edges were excluded. Each module was labelled with a common theme to represent the group of biological processes.

DEPICT requires at least ten genome-wide significant loci to be able to perform the analysis. Therefore, for African and Hispanic ancestries, candidate genes were identified using g:Profiler^108^, a functional enrichment tool for the annotation of a list of genes. It also enables mapping of variants to gene names, where they overlap with at least one protein coding Ensembl gene with annotation of predicted variant effects.

In addition to DEPICT gene prioritisation results, the list for each locus was supplemented with candidate genes highlighted by these bioinformatic analyses. A literature review was performed which also included a look up of genes using the Online Mendelian Inheritance in Man (OMIM) and International Mouse Phenotyping Consortium (IMP) databases.

#### Druggability analyses

To identify potential novel drug targets from our GWAS findings, we interrogated a previously published druggable gene set database developed by Finan et al. which includes detailed information on methods used to assemble and annotate the dataset^40^. In brief, this reference set contains predominantly protein-coding genes (as annotated from the Ensembl v.73). Genes were subsequently assembled into three tiers: Tier 1 includes targets of approved drugs and drugs in clinical development including targets of small molecule and biotherapeutic drugs. Tier 2 incorporates proteins closely related to drug targets or associated with drug-like compounds. Genes where one or more Ensembl peptide sequence shared ≥50% identity (over ≥75% of the sequence) with an approved drug target were included. Tier 3 incorporated extracellular proteins and members of key drug-target families (including G protein-coupled receptors, kinases, ion channels, nuclear hormone receptors, and phosphodiesterases). Using the list of “most likely candidate genes” at unreported GWAS loci and previously reported candidate genes for QT, JT and QRS, a look up was performed in the database. Genes which are existing drug targets for anti-arrhythmic drugs (annotated using KEGG drug (https://www.genome.jp/) were excluded. As genes which were significant findings in cardiac tissue-specific eQTL co-localization and Hi-C analyses may be favoured for prioritization, a look up was performed in our data to highlight these.

### Association between genetically determined QT, JT and QRS and relevant cardiovascular diseases

To determine relationships of genetically pre-determined QT JT QRS with cardiovascular diseases PRSs were constructed using the lead variants from the European-ancestry meta-analyses and tested for association with prevalent cases of atrial fibrillation, stroke, coronary artery disease, heart failure, non-ischaemic cardiomyopathy, conduction disease (“AVB or PPM implantation” and “fascicular block/BBB”) and ventricular arrhythmia in the UK Biobank. Secondary analyses were also performed to test for association with subgroups including myocardial infarction and stroke. Outcomes were identified using self-reported data, operation codes, ICD-9/ICD-10 codes from hospital episodes statistics and mortality registry (Supplementary note 2). Analyses were performed in individuals of European-ancestry without ECGs and therefore not included in the GWAS meta-analysis and related pairs up to the 2^nd^-degree (kinship coefficient <0.0884) were excluded. To take advantage of genotype probability data in BGEN format, PRSice-2^109^ was used to calculate the PRS. The PRS was calculated by summing the dosage of the ECG trait prolonging allele, weighted by the effect size from the corresponding GWAS. Associations with prevalent cases were identified using logistic regression including covariates age, sex, 10 PCs and genotype array. A Bonferroni-corrected threshold of 0.05/number of outcomes tested (0.05/7 = 7.1×10^-3^) was used to determine significant associations. P-values < 0.05 but greater than 7.1×10^-3^ were considered as suggestive associations.

To determine whether genetically determined QT, JT and QRS are associated with SCD, PRS were constructed for each trait and tested in four cohorts: the Atherosclerosis Risk in Communities study (ARIC)^110, 111^, the Cardiac Arrest Blood study (CABS)^112^, the Finnish Genetic Study for Arrhythmic Events (FinGesture) and Northern Finland Birth Cohort of 1966 (NFBC1966)^113^. The latter three are independent cohorts as they were not included in the GWAS meta-analysis. While ARIC was included in the GWAS meta-analysis, it contributes a relatively small proportion of the GWAS meta-analysis sample size and thus effects on beta-estimates would be negligible. Individual study information and case definitions used are available in Supplementary note 3. As FinGesture contains only cases, population controls were taken from NFBC1966^114^. For these analyses, only individuals of European-ancestry were included, due to low sample sizes of other ancestries. PRSs were constructed by averaging the dosage of each lead variant allele associated with prolongation of the ECG trait being tested, from the European ancestry meta-analysis, weighted by the effect size from the corresponding raw-phenotype meta-analysis. To ensure only high-quality variants were included, each study applied an imputation quality threshold (Rsq >0.8). The PRS was included in a logistic regression (Cox for ARIC) model along with covariates age (when appropriate), sex, 10 PCs and genotyping array (when appropriate). Sex-stratified analyses were performed to identify sex-specific effects. Per study summary statistics were subsequently meta-analysed using an inverse-variance weighted model with the R package ‘Meta’^115^.

## Data Availability

All data produced in the present study are available upon reasonable request to the authors

## Acknowledgements

All individual and study acknowledgements can be found in Supplementary Note 4, and funding information in Supplementary Note 5. The authors also wish to acknowledge the CHARGE infrastructure grant (HL105756).

## Author contributions

Interpreted results, writing, and editing the manuscript: W.J.Y, N.L, A.I, N.S, B.M, C.N.C, P.B.M. Conceptualization of project: N.S, C.N.C, P.B.M. Supervision of project: N.S, B.M, C.N.C, P.B.M. Contributed to GWAS analysis plan: W.J.Y, N.L, N.S, C.N.C, P.B.M. Performed meta-analyses: W.J.Y and N.L. Performed GCTA, genetic correlations, heritability, variant annotations, GTEx analyses, HiC analyses, gene-set enrichment and pathway analyses, druggability analyses, PRS analyses in UKB: W.J.Y. Performed PRS analyses in SCD cohorts: T.D, J.A.B. Contributed data for PRS SCD analyses: M.-R.J, H.V.H, R.N.M, J.J, P.F, R.L, K.P. Performed gene literature review: W.J.Y, L.F, F.A, R.S, D.R. Visualization of enrichment pathways: S.A.B. Contributed to study-specific GWAS by providing phenotype, genotype and performing data analyses: W.J.Y, A.I, T.D, L.F, J.A.B, R.N, J.-W.B, J.H, L.-P.L, L.R, M.P.C, M.B, S.W, A.R.B, T.M.B, J.P.C, D.S.E, R.F, O.H, J.L.I, H.L, H.M, A.M, M.M.-N, C.N, Y.Q, A.R, C.R, K.R, E.T.-S, S.T, S.V.D, H.R.W, J.Y. S.A, G.A, A.A, L.A, J.C.B, E.B, A.C, E.C, M.C, M.J.C, D.D, A.D.G, A.D.L, J.D, C.E, P.T.E, S.B.F, C.F, M.G, C.G, M.G, X.G, T.H, S.R.H, P.L.H, N.H.-K, A.I, R.D.J, M.J, J.J, M.K, J.A.K, T.P.L, H.J.L, L.L, A.L, S.L, P.M, M.M, T.M, M.Me, P.P.M, R.N.M, N.M, M.E.M, A.C.M, M.N, V.N, P.N, K.N, G.P, G.Pe, A.P, D.J.P, P.P.P, M.H.P, O.T.R, A.P.R, A.L.P.R, K.M.R, L.R, D.S, U.S, C.S, X.S, M.B.S, G.S, M.F.S, E.Z.S, M.S, K.S, K.T, K.D.T, A.T, S.T, A.U, U.V, H.V, M.W, L.-C.W, E.W, J.G.W, C.L.A, D.C, A.C, F.C, M.D, G.G, N.G, C.H, Y.Y, J.W.J, S.K, M.K, J.K.K, C.K, T.L, M.F.L.-C, Y.L, R.J.F.L, S.A.L, D.O.M.-K, A.P.M, J.R.O, M.S.O, M.O, S.P, C.P, A.P, B.M.P, J.I.R, B.S, P.H, C.M.D, N.V, J.F.W, D.E.A, J.R, P.D.L, N.S, P.B.M. All authors read, revised, and approved the manuscript.

## Footnotes

These authors jointly supervised this work: Nona Sotoodehnia, Borbala Mifsud, Christopher Newton-Cheh, Patricia B Munroe

## Data availability

The datasets generated from the meta-analyses, during the current study are available from the corresponding author on reasonable request.

## Code availability

Codes generated during the current study are available from the corresponding author on reasonable request.

## Supplementary Data

### Supplementary Figures

**Supplementary Figure 1:**
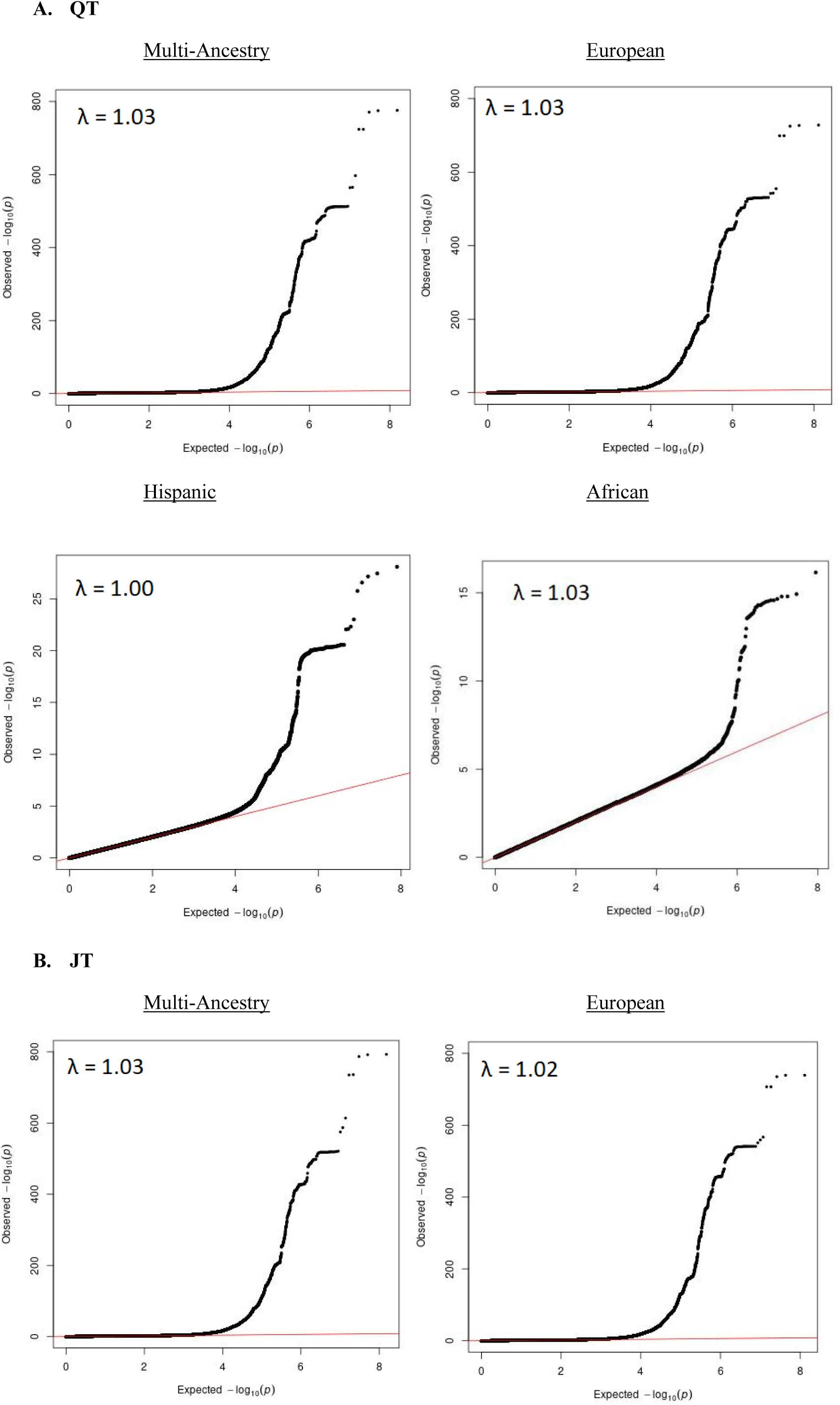
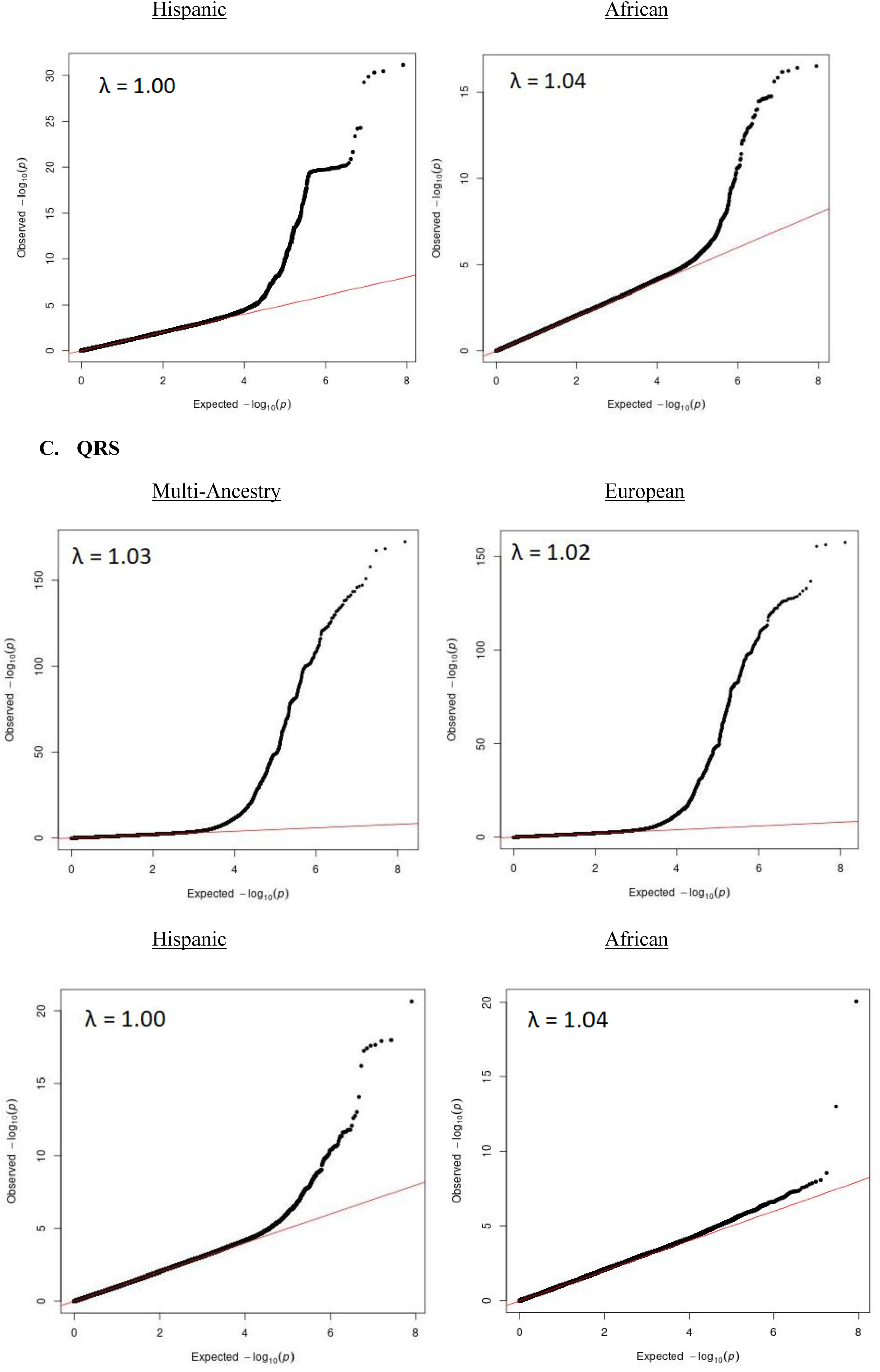
QQ plots for QT, JT and QRS meta-analysis. The primary analysis was multi-ancestry. Plots for ancestry-specific meta-analyses (secondary analyses) are also shown. λ = genomic control lambda. A) QT meta-analysis, B) JT C) QRS.

**Supplementary Figure 2:**
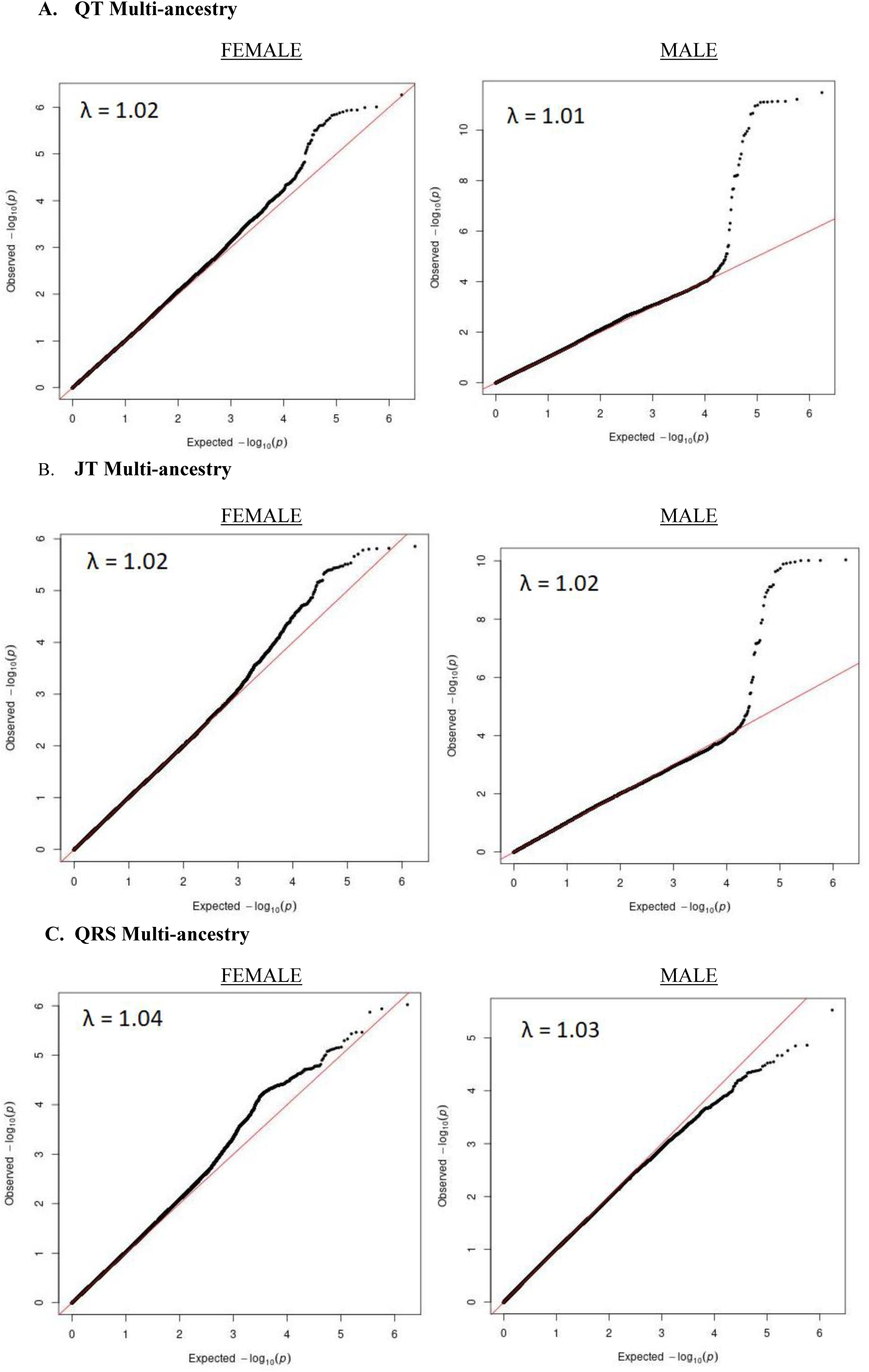
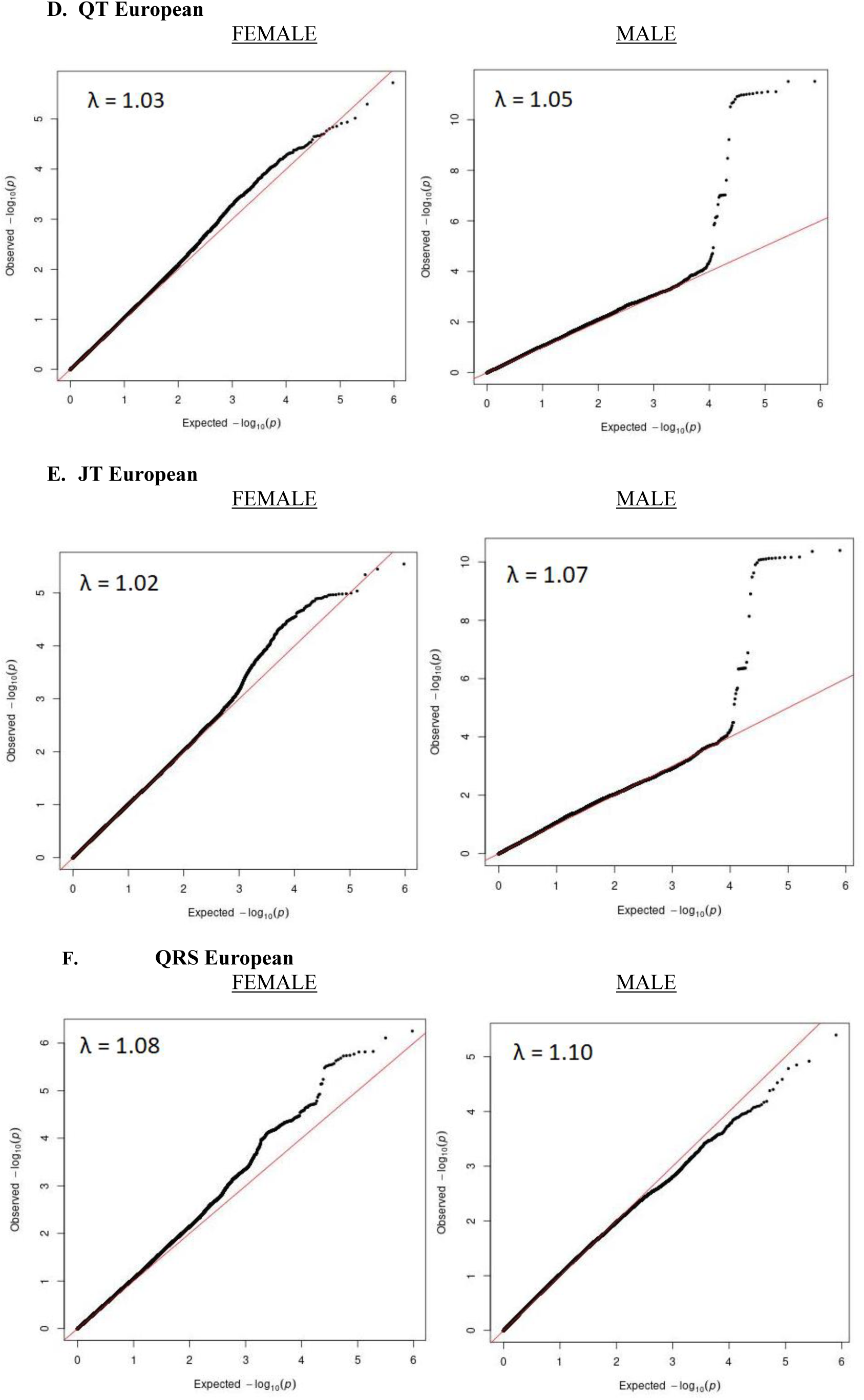
QQ plots for multi-ancestry and European X-chromosome analyses. Female and Male stratified results are in left and right panels respectively. λ = genomic control lambda imbedded in each plot (methods). A) Plots for QT multi-ancestry, B) JT multi-ancestry, c) QRS multi-ancestry D) QT European, E) JT European, F) QRS European meta-analyses.

**Supplementary Figure 3:**
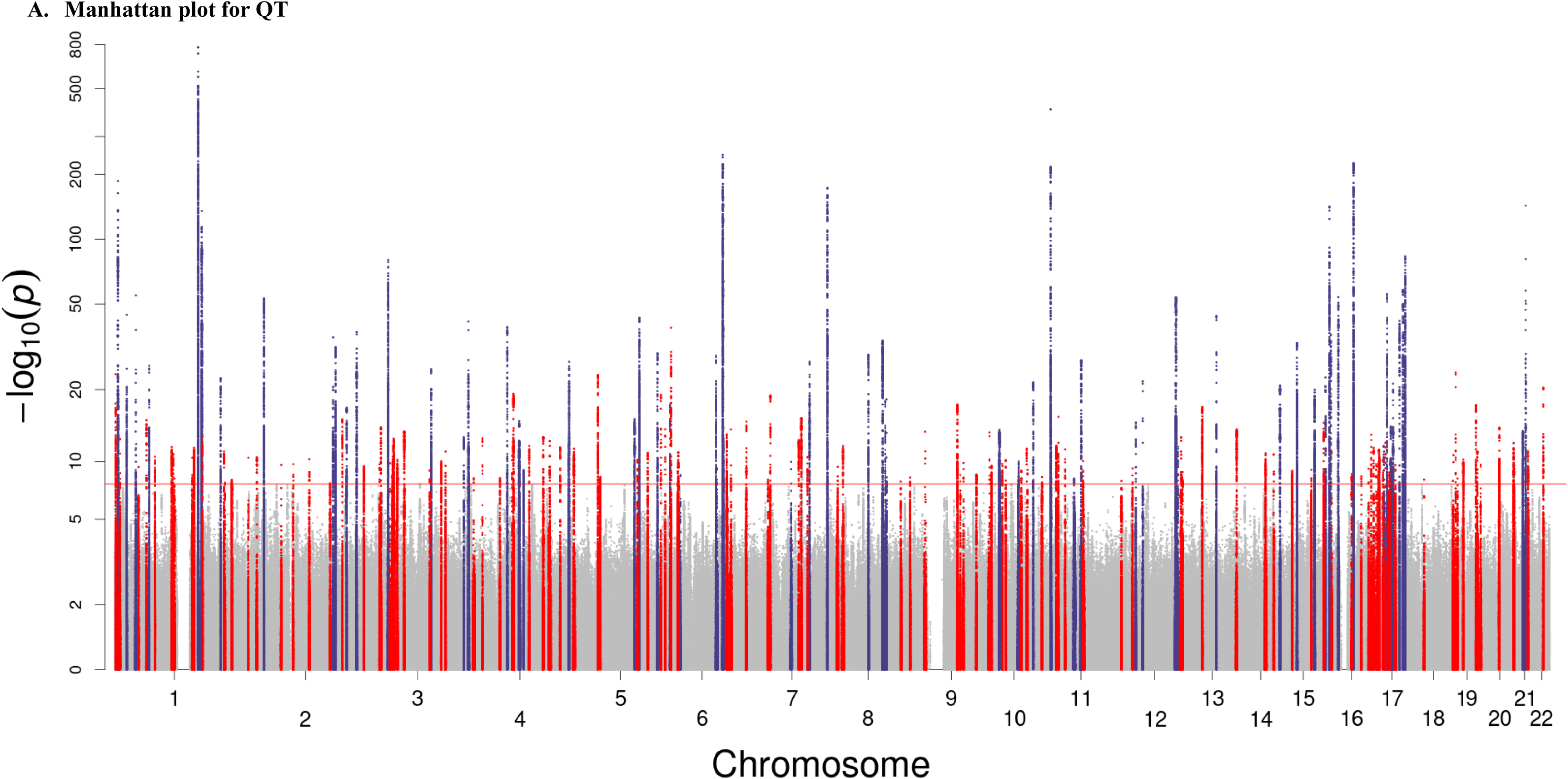
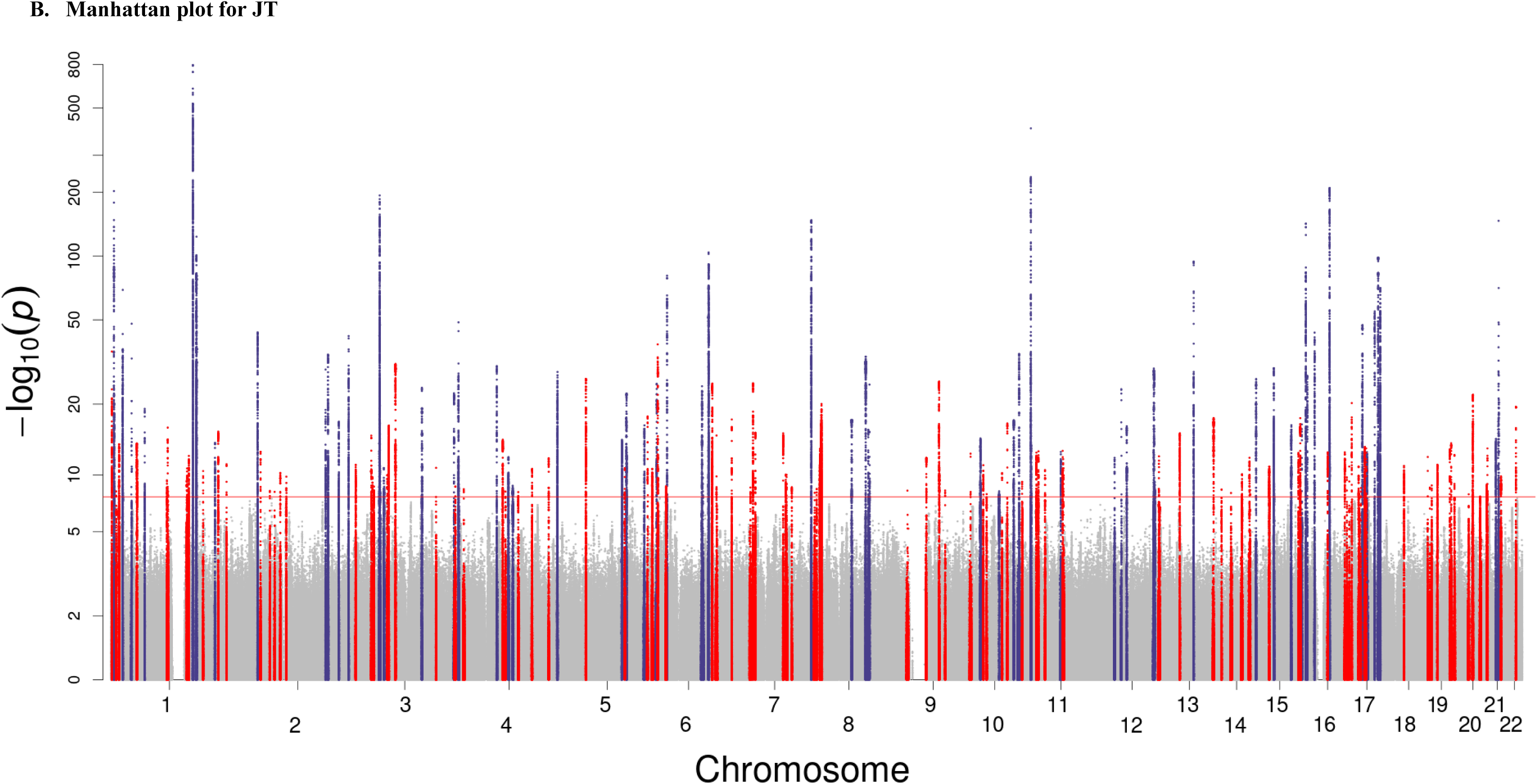
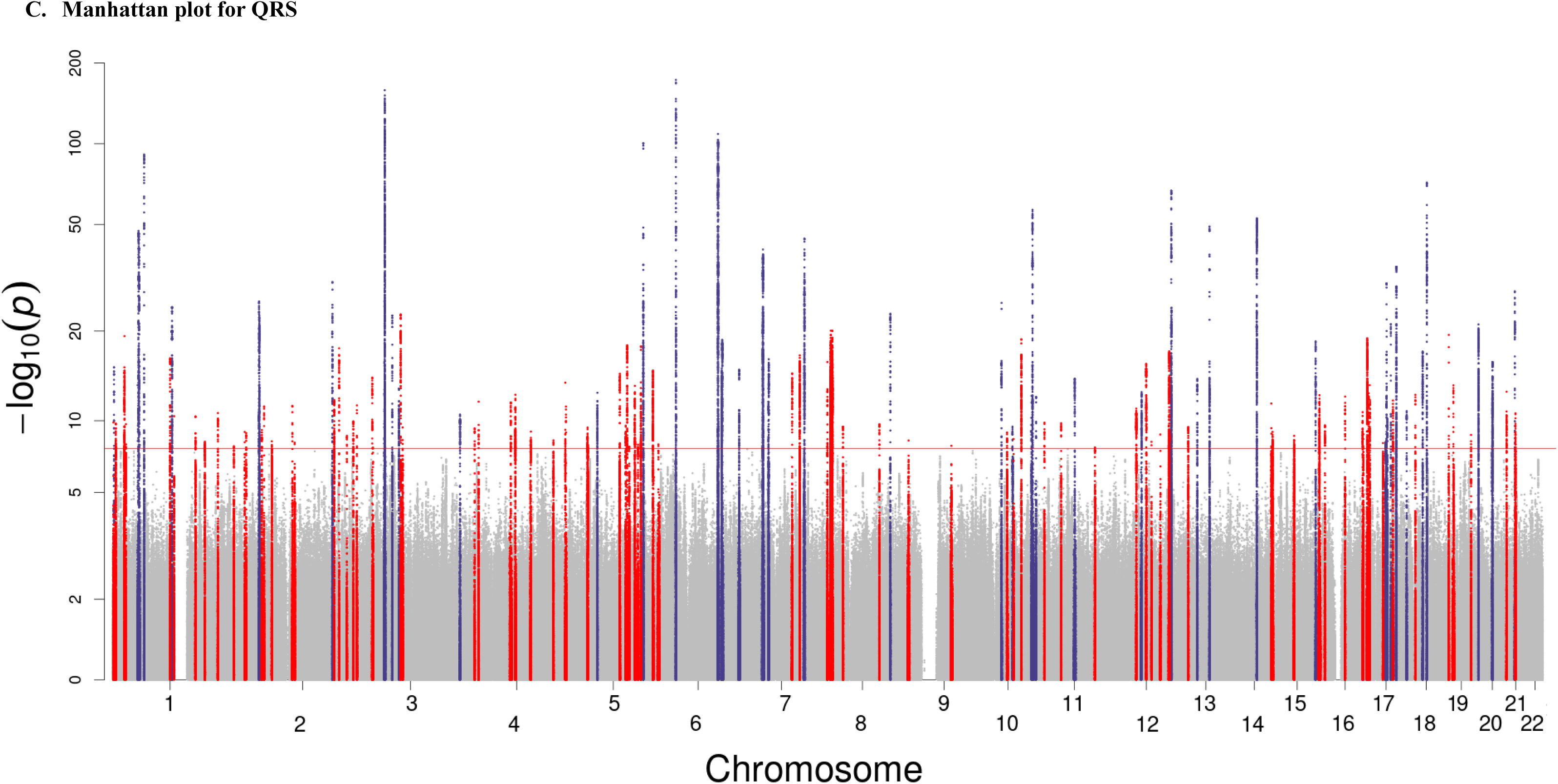
Manhattan plots for Qt, JT and QRS multi-ancestry meta-analyses. *P* values are plotted on the -log_10_ scale (Y axis). Red horizontal line = genome-wide significance (*P* < 5 × 10^−8^). Variants within the boundaries of previously unreported (N=114 for QT, N = 96 (JT), 77 (QRS)) and previously reported loci (N = 62 for QT, N = 59 (JT), 44 (QRS)) are plotted in red and dark blue colors respectively.

**Supplementary Figure 4:**
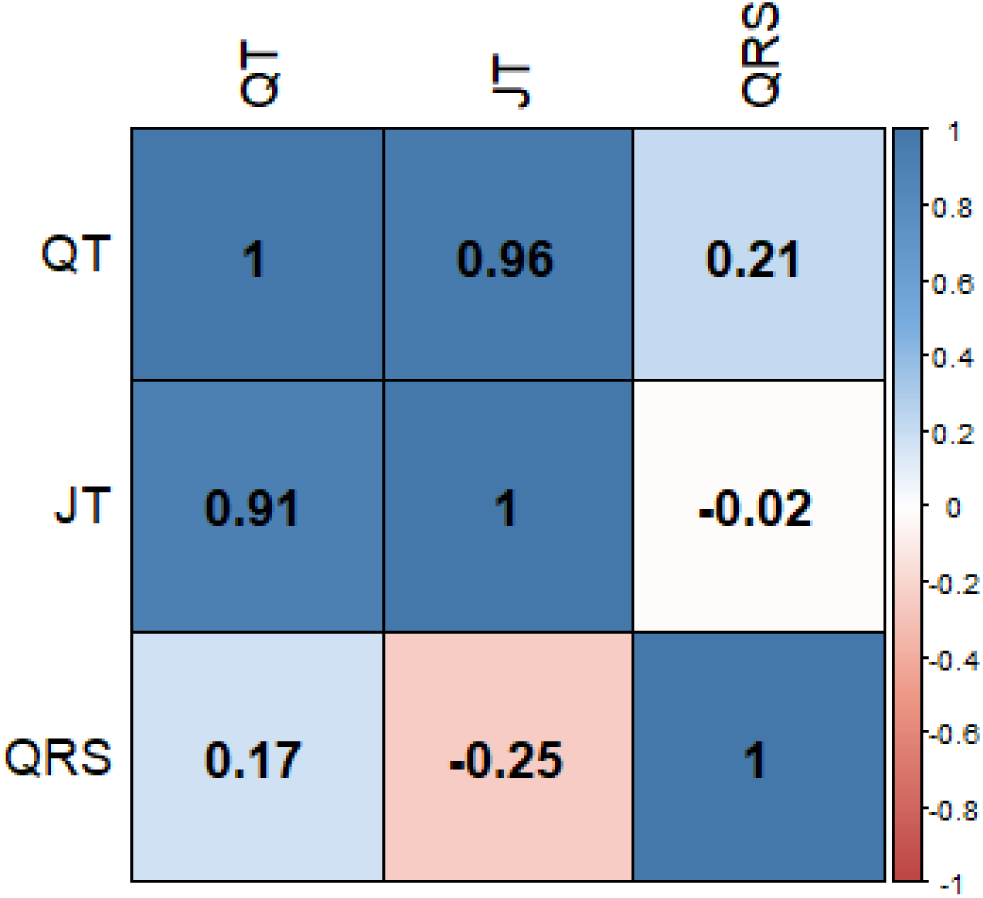
Genetic and phenotypic correlations of QT, JT and QRS. Genetic correlations (r_g_) calculated using LDSC regression and displayed in lower left triangle. Phenotypic correlations (Spearman’s rank correlation coefficients [r_s_]) were calculated in ∼51K UK Biobank individuals of European ancestry and are presented in the upper right triangle.

**Supplementary Figure 5:**
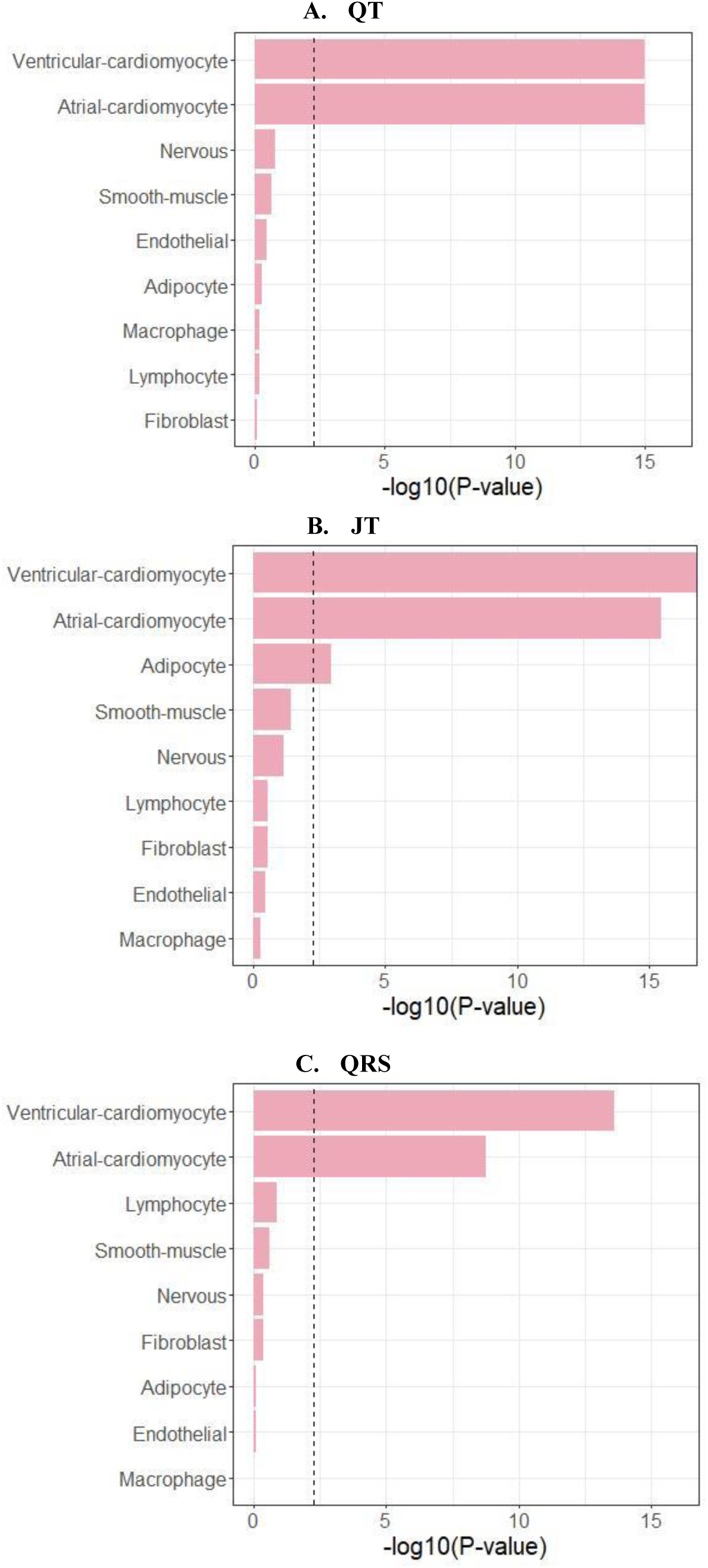
Cardiac cell-type overlap with snATAC-seq peaks for QT, JT and QRS. Results for singe nuclear ATAC seq (snATAC-seq) analyses for QT, JT and QRS multi-ancestry meta-analyses (methods). X-axis: log base 10 *P*-value for cell-type enrichment. Y-axis: Cell-type. Vertical dashed line: Bonferroni corrected significance threshold for number of cell types used.

**Supplementary Figure 6:**
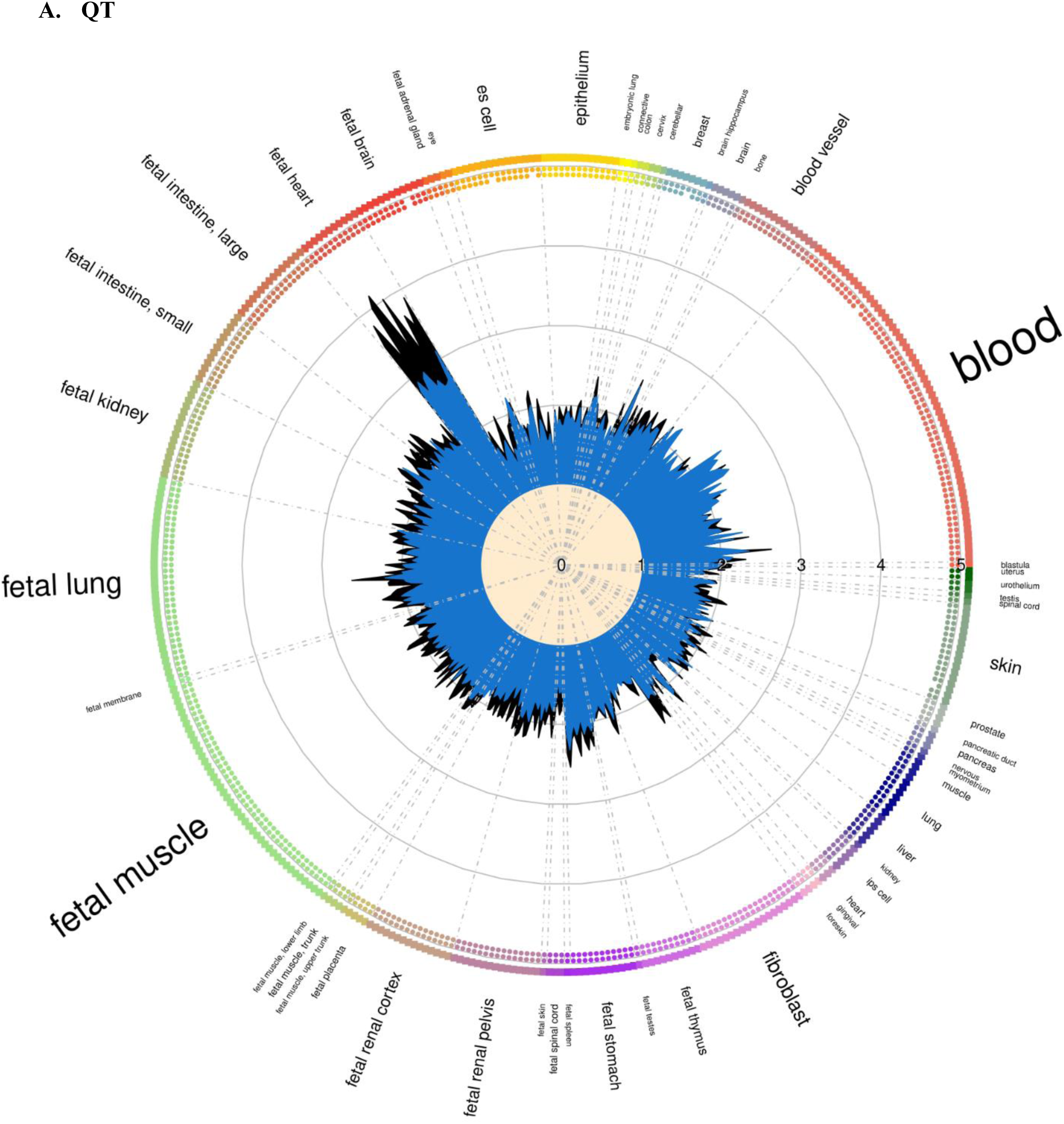
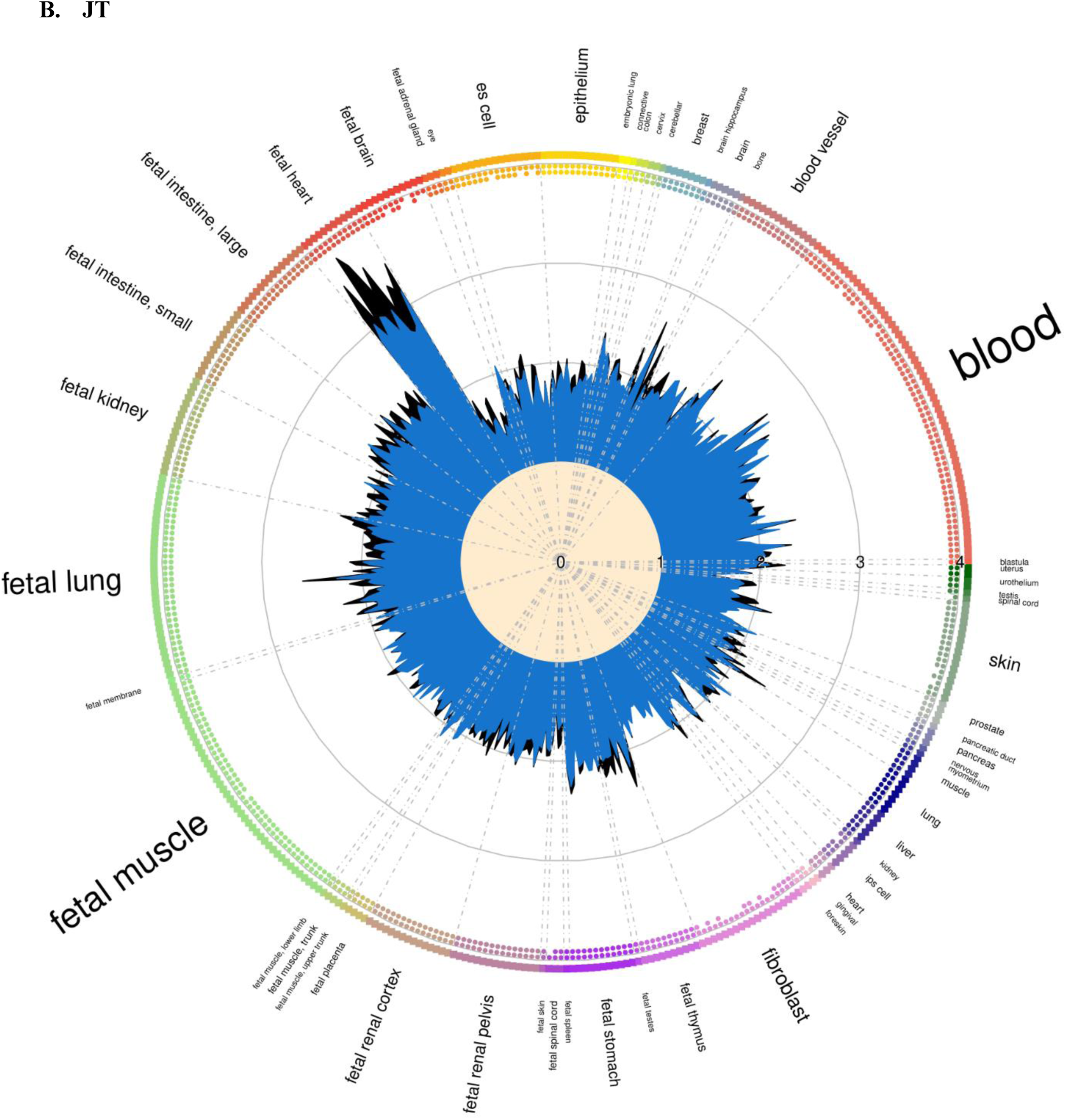
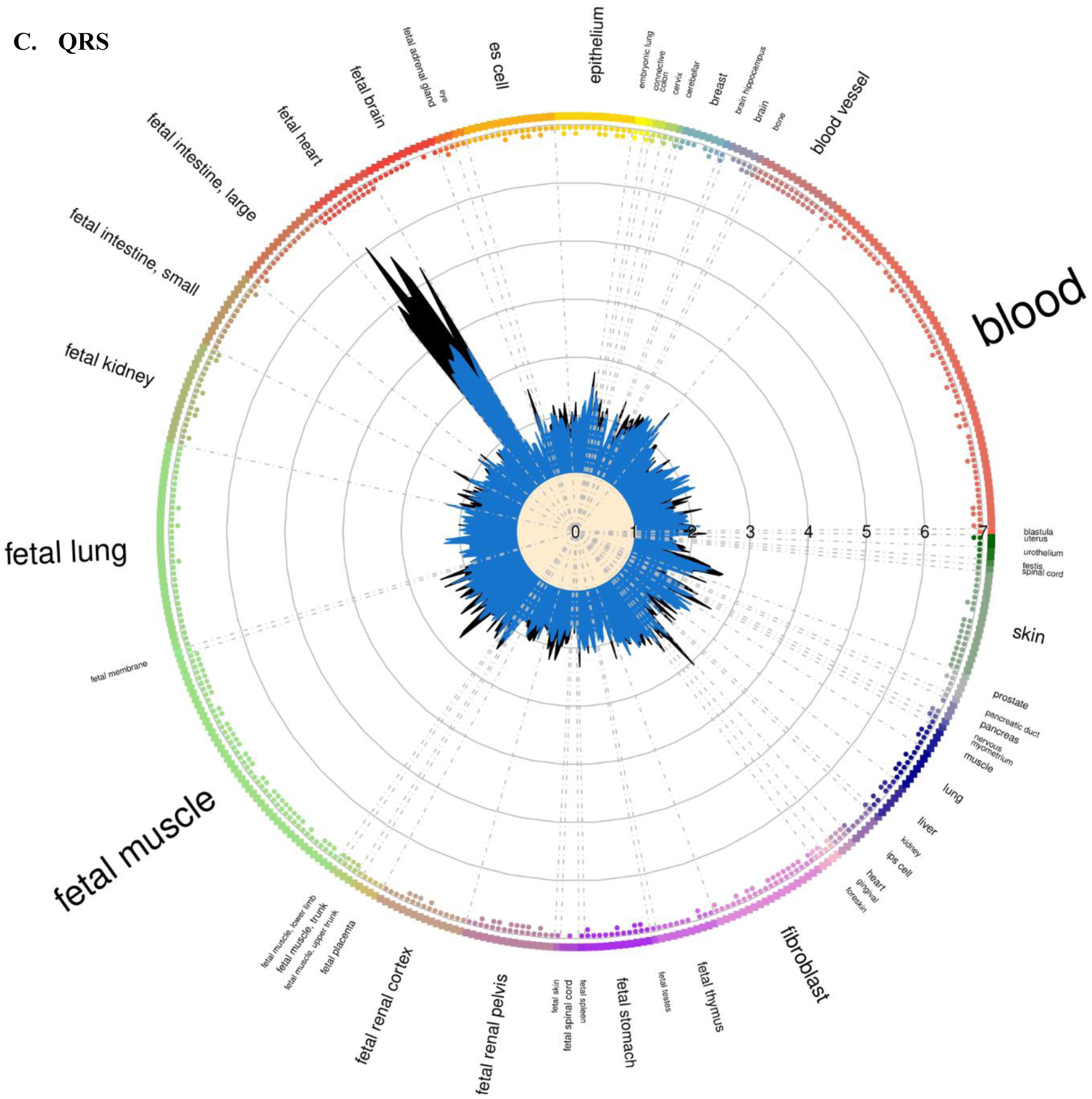
Enrichment of QT, JT and QRS variants in DNaseI Hypersensitivity sites. DNaseI hypersensitivity sites were derived from the Encyclopaedia of DNA elements (ENCODE) and Roadmap Epigenomics projects using GARFIELD (Methods). European ancestry meta-analysis results were used. Tissue font size is proportional to the number of cell types for that tissue. Radial plot shows odds ratio (OR) in each cell type at two GWAS *P*-value thresholds (<1×10^-5^ and < 5×10^-8^). Small dots on the inner side of the outer circle show if the observed enrichment is significant in direction outside to inside.

**Supplementary Figure 7:**
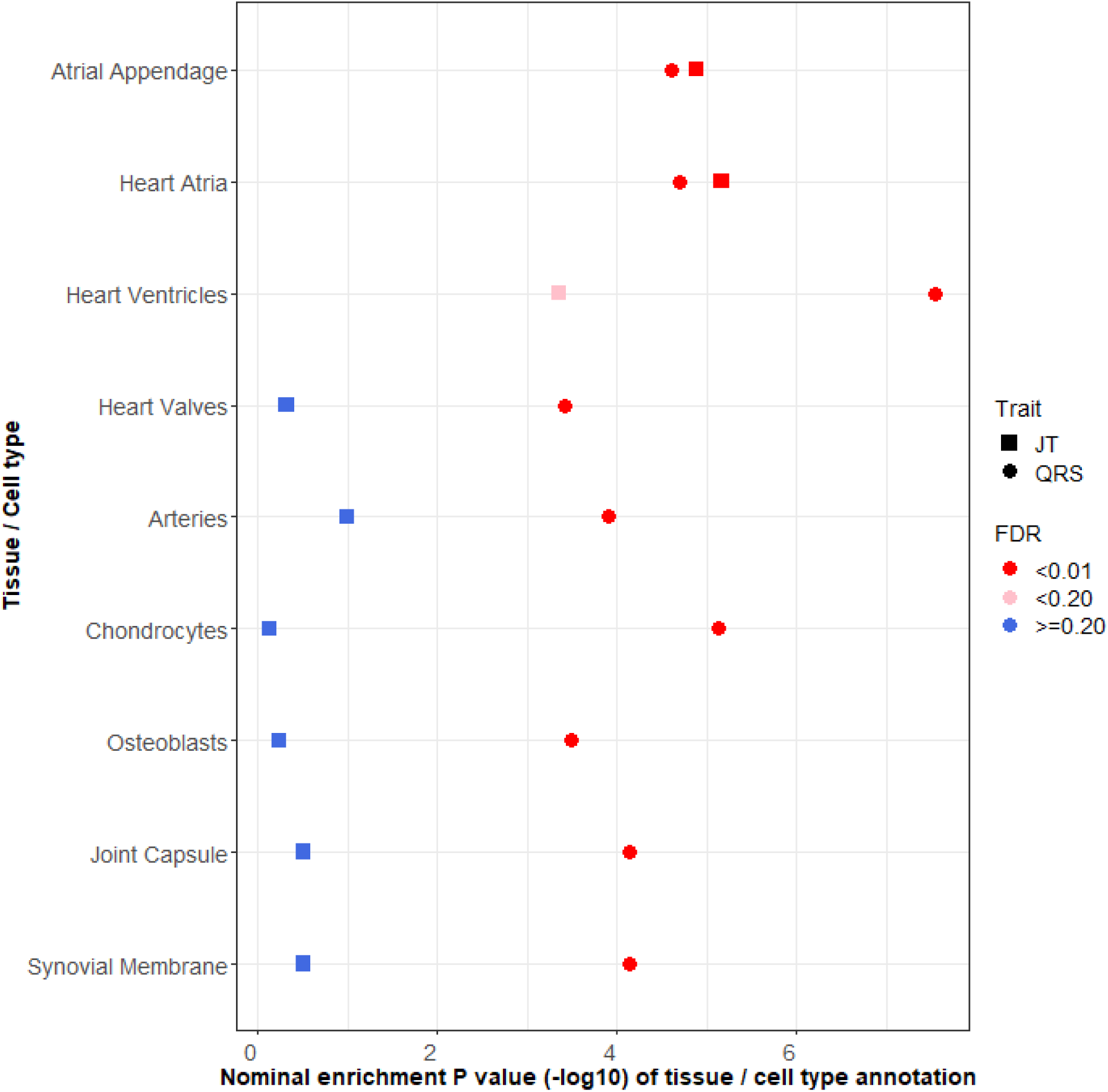
DEPICT tissue/cell type enrichment analysis for JT and QRS. X axis – Nominal P value for enrichment, Y axis – Tissue/cell types. Results indicated where significant with an FDR < 0.01 and comparisons are provided for the same tissue in the other trait. Full results with FDR < 0.01 including for QT are reported in Supplementary Table 15.

**Supplementary Figure 8:**
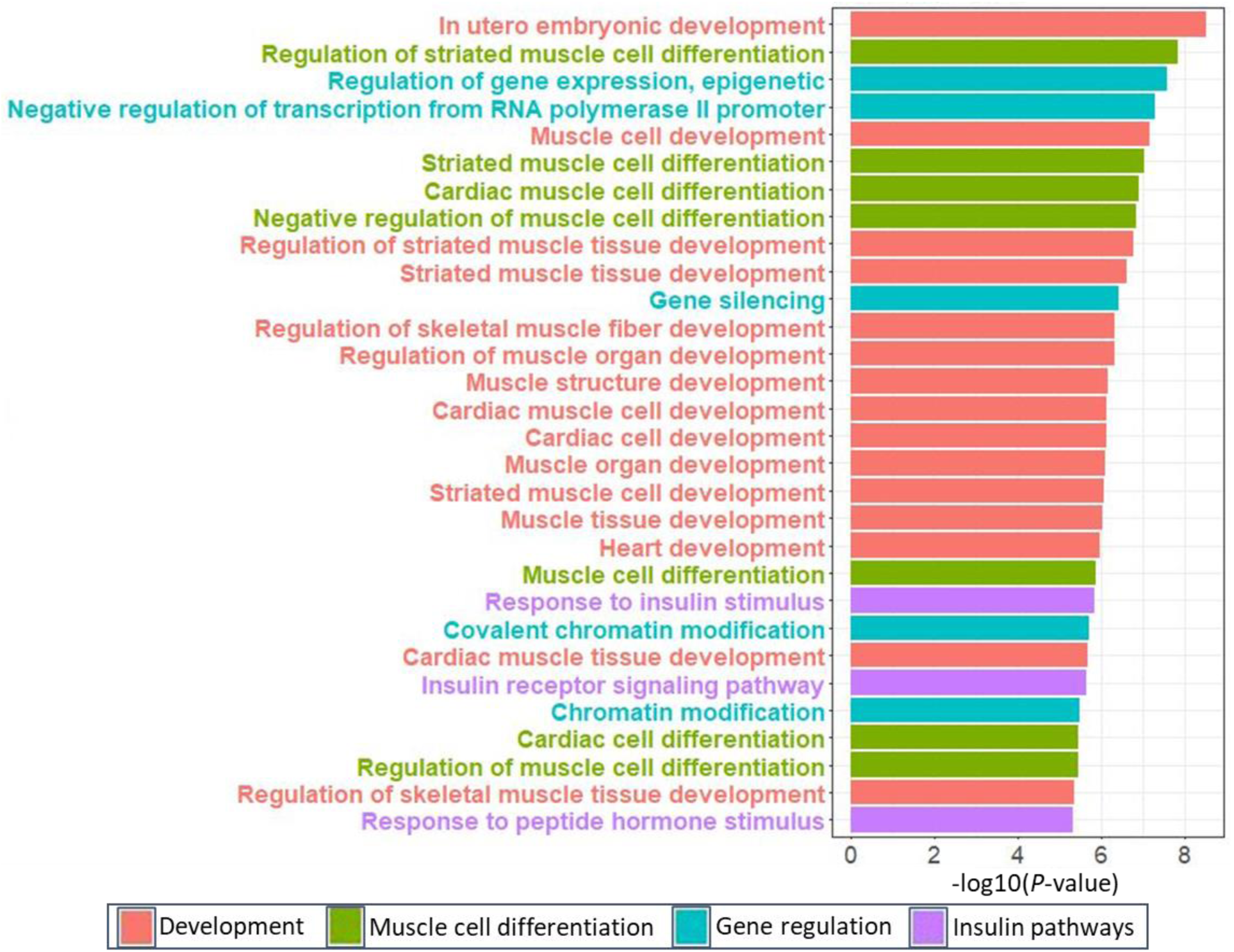
Top enriched Gene-Ontology biological processes for QT. Top GO terms (biological processes only) for QT. Due to the number of results, this figure has been limited to the top 30. Full results can be found in Supplementary Table 16. Results are color coded according to broad categories “Development”, “Muscle cell differentiation”, “Gene regulation”, “Insulin pathways”. *P*-value for enrichment is displayed on the -log base 10 scale.

**Supplementary Figure 9:**
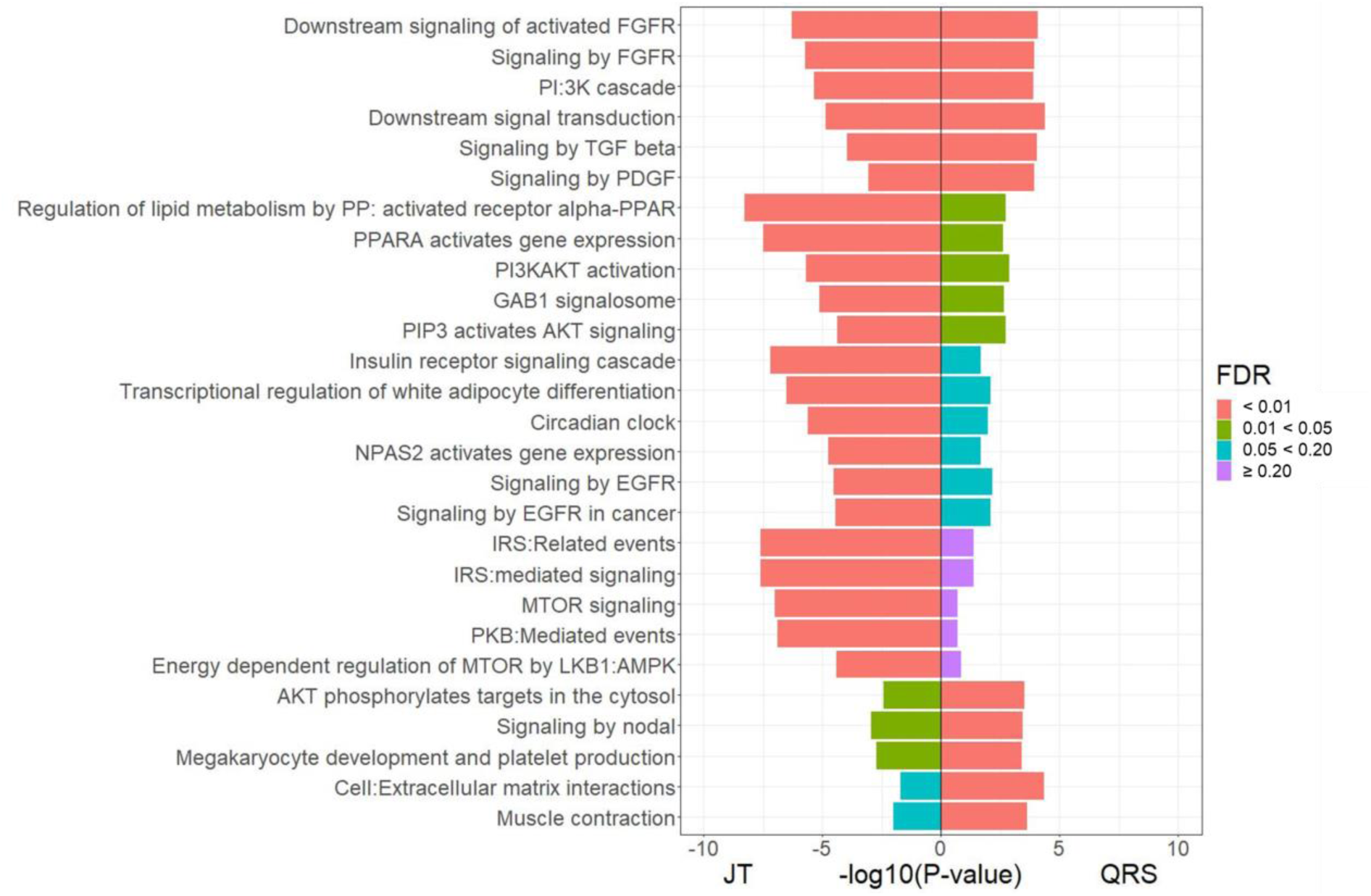
Top enriched Reactome pathways for JT and QRS. Comparison of lead Reactome pathways for JT and QRS from DEPICT analyses (methods). Results are color coded according to FDR (False discovery rate) and *P*-value for enrichment displayed on the log base 10 scale. AKT: Protein kinase B, EGFR: Epidermal growth factor receptor, FGFR: Fibroblast growth factor receptor, IRS: Insulin receptor signalling, MTOR: mammalian target of rapamycin, PI:3K: Phosphatidlyinositol-3-kinase, PDGF: Platelet-derived growth factor, PP: peroxisome proliferator, PPAR: Peroxisome proliferator-activated receptor, Full results can be found in Supplementary Table 16.

## Supplementary Note 1 – Study information for genome-wide association study

### ARIC - Atherosclerosis Risk in Communities

The Atherosclerosis Risk in Communities (ARIC) Study is a prospective community-based study of cardiovascular disease and its risk factors. At baseline (1987-89), 15,792 men and women age 45-64 were recruited from 4 communities in the US (Washington County, Maryland; Forsyth County, North Carolina; Jackson, Mississippi; Minneapolis suburbs, Minnesota). Participants were mostly white in the Minnesota and Washington County field centers, white and African American in Forsyth County, and exclusively African American in the Jackson field center. Cohort members completed five clinic examinations, conducted approximately three years apart between 1987 and 1998, with a fifth visit conducted from 2011 – 2013. Clinic examinations included assessment of cardiovascular risk factors, self-reported medical family history, employment and educational status, diet, physical activity, comorbidity, clinical and laboratory measurements. The present analyses utilized ECG measurements from the baseline assessment.

Digital 12-lead ECGs were obtained using MAC PC Personal Cardiographs (Marquette Electronics Inc., Milwaukee, WI) and were subsequently submitted to a central reading center at the EPICORE Center (University of Alberta, Edmonton, Alberta, Canada) and thereafter to the Epidemiological Cardiology Research Center (EPICARE), Wake Forest University, Winston-Salem, NC. All ECGs were visually inspected for quality and legibility at their acquisition and by the reading centers.

For assessment of QT interval, participants were asked not to smoke or ingest caffeine for at least 1 hour prior to the ECG being obtained. After resting for 5-10 minutes while the electrodes were being placed, a standard supine 12-lead electrocardiogram and a 2-minute paper recording of a three-lead (leads V1, II, and V5) rhythm strip were made. The QT interval from the digital 12-lead ECG was determined by identifying Q-onset and T-wave offset in all three leads. T-wave offset was defined as the point of maximum change of slope as the T-wave merges with the baseline as implemented in the Novacode ECG measurement and classification program as has been described in detail (https://pubmed.ncbi.nlm.nih.gov/9682893/) and used in prior ARIC studies of the QT interval (https://pubmed.ncbi.nlm.nih.gov/14975464/). U-waves were not detected by the Novacode algorithm. QRS and JT interval were measured automatically from baseline ECGs.

### Bambui - Brazilian Bambuí Cohort Study of Ageing

A cohort study designed to identify predictors of adverse health events in the elderly. The study population comprises all residents of Bambuí (Minas Gerais, Brazil), aged 60 or more years (n=1.742) at the baseline in 1997. From these, 92.2% were interviewed and 85.9% underwent clinical examination, consisting of hematological and biochemical tests, serology for Trypanosoma cruzi, anthropometric and blood pressure measures and electrocardiogram. Cohort members undergo annual follow-up visits, which consist of an interview and verification of death certificates. Other procedures were repeated in selected years (2000, 2002 and 2008). From 1997 to 2007, during a mean follow-up of 8.6 years, 641 participants died and 96 (6.0%) were lost to follow-up.

### BioMe - The IPM Bio*Me* Biobank

The Mount Sinai BioMe Biobank, founded in September 2007, is an ongoing, broadly consented EHR-linked bio- and data repository that enrolls participants non-selectively from the Mount Sinai Medical Center patient population. The BioMe Biobank draws from a population of over 70,000 inpatient and 800,000 outpatient visits annually from over 30 broadly selected clinical sites of the Mount Sinai Medical Center (MSMC). As of September 2017, BioMe has enrolled more than 42,000 patients that represent a broad racial, ethnic and socioeconomic diversity with a distinct and population-specific disease burden, characteristic of the communities served by Mount Sinai Hospital. BioMe participants are predominantly of African (AA, 24%), Hispanic/Latino (HL, 35%), European (EA, 32%), and other ancestry (OA, 10%). The BioMe Biobank Program operates under a Mount Sinai Institutional Review Board-approved research protocol. All study participants provided written informed consent.

### BRIGHT - British Genetics of Hypertension

Participants of the BRIGHT Study were recruited from the Medical Research Council General Practice Framework and other primary care practices in the UK. Each case had a history of hypertension diagnosed prior to 60 years of age with confirmed blood pressure recordings corresponding to seated levels >150/100mmHg (1 reading) or mean of 3 readings >145/95 mmHg. BRIGHT is focused on recruitment of hypertensive individuals with BMI<30. Sample selection for GWAS was based on DNA availability and quantity.

### Broad AF

The Broad AF Study is a collaborative project to investigate the genetic determinants of atrial fibrillation (AF), comprised of 17,517 AF cases and 10,987 referents from 26 studies. Details of study description, genotyping, and imputation were described previously. 2 Briefly, genetic variants were centrally genotyped on the Infinium PsychArray-24 v1.2 Bead Chip, and jointly 8 called and quality controlled at the Broad Institute. After pre-imputation quality control, variants were imputed using the 1000 Genomes reference panel. A total of 3,461 individuals free of AF met the inclusion criteria of the current study and were included in the PR analysis. Individuals from the following participating studies were included: Vanderbilt University Medical Center Biobank (BioVU), Australian Familial AF Study, Danish AF Study, Groningen Genetics of Atrial Fibrillation (GGAF), Genetic Risk Assessment of Defibrillator Events Study (GRADE), Malmö Preventive Project (MPP-AF, and MPP-Echo), and Intermountain INSPIRE Registry. Study details were previously reported.

### CAMP-MGH - MGH Cardiology and Metabolic Patient Cohort

The MGH Cardiology and Metabolic Patient Cohort is comprised of 3850 subjects recruited from the ambulatory MGH Cardiology Practice between 2009 and 2012.

### CHRIS - The Cooperative Health Research in South Tyrol study

The Cooperative Health Research In South Tyrol (CHRIS) study is a population-based study with a longitudinal lookout to investigate the genetic and molecular basis of age-related common chronic conditions and their interaction with life style and environment in the general population. The study was approved by the Ethics Committee of the Autonomous Province of Bolzano.

### CHS - Cardiovascular Health Study

The Cardiovascular Health Study (CHS) is a population-based cohort study of risk factors for coronary heart disease and stroke in adults ≥65 years conducted across four field centers . The original predominantly European ancestry cohort of 5,201 persons was recruited in 1989-1990 from random samples of the Medicare eligibility lists; subsequently, an additional predominantly African-American cohort of 687 persons was enrolled in 1992-1993 for a total sample of 5,888.

Blood samples were drawn from all participants at their baseline examination and DNA was subsequently extracted from available samples. Genotyping was performed at the General Clinical Research Center’s Phenotyping/Genotyping Laboratory at Cedars-Sinai among CHS participants who consented to genetic testing and had DNA available using the Illumina 370CNV BeadChip system (for European ancestry participants, in 2007) or the Illumina HumanOmni1-Quad_v1 BeadChip system (for African-American participants, in 2010).

CHS was approved by institutional review committees at each field center and individuals in the present analysis had available DNA and gave informed consent including consent to use of genetic information for the study of cardiovascular disease.

### ERF - Erasmus Rucphen Family Study

The Erasmus Rucphen Family study is comprised of a family-based cohort embedded in the Genetic Research in Isolated Populations (GRIP) program in the southwest of the Netherlands. The aim of this program is to identify genetic risk factors for the development of complex disorders. In ERF, twenty-two families that had a large number of children baptized in the community church between 1850 and 1900 were identified with the help of detailed genealogical records. All living descendants of these couples, and their spouses, were invited to take part in the study. Comprehensive interviews, questionnaires, and examinations were completed at a research center in the area; approximately 3,200 individuals participated. Examinations included 12 lead ECG measurements.

Electrocardiograms were recorded on ACTA electrocardiographs (ESAOTE, Florence, Italy) and digital measurements of the QRS and QT intervals were made using the Modular ECG Analysis System (MEANS). Data collection started in June 2002 and was completed in February 2005. In the current analyses, 2442 participants for whom complete phenotypic, genotypic and genealogical information was available were studied.

### FINCAVAS - Finnish Cardiovascular Study

The purpose of the Finnish Cardiovascular Study (FINCAVAS) is to construct a risk profile - using genetic, hemodynamic and electrocardiographic (ECG) markers - of individuals at high risk of cardiovascular diseases, events and deaths. All patients scheduled for an exercise stress test at Tampere University Hospital and willing to participate have been recruited between October 2001 and December 2007. The final number of participants is 4,567. In addition to repeated measurement of heart rate and blood pressure, digital high-resolution ECG at 500 Hz was recorded continuously during the entire exercise test, including the resting and recovery phases. About 20% of the patients are were examined with coronary angiography. Genetic variations known or suspected to alter cardiovascular function or pathophysiology are were analyzed to elucidate the effects and interactions of these candidate genes, exercise and commonly used cardiovascular medications.

### GAPP - Genetic and phenotypic determinants of blood pressure and other cardiovascular risk factors

GAPP is a population-based prospective cohort study involving a representative sample of healthy adults aged 25-41 years residing in the Principality of Liechtenstein. Exclusion criteria were the presence of cardiovascular disease, diabetes, obstructive sleep apnea and a body mass index >35kg/m2. A standardized 12-lead ECG was obtained in all participants.

### GESUS - The Danish General Suburban Population Study

GESUS is a population-based prospective cohort study from Naestved Municipality (70km south of Copenhagen), Denmark. The study enrolled 21,205 adult participants between 2010-2013. Age was 20 years or above (20-100 years). All participants answered a questionnaire and had a physical examination (including EKG, laboratory tests, anthropometrics, biological samples for biobank etc.) performed at the Department of Clinical Biochemistry, Naestved Hospital, Denmark. A standard 12- lead paper ECG was obtained in all participants, and a corresponding electronic ECG was obtained in 8939 participants. ECG information was obtained from the MUSE Cardiology Information System (GE Healthcare, Wauwatosa, Wisconsin, USA) and analyzed by Marquette 12SL algorithm version 21. The ECGs were recorded with a sample rate of 500 Hz and a resolution of 4.88 μV per least significant bit. At the time of this study, 3004 participants were genotyped and included.

### GS20 - Generation Scotland: Scottish Family Health Study

The GS:SFHS study recruited 23,960 participants aged 18–100 years between 2006–11; full details are reported elsewhere [Smith et al, IJE 2012]. Participants came from across Scotland, with some family members from further afield. The sample was 59% female, with a wide range of ages and socio-demographic characteristics. Most (87%) participants were born in Scotland and 96% in the UK or Ireland.

### HCHS/SOL - Hispanic Community Health Survey/Study of Latinos

The Hispanic Community Health Study/ Study of Latinos (HCHS/SOL) The HCHS/SOL is a multicenter, community-based cohort study of U.S. Hispanics/Latinos. Goals of the study are to examine the prevalence of and risk factors for several disorders including heart, lung, blood, and kidney phenotypes. HCHS/SOL investigators sampled 16,415 males and females aged 18-74 years at baseline from four study communities: The Bronx, NY, Chicago, IL, Miami, FL, and San Diego, CA. HCHS/SOL recruitment centers were selected so that the study would include at least 2,000 participants in each of the following designations: Mexican, Puerto Rican, Dominican, Cuban, and Central and South American.

### Health ABC - Health, Aging, and Body Composition Study

The Health Aging and Body Composition (Health ABC) Study is a NIA-sponsored cohort study of the factors that contribute to incident disability and the decline in function of healthier older persons, with a particular emphasis on changes in body composition in old age. Between 4/15/97 and 6/5/98 the Health ABC study has recruited 3,075 70-79 year old community-dwelling adults (41% African-American), who were initially free of mobility and activities of daily living disability. The key components of Health ABC include a baseline exam, annual follow-up clinical exams, and phone contacts every 6 months to identify major health events and document functional status between clinic visits. Provision has been made for banking of blood specimens and extracted DNA (HealthABC repository).

### INGI-CAR - INGI-CARLANTINO

INGI-CAR consisted of about 1000 subjects who were drawn from Carlantino, an isolated village of southern Italy. Ethics approval was obtained from the Ethics Committee of the IRCCS Burlo Garofolo in Trieste. Written informed consent was obtained from every participant of the study. The study population had undergone clinical and instrumental evaluations between 1998 and 2005. For all subjects, anthropometrics variables (such as height, weight, etc.) were taken and a structured questionnaire about lifestyle and medical history was filled out. In addition, blood pressure, body-mass index, biochemical analyses, ECG and cardiovascular evaluation were collected.

### INGI-FVG - INGI-Friuli Venezia Giulia

The INGI-FVG cohort consisted of about 1700 subjects drawn from the project “Genetic Park of Friuli Venezia Giulia”. This study examined 6 isolated villages in the North-East of Italy between 2008 and 2010. Ethics approval was obtained from the Ethics Committee of the IRCCS Burlo Garofolo in Trieste. Written informed consent was obtained from every participant of the study. The study population had undergone clinical and instrumental evaluations. For all subjects, anthropometrics variables (such as height, weight, etc) were taken and a structured questionnaire about lifestyle and medical history was filled out. In addition, blood pressure, body-mass index, biochemical analyses, ECG and cardiovascular evaluation were collected.

### INTER99 - Inter99

The Inter99 study carried out in 1999-2001 included invitation of12934 persons aged 30-60 years drawn from an age- and sex-stratified random sample of the population (16). The baseline participation rate was 52.5%, and the study included 6784 persons. The Inter99 study was a population-based randomized controlled trial (CT00289237, ClinicalTrials.gov) and investigated the effects of lifestyle intervention on CVD. Here 5827 participants with information on lipids and exome chip were analyzed. ECG information was obtained from the MUSE Cardiology Information System (GE Healthcare, Wauwatosa, Wisconsin) analyzed by Marquette 12SL algorithm version 21.

### JHS - Jackson Heart Study

The JHS is a single-site cohort study of 5,306 extensively phenotyped African American women and men. Three clinical examinations have been completed, including the baseline examination, Examination 1 (2000–2004), Examination 2 (2005–2008), and Examination 3 (2009–2013), allowing comprehensive assessment of cardiovascular health and disease of the cohort at approximately four-year intervals. Ongoing monitoring of hospitalizations for cardiovascular events (coronary heart disease, heart failure and stroke) and deaths among cohort participants are accomplished by annual telephone follow-up interviews, surveillance of hospital discharge records (since 2000 for coronary heart disease and stroke, and since 2005 for heart failure), and vital records.

### KORA F3/S4 - Kooperative Gesundheitsforschung in der Region Augsburg F3/S4

The KORA study is a series of independent population-based epidemiological surveys of participants living in the city of Augsburg, Southern Germany, or the two adjacent counties. All survey participants are residents of German nationality identified through the registration office and aged between 25 and 74 years at recruitment. KORA F3: The baseline survey KORA S3 was conducted in the years 1994/95. 3,006 participants from KORA S3 were reexamined in a 10-year follow-up (KORA F3) in the years 2004/05. KORA S4: All survey participants are residents of German nationality identified through the registration office and aged between 25 and 74 years at recruitment. The baseline survey KORA S4 was conducted in the years 1999-2001.

### LIFELINES - LifeLines, a three-generation cohort study and biobank

The LifeLines Cohort Study, and generation and management of GWAS genotype data for the LifeLines Cohort Study is supported by the Netherlands Organization of Scientific Research NWO (grant 175.010.2007.006), the Economic Structure Enhancing Fund (FES) of the Dutch government, the Ministry of Economic Affairs, the Ministry of Education, Culture and Science, the Ministry for Health, Welfare and Sports, the Northern Netherlands Collaboration of Provinces (SNN), the Province of Groningen, University Medical Center Groningen, the University of Groningen, Dutch Kidney Foundation and Dutch Diabetes Research Foundation.

### MESA - Multi-Ethnic Study of Atherosclerosis

The Multi-Ethnic Study of Atherosclerosis (MESA) is a study of the characteristics of subclinical cardiovascular disease (disease detected non-invasively before it has produced clinical signs and symptoms) and the risk factors that predict progression to clinically overt cardiovascular disease or progression of the subclinical disease. The cohort is a diverse, population-based sample of 6,814 asymptomatic men and women aged 45-84. Approximately 38 percent of the recruited participants are white, 28 percent African-American, 22 percent Hispanic, and 12 percent Asian (predominantly of Chinese descent). Participants were recruited during 2000-2002 from 6 field centers across the U.S. (at Wake Forest University; Columbia University; Johns Hopkins University; the University of Minnesota; Northwestern University; and the University of California – Los Angeles). All underwent anthropomorphic measurement and extensive evaluation by questionnaires at baseline, followed by 5 subsequent examinations at intervals of approximately 2-4 years. Age and sex were self-reported.

### NEO - Netherlands Epidemiology of Obesity

The NEO was designed for extensive phenotyping to investigate pathways that lead to obesity-related diseases. The NEO study is a population-based, prospective cohort study that includes 6,671 individuals aged 45–65 years, with an oversampling of individuals with overweight or obesity. At baseline, information on demography, lifestyle, and medical history have been collected by questionnaires. In addition, samples of 24-h urine, fasting and postprandial blood plasma and serum, and DNA were collected. Genotyping was performed using the Illumina HumanCoreExome chip, which was subsequently imputed to the 1000 genome reference panel. Participants underwent an extensive physical examination, including anthropometry, electrocardiography, spirometry, and measurement of the carotid artery intima-media thickness by ultrasonography. In random subsamples of participants, magnetic resonance imaging of abdominal fat, pulse wave velocity of the aorta, heart, and brain, magnetic resonance spectroscopy of the liver, indirect calorimetry, dual energy X-ray absorptiometry, or accelerometry measurements were performed. The collection of data started in September 2008 and completed at the end of September 2012. Participants are currently being followed for the incidence of obesity-related diseases and mortality.

### OOA - Old Order Amish

The Old Order Amish (OOA) subjects included in this study were participants of several studies of cardiovascular health in relatively healthy volunteers from the OOA community of Lancaster County, PA and their family members. The studies were carried out at the University of Maryland as part of the Amish Complex Disease Research Program (ACDRP). The OOA population of Lancaster County, PA immigrated to the Colonies from Western Europe in the early 1700’s. All study protocols were approved by the institutional review board at the University of Maryland and participating institutions. Informed consent was obtained from each of the study participants.

### ORCADES - Orkney Complex Disease Study

The Orkney Complex Disease Study (ORCADES) is a family-based, cross-sectional study that seeks to identify genetic factors influencing cardiovascular and other disease risk in the isolated archipelago of the Orkney Isles in northern Scotland (McQuillan et al., 2008). Genetic diversity in this population is decreased compared to Mainland Scotland, consistent with the high levels of endogamy historically. 2078 participants aged 16-100 years were recruited between 2005 and 2011, most having three or four grandparents from Orkney, the remainder with two Orcadian grandparents. Fasting blood samples were collected and many health-related phenotypes and environmental exposures were measured in each individual. All participants gave written informed consent and the study was approved by Research Ethics Committees in Orkney and Aberdeen (North of Scotland REC).

### PIVUS - Prospective Investigation of the Vasculature of Uppsala Seniors

The Prospective Investigation of Vasculature in Uppsala Seniors study was initiated in 2001 to investigate the predictive power of different measurements of vascular characteristics for future cardiovascular events, and secondary aims included measurements of cardiac and metabolic function, as well as serum biomarkers and levels of environmental pollutants. All individuals aged 70 living in the community of Uppsala in Sweden were deemed eligible for the study. The subjects were selected from the community register and invited in randomized order between April 2001 and June 2004.

They received an invitation letter for participation within 2 months of their 70th birthday. Of the 2,025 subjects invited, 1,016 (507 male, 509 female) subjects agreed to participate. The participants were asked to answer a questionnaire about their medical history, smoking habits and regular medication.

### PREVEND - Prevention of REnal and Vascular ENd stage Disease

Prevend is an ongoing prospective study investigating the natural course of increased levels of urinary albumin excretion and its relation to renal and cardiovascular disease. Details of the protocol have been described elsewhere.

### PROSPER - PROSpective study of pravastatin in the elderly at Risk for vascular disease

All data come from the PROspective Study of Pravastatin in the Elderly at Risk (PROSPER). A detailed description of the study has been published elsewhere. PROSPER was a prospective multicenter randomized placebo-controlled trial to assess whether treatment with pravastatin diminishes the risk of major vascular events in elderly. Between December 1997 and May 1999, we screened and enrolled subjects in Scotland (Glasgow), Ireland (Cork), and the Netherlands (Leiden). Men and women aged 70-82 years were recruited if they had pre-existing vascular disease or increased risk of such disease because of smoking, hypertension, or diabetes. A total number of 5,804 subjects were randomly assigned to pravastatin or placebo. A large number of prospective tests were performed including Biobank tests and cognitive function measurements. A whole genome wide screening has been performed in the sequential PHASE project. Of 5,763 subjects DNA was available for genotyping. Genotyping was performed with the Illumina 660K beadchip, after QC (call rate <95%) 5,244 subjects and 557,192 SNPs were left for analysis. These SNPs were imputed to 2.5 million SNPs based on the HAPMAP built 36 with MACH imputation software. The study was approved by the institutional ethics review boards of centers of Cork University (Ireland), Glasgow University (Scotland) and Leiden University Medical Center (the Netherlands) and all participants gave written informed consent.

### RS - Rotterdam Study

Rotterdam Study, a prospective population-based cohort study. Details regarding design, objectives, and methods of the Rotterdam Study have been described in detail. (Hofman et al, 2016) In short, the Rotterdam study started in 1989 with an initial cohort of 7,983 persons (out of 10,215 invitees; response rate 78%) 55 years of age or older living in the Ommoord district in the city of Rotterdam in the Netherlands. In 2000, 3,011 participants (out of 4,472 invitees, response rate 67%) who had become 55 years of age or moved into the study district were added to the cohort. As of In 2006, a further extension of the cohort was initiated in which 3932 subjects were included, aged 45–54 years, out of 6057 invited, living in the Ommoord district. In summer of 2016, the recruitment of another extension started that targeted participants aged 40 years and over. The establishment of this extension is expected to be completed by early 2020 and to yield around 3000 new participants. The participants were all extensively examined at study entry (i.e. baseline) and subsequent follow-up visits that take place every 3 to 6 years. They were interviewed at home (2 h) and then underwent an extensive set of examinations (a total of 5 h) in a specially built research facility in the centre of the district. These examinations focused on possible causes of invalidating diseases in the elderly in a clinically state-of-the-art manner, as far as the circumstances allowed. The emphasis was put on imaging (of heart, blood vessels, eyes, skeleton and later brain) and on collecting biospecimens that enabled further in-depth molecular and genetic analyses. Approximately every 4-5 years follow-up examinations are conducted. Examinations consist of a home interview and an extensive set of test at a research facility in the study district. By linking the general practitioners’ and municipality records to the study database, participants are continuously monitored for major morbidity and mortality.

### SardiNIA - SardiNIA Project

To identify genetic bases for prominent age-associated changes, including cardiovascular risk factors and determinants of personality traits, in a founder population. The results of the study will extend the studies of aging-associated conditions of outbred populations.

### SHIP-START and SHIP-TREND - Study of Health in Pomerania

The Study of Health In Pomerania is a prospective longitudinal population-based cohort study in Western Pomerania assessing the prevalence and incidence of common diseases and their risk factors. SHIP encompasses the two independent cohorts SHIP-START and SHIP-TREND. The detailed study design has been published previously. In brief, participants aged 20 to 79 with German citizenship and principal residency in the study area were recruited from a random sample of residents living in the three local cities (with 17,076 to 65,977 inhabitants), 12 towns (with 1,516 to 3,044 inhabitants) as well as 17 out of 97 (with less than 1,500 inhabitants) randomly selected smaller towns. Individuals were randomly selected in proportion to the population size of the community and stratified by age and sex. For SHIP-START, a total of 4,308 participants were recruited between 1997 and 2001. Between 2008 and 2012 a total of 4,420 participants were recruited in the SHIP-TREND cohort. Individuals were invited to the SHIP study center for computer-assisted personal interviews and extensive physical examinations. Individuals of both cohorts were analyzed separately.

### TWINSUK - TWINSUK

There are currently >13,500 twins registered participants in the TwinsUK study, of which over 9,000 are actively participating. The twins are aged 16 to 100 with approximately equal numbers of identical (MZ) and non-identical (DZ) twins and are predominantly female (80%) for historical reasons. Clinical, physiological, behavioral and lifestyle data is collected at either twin visits or via self-administered questionnaires, which volunteers complete either once or twice a year via the post or email. All studies have ethical approval from the Guy’s and St Thomas’ (GSTT) Ethics Committee.

### UK Biobank - UK Biobank Study

UK Biobank (UKB, www.ukbiobank.ac.uk) is a large longitudinal biobank study in the United Kingdom which was established to improve understanding of the genetic and environmental causes of common diseases including CVDs. In addition to self-reported disease outcomes and extensive health and life-style questionnaire data, UKB participants are being tracked through their NHS records and national registries (including cause of death and Hospital Episode Statistics). In 2017, UKB released the genotypes of 488,377 participants profiled with a custom SNP array. Genotyping QC was performed centrally by UKB, and genotypes imputed to Haplotype Reference Consortium (HRC) panel were released for 488,377 participants. For the UKB-EXECG sub-cohort, ECG measures were calculated from 4-lead ECGs (CAM-USB 6.5, Cardiosoft v6.51) recorded during a 15 second rest period prior to an exercise test while subjects were sitting on a stationary bike (eBike, Firmware v1.7). Electrodes were placed on the right and left antecubital fossae, and left and right wrist and the ECG was sampled at 500 Hz; lead I was used to derive ECG measures. To reduce the influence of noise, QT, JT and QRS were measured from a signal averaged ECG waveform computed from the heartbeats available in the 15s trace. For this, we first identified the QRS-complexes using fully automatic in-house algorithms. Before averaging, we removed ectopic beats and artefacts, as well as beats with an RR interval longer or shorter than 10ms compared to the mean RR interval were not included for averaging. A signal-averaged heartbeat was then computed from the remaining beats provided that the number of available beats was not less than 5. For the UKB-12lead sub-cohort, ECG measures were calculated from a resting 12-lead ECG. As described above, to reduce the influence of noise, ECG measures were calculated from signal averaged ECG wave forms computed from lead I. When individuals had both 12-lead ECG and exercise test ECG data, the 12-lead recording was used, to ensure no overlap between sub-cohorts.

### VIKING - Viking Health Study

The Viking Health Study - Shetland (VIKING) is a family-based, cross-sectional study that seeks to identify genetic factors influencing cardiovascular and other disease risk in the population isolate of the Shetland Isles in northern Scotland. Genetic diversity in this population is decreased compared to Mainland Scotland, consistent with the high levels of endogamy historically. 2105 participants were recruited between 2013 and 2015, most having at least three grandparents from Shetland. Fasting blood samples were collected and many health-related phenotypes and environmental exposures were measured in each individual. All participants gave informed consent and the study was approved by the South East Scotland Research Ethics Committee.

### WHI - Women’s Health Initiative

The Women’s Health Initiative (WHI) is a long-term national health study focused on strategies for preventing heart disease, breast and colorectal cancer, and osteoporotic fractures in postmenopausal women. Launched in 1993, the WHI enrolled 161,808 women aged 50-79 into one or more randomized Clinical Trials (CT), testing the health effects of hormone therapy (HT), dietary modification (DM), and/or calcium and Vitamin D supplementation (CaD) or to an Observational Study (OS).

This ground-breaking study changed the way health care providers prevent and treat some of the major diseases impacting postmenopausal women. Results from the WHI Hormone Trials have been estimated to have already saved $35.2 billion in direct medical costs in the US alone. To date, WHI has published over 1,400 articles and approved and funded 289 ancillary studies. The GWAS data used in this paper comes from six ancillary studies.

### YFS - Young Finns Study

The YFS is a population-based follow up-study started in 1980. The main aim of the YFS is to determine the contribution made by childhood lifestyle, biological and psychological measures to the risk of cardiovascular diseases in adulthood. In 1980, over 3,500 children and adolescents all around Finland participated in the baseline study. The follow-up studies have been conducted mainly with 3- year intervals. The 27-year follow-up study was conducted in 2007 (ages 30-45 years) with 2,204 participants. The study was approved by the local ethics committees (University Hospitals of Helsinki, Turku, Tampere, Kuopio and Oulu) and was conducted following the guidelines of the Declaration of Helsinki. All participants gave their written informed consent.

## Supplementary Note 2 – Codes used to define each clinical outcome in the UK Biobank

ICD10 and ICD9 codes were used to extract hospital episode statistics and death registry information. OPCS4 are operation codes also used. The following codes were used to define atrial fibrillation, stroke, coronary artery disease, atrioventricular block/pacemaker implantation, bundle branch block/fascicular block, heart failure, non-ischaemic cardiomyopathy and ventricular arrhythmia.

|  |  |
| --- | --- |
| **Atrial Fibrillation** |  |
| ICD10 codes | Definition |
| I48 | Atrial fibrillation |
| I48.0 | Paroxysmal atrial fibrillation |
| I48.1 | Persistent atrial fibrillation |
| I48.2 | Chronic atrial fibrillation |
| ICD9 codes | Definition |
| 4273 | Atrial fibrillation and flutter |
| OPCS4 | Definition |
| K62.1 | Percutaneous transluminal ablation of pulmonary vein to left atrium conducting system |
| K62.5 | Percutaneous transluminal occlusion of left atrial appendage |
| K57.1 | Percutaneous transluminal ablation of atrioventricular node |
| K57.2 | Percutaneous transluminal ablation of conducting system of heart NEC |
| **Stroke** |  |
| ICD10 codes | Definition |
| I63.0 | Cerebral infarction due to thrombosis of precerebral arteries |
| I63.1 | Cerebral infarction due to embolism of precerebral arteries |
| I63.2 | Cerebral infarction due to unspecified occlusion or stenosis of precerebral arteries |
| I63.3 | Cerebral infarction due to thrombosis of cerebral arteries |
| I63.4 | Cerebral infarction due to embolism of cerebral arteries |
| I63.5 | Cerebral infarction due to unspecified occlusion or stenosis of cerebral arteries |
| I63.8 | Other cerebral infarction |
| I63.9 | Cerebral infarction, unspecified |
| I61.0 | Intracerebral hemorrhage in hemisphere, subcortical |
| I61.1 | Intracerebral hemorrhage in hemisphere, cortical |
| I61.2 | Intracerebral hemorrhage in hemisphere, unspecified |
| I61.3 | Intracerebral hemorrhage in brain stem |
| I61.4 | Intracerebral hemorrhage in cerebellum |
| I61.5 | Intracerebral hemorrhage, intraventricular |
| I61.6 | Intracerebral hemorrhage, multiple localized |
| I61.8 | Other intracerebral hemorrhage |
| I61.9 | Intracerebral hemorrhage, unspecified |
| I64 | Stroke, not specified as hemorrhage or infarction |
| ICD9 codes | Definition |
| 434.91 | Cerebral infarction due to thrombosis of precerebral arteries |
| 434.91 | Cerebral infarction due to embolism of precerebral arteries |
| 434.91 | Cerebral infarction due to unspecified occlusion or stenosis of precerebral arteries |
| 434.01 | Cerebral infarction due to thrombosis of cerebral arteries |
| 434.11 | Cerebral infarction due to embolism of cerebral arteries |
| 434.91 | Cerebral infarction due to unspecified occlusion or stenosis of cerebral arteries |
| 434.91 | Cerebral infarction, unspecified |
| 436 | Stroke, not specified as hemorrhage or infarction |
| **Ischemic stroke** |  |
| ICD10 codes | Definition |
| I63.0 | Cerebral infarction due to thrombosis of precerebral arteries |
| I63.1 | Cerebral infarction due to embolism of precerebral arteries |
| I63.2 | Cerebral infarction due to unspecified occlusion or stenosis of precerebral arteries |
| I63.3 | Cerebral infarction due to thrombosis of cerebral arteries |
| I63.4 | Cerebral infarction due to embolism of cerebral arteries |
| I63.5 | Cerebral infarction due to unspecified occlusion or stenosis of cerebral arteries |
| I63.8 | Other cerebral infarction |
| I63.9 | Cerebral infarction, unspecified |
| ICD9 codes | Definition |
| 434.91 | Cerebral infarction due to thrombosis of precerebral arteries |
| 434.91 | Cerebral infarction due to embolism of precerebral arteries |
| 434.91 | Cerebral infarction due to unspecified occlusion or stenosis of precerebral arteries |
| 434.01 | Cerebral infarction due to thrombosis of cerebral arteries |
| 434.11 | Cerebral infarction due to embolism of cerebral arteries |
| 434.91 | Cerebral infarction due to unspecified occlusion or stenosis of cerebral arteries |
| 434.91 | Cerebral infarction, unspecified |
| **Coronary artery disease** |  |
| ICD10 codes | Definition |
| I20 | Unstable angina |
| I21.0 | Acute transmural myocardial infarction of anterior wall |
| I21.1 | Acute transmural myocardial infarction of inferior wall |
| I21.2 | Acute transmural myocardial infarction of other sites |
| I21.3 | Acute transmural myocardial infarction of unspecified site |
| I21.4 | Acute subendocardial myocardial infarction |
| I21.9 | Acute myocardial infarction, unspecified |
| I22.0 | Subsequent myocardial infarction of anterior wall |
| I22.1 | Subsequent myocardial infarction of inferior wall |
| I22.8 | Subsequent myocardial infarction of other sites |
| I22.9 | Subsequent myocardial infarction of unspecified site |
| I23.0 | Hemopericardium as current complication following acute myocardial infarction |
| I23.1 | Atrial septal defect as current complication following acute myocardial infarction |
| I23.2 | Ventricular septal defect as current complication following acute myocardial infarction |
| I23.3 | Rupture of cardiac wall without hemopericardium as current complication following acute myocardial infarction |
| I23.4 | Rupture of chordae tendineae as current complication following acute myocardial infarction |
| I23.5 | Rupture of papillary muscle as current complication following acute myocardial infarction |
| I23.6 | Thrombosis of atrium , auricular appendage and ventricle as current complications following acute myocardial infarction |
| I23.8 | Other current complications following acute myocardial infarction |
| I24.0 | Coronary thrombosis not resulting in myocardial infarction |
| I24.8 | Other forms of acute ischemic heart disease |
| I24.9 | Acute ischemic heart disease, unspecified |
| I25.0 | Atherosclerotic cardiovascular disease, so described |
| I25.1 | Atherosclerotic heart disease |
| I25.2 | Old myocardial infarction |
| I25.3 | Aneurysm of heart |
| I25.4 | Coronary artery aneurysm |
| I25.5 | Ischemic cardiomyopathy |
| I25.6 | Silent myocardial ischemia |
| I25.8 | Other forms of chronic ischemic heart disease |
| I25.9 | Chronic ischemic heart disease, unspecified |
| ICD9 codes | Definition |
| 4109 | Acute myocardial infarction |
| 4119 | Other acute and subacute forms of ischemic heart disease |
| 4129 | Old myocardial infarction |
| 4139 | Angina pectoris |
| 4140 | Coronary atherosclerosis |
| 4141 | Aneurysm of heart |
| 4148 | Other specified forms of chronic ischemic heart disease |
| 4149 | Chronic ischemic heart disease, unspecified |
| OPCS4 | Definition |
| K40.1 | Saphenous vein graft replacement of one coronary artery |
| K40.2 | Saphenous vein graft replacement of two coronary arteries |
| K40.3 | Saphenous vein graft replacement of three coronary arteries |
| K40.4 | Saphenous vein graft replacement of four or more coronary arteries |
| K40.9 | Unspecified saphenous vein graft replacement of coronary artery |
| K41.1 | Autograft replacement of one coronary artery NEC |
| K41.2 | Autograft replacement of two coronary arteries NEC |
| K41.3 | Autograft replacement of three coronary arteries NEC |
| K41.4 | Autograft replacement of four or more coronary arteries NEC |
| K42.4 | Allograft replacement of four or more coronary arteries |
| K44.1 | Replacement of coronary arteries using multiple methods |
| K44.2 | Revision of replacement of coronary artery |
| K44.9 | Unspecified other replacement of coronary artery |
| K45.1 | Double anastomosis of mammary arteries to coronary arteries |
| K45.2 | Double anastomosis of thoracic arteries to coronary arteries NEC |
| K45.3 | Anastomosis of mammary artery to left anterior descending coronary artery |
| K45.4 | Anastomosis of mammary artery to coronary artery NEC |
| K45.5 | Anastomosis of thoracic artery to coronary artery NEC |
| K45.6 | Revision of connection of thoracic artery to coronary artery |
| K45.8 | Other specified connection of thoracic artery to coronary artery |
| K45.9 | Unspecified connection of thoracic artery to coronary artery |
| K49.1 | Percutaneous transluminal balloon angioplasty of one coronary artery |
| K49.2 | Percutaneous transluminal balloon angioplasty of multiple coronary arteries |
| K49.3 | Percutaneous transluminal balloon angioplasty of bypass graft of coronary artery |
| K49.4 | Percutaneous transluminal cutting balloon angioplasty of coronary artery |
| K49.8 | Other specified transluminal balloon angioplasty of coronary artery |
| K49.9 | Unspecified transluminal balloon angioplasty of coronary artery |
| K50.1 | Percutaneous transluminal laser coronary angioplasty |
| K50.2 | Percutaneous transluminal coronary thrombolysis using streptokinase |
| K50.3 | Percutaneous transluminal injection of therapeutic substance into coronary artery NEC |
| K50.4 | Percutaneous transluminal atherectomy of coronary artery |
| K50.8 | Other specified other therapeutic transluminal operations on coronary artery |
| K50.9 | Unspecified other therapeutic transluminal operations on coronary artery |
| K75.1 | Percutaneous transluminal balloon angioplasty and insertion of 1-2 drug-eluting stents into coronary artery |
| K75.2 | Percutaneous transluminal balloon angioplasty and insertion of 3 or more drug-eluting stents into coronary artery |
| K75.3 | Percutaneous transluminal balloon angioplasty and insertion of 1-2 stents into coronary artery |
| K75.4 | Percutaneous transluminal balloon angioplasty and insertion of 3 or more stents into coronary artery NEC |
| K75.8 | Other specified percutaneous transluminal balloon angioplasty and insertion of stent into coronary artery |
| K75.9 | Unspecified percutaneous transluminal balloon angioplasty and insertion of stent into coronary artery |
| **Myocardial infarction** |  |
| ICD10 codes | Definition |
| I21.0 | Acute transmural myocardial infarction of anterior wall |
| I21.1 | Acute transmural myocardial infarction of inferior wall |
| I21.2 | Acute transmural myocardial infarction of other sites |
| I21.3 | Acute transmural myocardial infarction of unspecified site |
| I21.4 | Acute subendocardial myocardial infarction |
| I21.9 | Acute myocardial infarction, unspecified |
| I22.0 | Subsequent myocardial infarction of anterior wall |
| I22.1 | Subsequent myocardial infarction of inferior wall |
| I22.8 | Subsequent myocardial infarction of other sites |
| I22.9 | Subsequent myocardial infarction of unspecified site |
| I23.0 | Hemopericardium as current complication following acute myocardial infarction |
| I23.1 | Atrial septal defect as current complication following acute myocardial infarction |
| I23.2 | Ventricular septal defect as current complication following acute myocardial infarction |
| I23.3 | Rupture of cardiac wall without hemopericardium as current complication following acute myocardial infarction |
| I23.4 | Rupture of chordae tendineae as current complication following acute myocardial infarction |
| I23.5 | Rupture of papillary muscle as current complication following acute myocardial infarction |
| I23.6 | Thrombosis of atrium, auricular appendage and ventricle as current complications following acute myocardial infarction |
| I23.8 | Other current complications following acute myocardial infarction |
| ICD9 codes | Definition |
| 4109 | Acute myocardial infarction |
| **Bundle branch block/fascicular block** |  |
| ICD10 codes | Definition |
| I444 | Left anterior fascicular block |
| I445 | Left posterior fascicular block |
| I446 | Other and unspecified fascicular block |
| I447 | Left bundle-branch block, unspecified |
| ICD9 codes | Definition |
| 4263 | Other left bundle branch block |
| 4264 | Right bundle branch block |
| 4265 | Bundle branch block, unspecified |
| **Atrioventricular block/Pacemaker** |  |
| ICD10 codes | Definition |
| Z450 | Adjustment and management of cardiac pacemaker |
| I441 | Atrioventricular block, second degree |
| I442 | Atrioventricular block, complete |
| ICD9 codes | Definition |
| 4260 | Atrioventricular block, complete |
| OPCS4 | Definition |
| K60.1 | Implantation of intravenous cardiac pacemaker system NEC |
| K60.2 | Resiting of lead of intravenous cardiac pacemaker system |
| K60.3 | Renewal of intravenous cardiac pacemaker system |
| K60.4 | Removal of intravenous cardiac pacemaker system |
| K60.5 | Implantation of intravenous single chamber cardiac pacemaker system |
| K60.6 | Implantation of intravenous dual chamber cardiac pacemaker system |
| K60.7 | Implantation of intravenous biventricular cardiac pacemaker system |
| K60.8 | Other specified cardiac pacemaker system introduced through vein |
| K60.9 | Unspecified cardiac pacemaker system introduced through vein |
| K61.1 | Implantation of cardiac pacemaker system NEC |
| K61.2 | Resiting of lead of cardiac pacemaker system NEC |
| K61.3 | Renewal of cardiac pacemaker system NEC |
| K61.4 | Removal of cardiac pacemaker system NEC |
| K61.5 | Implantation of single chamber cardiac pacemaker system |
| K61.6 | Implantation of dual chamber cardiac pacemaker system |
| K61.7 | Implantation of biventricular cardiac pacemaker system |
| K61.8 | Other specified other cardiac pacemaker system |
| K61.9 | Unspecified other cardiac pacemaker system |
| **Heart Failure** |  |
| ICD10 codes | Definition |
| I13.0 | Hypertensive heart and renal disease with both (congestive) heart failure |
| I13.2 | Hypertensive heart and renal disease with both (congestive) heart failure and renal failure |
| I50 | Heart failure |
| I50.0 | Congestive heart failure |
| I50.1 | Left ventricular failure |
| I50.9 | Heart failure, unspecified |
| ICD9 codes | Definition |
| 4280 | Congestive heart failure |
| 4281 | Left heart failure |
| 4289 | Heart failure, unspecified |
| OPCS4 | Definition |
| K59.6 | Implantation of cardioverter defibrillator using three electrode leads |
| K61.7 | Implantation of biventricular cardiac pacemaker system |
| K60.7 | Implantation of intravenous biventricular cardiac pacemaker system |
| **Non-ischaemic cardiomyopathy** |  |
| ICD10 codes | Definition |
| I11.0 | Hypertensive heart disease with (congestive) heart failure |
| I11.9 | Hypertensive heart disease without (congestive) heart failure |
| I42.0 | Dilated cardiomyopathy |
| I42.1 | Obstructive hypertrophic cardiomyopathy |
| I42.2 | Other hypertrophic cardiomyopathy |
| I42.3 | Endomyocardial (eosinophilic) disease |
| I42.4 | Endocardial fibroelastosis |
| I42.5 | Other restrictive cardiomyopathy |
| I42.6 | Alcoholic cardiomyopathy |
| I42.7 | Cardiomyopathy due to drugs and other external agents |
| I42.8 | Other cardiomyopathies |
| I42.9 | Cardiomyopathy, unspecified |
| I43.0 | Cardiomyopathy in infectious and parasitic diseases classified elsewhere |
| I43.1 | Cardiomyopathy in metabolic diseases |
| I43.2 | Cardiomyopathy in nutritional diseases |
| I43.8 | Cardiomyopathy in other diseases classified elsewhere |
| ICD9 codes | Definition |
| 4254 | Dilated cardiomyopathy |
| 42511 | Obstructive hypertrophic cardiomyopathy |
| 42518 | Other hypertrophic cardiomyopathy |
| 425 | Endomyocardial (eosinophilic) disease |
| 4253 | Endocardial fibroelastosis |
| 4254 | Other restrictive cardiomyopathy |
| 4255 | Alcoholic cardiomyopathy |
| 4259 | Cardiomyopathy due to drugs and other external agents |
| 4252/4254 | Other cardiomyopathies |
| 4254/4259 | Cardiomyopathy, unspecified |
| 4258 | Cardiomyopathy in diseases classified elsewhere |
| 4257 | Cardiomyopathy in metabolic diseases |
| 4257 | Cardiomyopathy in nutritional diseases |
| 4258 | Cardiomyopathy in other diseases classified elsewhere |
| **Ventricular**<br>**arrhythmia** |  |
| ICD10 codes | Definition |
| I47.2 | Ventricular tachycardia |
| I49.0 | Ventricular fibrillation and flutter |
| ICD9 codes | Definition |
| 4272 | Paroxysmal ventricular tachycardia |
| 4274 | Ventricular fibrillation and flutter |

## Supplementary Note 3 – Study information for sudden cardiac death cohorts

| Study | Study full name | Study reference (PMID) | Imputation panel | No. variants in PRS | Mean age (years) | Age range | % female | No. cases | No. controls | Definition of SCD |
| --- | --- | --- | --- | --- | --- | --- | --- | --- | --- | --- |
| ARIC | Atherosclerosis Risk in Communities Study | 2646917 | Affy 6.0 Array, TOPMed | QT=152; JT=135; QRS=105 | 54.3 | 44-66 | 53.0 | 230 | 8656 | ARIC is a prospective community-based study of cardiovascular disease containing SCD cases which were reviewed by a committee of physicians to determine the likelihood of sudden arrhythmic death. SCD was defined as a sudden pulseless condition presumed due to a ventricular tachyarrhythmia in a previously stable individual without evidence of a non-cardiac cause of cardiac arrest. All cardiac arrest events occurred out of the hospital or in the emergency room.<br>Further study information is available in the GWAS meta-analysis study sections. |
| CABS | Cardiac Arrest Blood Study | 16618831, 30169657 | Affymetrix Axiom | QT=148; JT=132; QRS=99 | 67.1 | 20-101 | 22.5 | 2165 | 2430 | The Cardiac Arrest Blood Study (CABS) is a large community-based repository of data and specimens from adult out-of-hospital cardiac arrest patients who were attended by paramedics in Seattle and King County, Washington. Clinical data was reviewed along with the autopsy reports when available, to determine likelihood of a primary arrhythmic event. Sudden cardiac arrest was defined as a sudden pulseless condition presumed due to a cardiac arrhythmia in the absence of any evidence of a non-cardiac cause of arrest. |
| FinGesture | Finnish Genetic Study for Arrhythmic Events | N/A | Affy 6.0 Array + GSA Array, TOPMed | QT=138; JT=122; QRS=99 | 62.0 | 25-85 | 19.7 | 1586 | N/A | The Fingesture study, started in 1998, aimed to collect consecutive individuals with of out-of-hospital sudden death from a defined geographic area, Oulu University Hospital District in northern Finland. All individuals with sudden death were autopsied at the Department of Forensic Medicine, University of Oulu, Oulu, Finland. Individuals with SCD were defined as those with a witnessed sudden death within 6 hours of the onset of the symptoms or within 24 hours of the time that the individual was last seen alive in a normal state of health. The underlying pathologies were divided into 3 categories: (1) ischemic, (2) nonischemic, and (3) other disease. The individuals with ischemic SCD included individuals with evidence of a coronary complication, defined as a fresh intracoronary thrombus, plaque rupture or erosion, intraplaque hemorrhage, or critical coronary stenosis (>75%) in the main coronary artery. The individuals with nonischemic SCD included individuals with the following conditions: hypertrophy caused by hypertension, valve disease, cardiomyopathy attributable to alcohol use, dilated cardiomyopathy, hypertrophic obstructive cardiomyopathy, cardiomyopathy caused by obesity, arrhythmogenic right ventricular cardiomyopathy, and primary myocardial fibrosis. The “other” individuals with SCD included individuals with the following conditions: myocarditis, cardiac anomaly, and individuals with a normal autopsy (eg, individuals with a channelopathy). |
| NFBC1966 | Northern Finland Birth Cohort of 1966 | 19060910 | GSA Array, TOPMed | QT=139;<br>JT=122;<br>QRS=99 | 31.0 | 31 (one year birth cohort) | 53.5 | N/A | 761 | This study was used as a population control group for FinGesture study analyses. The NFBC1966 (Northern Finland Birth Cohort) study is the product of a project initiated in the 1960s to examine risk factors involved in preterm birth and intrauterine growth restriction, and the consequences of these early adverse outcomes on subsequent morbidity. The NFBC1966 cohort comprised 12 068 mothers and 12 231 children with an expected date of birth in 1966 within the province of Oulu, Finland. Study samples consisted of DNA extracted from the blood of the offspring at their 31-year follow-up visit. |

No.variants included in PRS; Number of lead variants at loci in the European-ancestry meta-analysis for each ECG trait, which passed the imputation quality threshold ( $R_{sq} > 0.8$ ) and were thus included in the model

## Supplementary Note 4 – Study Acknowledgements

### ARIC - Atherosclerosis Risk in Communities

The authors thank the staff and participants of the ARIC study for their important contributions.

### BioMe - The IPM Bio*Me* Biobank

The Mount Sinai BioMe Biobank is supported by The Andrea and Charles Bronfman Philanthropies.

### BRIGHT - British Genetics of Hypertension

The BRIGHT study is extremely grateful to all the patients who participated in the study and the BRIGHT nursing team. This work forms part of the research program of the National Institutes of Health Research (NIHR Cardiovascular Biomedical Research) Cardiovascular Biomedical Unit at Barts and The London, QMUL. P.B.M. wishes to acknowledge the NIHR Cardiovascular Biomedical Research Unit at Barts and The London, Queen Mary University of London, UK for support.

### CHRIS - The Cooperative Health Research in South Tyrol study

The CHRIS study is a collaborative effort between the Center for Biomedicine of the European Academy of Bolzano/Bozen (EURAC) and the Healthcare System of the Autonomous Province of Bolzano (Südtiroler Sanitätsbetrieb/Azienda Sanitaria dell’Alto Adige). The CHRIS Study is affiliated to the “German National Cohort” (Germany) and is indebted with the investigators of this study for their support in the study protocol definition. Full acknowledgements for the CHRIS study are reported here: http://translational-medicine.biomedcentral.com/articles/10.1186/s12967-015-0704-9#Declarations.

### ERF - Erasmus Rucphen Family Study

We are grateful to all study participants and their relatives, general practitioners and neurologists for their contributions to the ERF study and to P Veraart for her help in genealogy, J Vergeer for the supervision of the laboratory work and P Snijders for his help in data collection.

### FINCAVAS - Finnish Cardiovascular Study

The authors thank the staff of the Department of Clinical Physiology for collecting the exercise test data.

### GAPP - Genetic and phenotypic determinants of blood pressure and other cardiovascular risk factors

We thank the GAPP staff and all GAPP study participants for their important contributions

### GESUS - The Danish General Suburban Population Study GS20 - Generation Scotland: Scottish Family Health Study

We are grateful to all the families who took part, the general practitioners and the Scottish School of Primary Care for their help in recruiting them, and the whole Generation Scotland team, which includes interviewers, computer and laboratory technicians, clerical workers, research scientists, volunteers, managers, receptionists, healthcare assistants and nurses.

### HCHS/SOL - Hispanic Community Health Survey/Study of Latinos

Hispanic Community Health Study/Study of Latinos (HCHS/SOL): We thank the participants and staff of the HCHS/SOL study for their contributions to this study. The baseline examination of HCHS/SOL was carried out as a collaborative study supported by contracts from the National Heart, Lung, and Blood Institute (NHLBI) to the University of North Carolina (N01-HC65233), University of Miami (N01-HC65234), Albert Einstein College of Medicine (N01-HC65235), Northwestern University (N01-HC65236) and San Diego State University (N01-HC65237).

### Health ABC - Health, Aging, and Body Composition Study

This research was supported by NIA contracts N01AG62101, N01AG62103, and N01AG62106. The genome-wide association study was funded by NIA grant 1R01AG032098-01A1 to Wake Forest University Health Sciences and genotyping services were provided by the Center for Inherited Disease Research (CIDR). CIDR is fully funded through a federal contract from the National Institutes of Health to The Johns Hopkins University, contract number HHSN268200782096C. This research was supported in part by the Intramural Research Program of the NIH, National Institute on Aging.

### INGI-CAR - INGI-CARLANTINO / INGI-FVG - INGI- Friuli Venezia Giulia

We are very grateful to the municipal administrators for their collaboration on the project and for logistic support. We would like to thank all participants to this study.

### INTER99 - Inter99

The Inter99 was initiated by Torben Jørgensen (PI), Knut Borch-Johnsen (co-PI), Hans Ibsen and Troels F. Thomsen. The steering committee comprises the former two and Charlotta Pisinger.

### JHS - Jackson Heart Study

We thank the Jackson Heart Study (JHS) participants and staff for their contributions to this work.

### KORA F3/S4 - Kooperative Gesundheitsforschung in der Region Augsburg F3/S4

The KORA study was initiated and financed by the Helmholtz Zentrum München – German Research Center for Environmental Health, which is funded by the German Federal Ministry of Education and Research (BMBF) and by the State of Bavaria. Furthermore, KORA research was supported within the Munich Center of Health Sciences (MC-Health), Ludwig-Maximilians-Universität, as part of LMUinnovativ.

### LIFELINES - LifeLines, a three-generation cohort study and biobank

We thank U. Bultmann (1), J.M. Geleijnse (2), P. van der Harst (3), S. Mulder (4), J.G.M. Rosmalen (5), E.F.C. van Rossum (6), H.A. Smit (7), M. Swertz (8), E.A.L.M. Verhagen (9). (1) Department of Social Medicine, University Medical Center Groningen, University of Groningen, The Netherlands (2) Department of Human Nutrition, Wageningen University, The Netherlands (3) Department of Cardiology, University Medical Center Groningen, University of Groningen, The Netherlands (4) Lifelines Cohort Study, The Netherlands (5) Interdisciplinary Center of Psychopathology of Emotion Regulation (ICPE), Department of Psychiatry, University Medical Center Groningen, University of Groningen, The Netherlands (6) Department of Endocrinology, Erasmus Medical Center, Rotterdam, The Netherlands (7) Department of Public Health, University Medical Center Utrecht, The Netherlands (8) Department of Genetics, University Medical Center Groningen, University of Groningen, The Netherlands (9) Department of Public and Occupational Health, VU Medical Center, Amsterdam, The Netherlands

### NEO - Netherlands Epidemiology of Obesity

The authors of the NEO study thank all individuals who participated in the Netherlands Epidemiology in Obesity study, all participating general practitioners for inviting eligible participants and all research nurses for collection of the data. We thank the NEO study group, Pat van Beelen, Petra Noordijk and Ingeborg de Jonge for the coordination, lab and data management of the NEO study. We also thank Arie Maan for the analyses of the electrocardiograms.

### OOA - Old Order Amish

We gratefully thank our Amish community and research volunteers for their long-standing partnership in research, and acknowledge the dedication of our Amish liaisons, field workers and the Amish Research Clinic staff, without which these studies would not have been possible.

### ORCADES - Orkney Complex Disease Study

DNA extractions were performed at the Wellcome Trust Clinical Research Facility in Edinburgh. We would like to acknowledge the invaluable contributions of the research nurses in Orkney, the administrative team in Edinburgh and the people of Orkney.

### PIVUS - Prospective Investigation of the Vasculature of Uppsala Seniors

DNA extractions were performed at the Wellcome Trust Clinical Research Facility in Edinburgh. We would like to acknowledge the invaluable contributions of the research nurses in Orkney, the administrative team in Edinburgh and the people of Orkney.

### PROSPER - PROSpective study of pravastatin in the elderly at Risk for vascular disease

The PROSPER study was supported by an investigator initiated grant obtained from Bristol-Myers Squibb. J.Wouter Jukema is an Established Clinical Investigator of the Netherlands Heart Foundation (grant 2001 D 032). Support for genotyping was provided by the seventh framework program of the European commission (grant 223004) and by the Netherlands Genomics Initiative (Netherlands Consortium for Healthy Aging grant 050-060-810).

### RS - Rotterdam Study

We thank Pascal Arp, Mila Jhamai, Dr Michael Moorhouse, Marijn Verkerk, and Sander Bervoets for their help in creating the GWAS database. The authors are very grateful to the participants and staff from the Rotterdam Study, the participating general practitioners and the pharmacists.

### SardiNIA - SardiNIA Project

We thank the many volunteers who generously participated in this study, the Mayors and citizens of the Sardinian towns involved, the head of the Public Health Unit ASL4, and the province of Ogliastra for their volunteerism and cooperation. In addition, we are grateful to the Mayor and the administration in Lanusei for providing and furnishing the clinic site. We are grateful to the physicians Angelo Scuteri, Marco Orrù, Maria Grazia Pilia, Liana Ferreli, Francesco Loi, nurses Paola Loi, Monica Lai and Anna Cau who carried out participant physical exams; the recruitment personnel Susanna Murino; and Mariano Dei, Sandra Lai, Andrea Maschio, Fabio Busonero for genotyping.

### SHIP and SHIP-TREND - Study of Health in Pomerania

SHIP is part of the Community Medicine Research Network of the University Medicine Greifswald, which is supported by the German Federal State of Mecklenburg-West Pomerania.

### UK Biobank - UK Biobank Study

This research has been conducted using the UK Biobank Resource (application 8256 - Understanding genetic influences in the response of the cardiac electrical system to exercise). This work forms part of the research program of the National Institutes of Health Research (NIHR Cardiovascular Biomedical Research) Cardiovascular Biomedical Centre at Barts and The London, QMUL. PD Lambiase acknowledges support from the UCLH Biomedicine NIHR. JR acknowledges support from the People Programme (Marie Curie Actions) of the European Union’s Seventh Framework Programme (FP7/2007/2013) under REA grant agreement 608765

### VIKING - Viking Health Study

DNA extractions and genotyping were performed at the Edinburgh Clinical Research Facility, University of Edinburgh. We would like to acknowledge the invaluable contributions of the research nurses in Shetland, the administrative team in Edinburgh and the people of Shetland.

### WHI - Women’s Health Initiative

The WHI program is funded by the National Heart, Lung, and Blood Institute, National Institutes of Health. The authors thank the WHI investigators and staff for their dedication, and the study participants for making the program possible. A full listing of WHI investigators can be found at: https://www-whi-org.s3.us-west-2.amazonaws.com/wp-content/uploads/WHI-Investigator-Long-List.pdf.

### YFS - Young Finns Study

We thank the teams that collected data at all measurement time points; the persons who participated as both children and adults in these longitudinal studies; and biostatisticians Irina Lisinen, Johanna Ikonen, Noora Kartiosuo, Ville Aalto, and Jarno Kankaanranta for data management and statistical advice.

### Additional acknowledgements

This research utilized Queen Mary’s Apocrita HPC facility, supported by QMUL Research-IT. http://doi.org/10.5281/zenodo.438045

## Supplementary Note 5 – Study Funding

### ARIC - Atherosclerosis Risk in Communities

The Atherosclerosis Risk in Communities study has been funded in whole or in part with Federal funds from the National Heart, Lung, and Blood Institute, National Institutes of Health, Department of Health and Human Services, under Contract nos. (HHSN268201700001I, HHSN268201700002I, HHSN268201700003I, HHSN268201700004I, HHSN268201700005I). Funding was also supported by R01HL087641, R01HL059367 and R01HL086694; National Human Genome Research Institute contract U01HG004402; and National Institutes of Health contract HHSN268200625226C. Infrastructure was partly supported by Grant Number UL1RR025005, a component of the National Institutes of Health and NIH Roadmap for Medical Research. Alvaro Alonso is supported by an NHLBI award K24HL148521.

### Bambui - Brazilian Bambuí Cohort Study of Ageing

Brazilian Ministry of Health (DECIT/MS, EPIGEN-Brazil Project), Brazilian Ministry of Science and Technology (Financiadora de Estudos e Projetos (FINEP), Brazilian Conselho Nacional de Desenvolvimento Científico e Tecnologico (CNPq), Fundação de Amparo a Pesquisa do Estado de Minas Gerais (FAPEMIG). Antonio Luiz P Ribeiro is supported in part by CNPq (310679/2016-8 and 465518/2014-1) and by FAPEMIG (PPM-00428-17 and RED-00081-16). Eduardo Tarazona-Santos, Maria Fernanda Lima-Costa and Thiago P Leal are supported by Brazilian Conselho Nacional de Desenvolvimento Científico e Tecnologico (CNPq). Eduardo Tarazona-Santos and Maria Fernanda Lima-Costa are also supported by the Brazilian Ministry of Health (DECIT/MS, EPIGEN-Brazil Project), Brazilian Ministry of Science and Technology (Financiadora de Estudos e Projetos (FINEP) and Fundação de Amparo a Pesquisa do Estado de Minas Gerais (FAPEMIG).

### BioMe - The IPM Bio*Me* Biobank

Ruth Loos is supported by the National Institutes of Health (R01DK110113; R01DK107786; R01HL142302; R01DK124097)

### BRIGHT - British Genetics of Hypertension

This work was supported by the Medical Research Council (MRC) of Great Britain (grant number G9521010D) and the British Heart Foundation (grant number PG/02/128). William Jon Young is supported by an MRC grant MR/R017468/1. Patricia B Munroe is also supported by an MRC grant MR/N025083/1.

### Broad AF

Michael J Cutler is supported by funding from the Dell Loy Hansen Heart Foundation. M.Benjamin Shoemaker is supported by NIHK23HL127704

### CABS - Cardiac Arrest Blood Study

Rozenn Lemaitre reports funding from grant: AHA 19SFRN34830063.

### CAMP-MGH - MGH Cardiology and Metabolic Patient Cohort

Steven A Lubitz is supported by NIH grant 1R01HL139731 and American Heart Association 18SFRN34250007. Patrick T Ellinor was supported by grants from Bayer AG, the Fondation Leducq (14CVD01), the NIH (1RO1HL092577, R01HL128914, K24HL105780), and the American Heart Association (18SFRN34110082). Lu-Chen Weng is supported by the American Heart Association (18SFRN34110082). Victor Nauffal is funded by a T32 training grant from the National Institutes of Health (5T32HL007604-35)

### CHRIS - The Cooperative Health Research in South Tyrol study

The CHRIS study was funded by the Department of Innovation, Research, and University of the Autonomous Province of Bolzano-South Tyrol.

### CHS - Cardiovascular Health Study

Cardiovascular Health Study: This CHS research was supported by NHLBI contracts HHSN268200960009C, HHSN268201200036C, HHSN268200800007C, HHSN268201800001C, N01HC55222, N01HC85079, N01HC85080, N01HC85081, N01HC85082, N01HC85083, N01HC85086, 75N92021D00006; and NHLBI grants U01HL080295, R01HL085251, R01HL087652, R01HL105756, R01HL103612, R01HL120393, and U01HL130114 with additional contribution from the National Institute of Neurological Disorders and Stroke (NINDS). Additional support was provided through R01AG023629 from the National Institute on Aging (NIA). A full list of principal CHS investigators and institutions can be found at CHS-NHLBI.org. The provision of genotyping data was supported in part by the National Center for Advancing Translational Sciences, CTSI grant UL1TR001881, and the National Institute of Diabetes and Digestive and Kidney Disease Diabetes Research Center (DRC) grant DK063491 to the Southern California Diabetes Endocrinology Research Center. The content is solely the responsibility of the authors and does not necessarily represent the official views of the National Institutes of Health. Nona Sotoodehnia is supported by grants AHA19SFRN348300063, R01HL141989, Medic One Foundation, Laughlin Family.

### ERF - Erasmus Rucphen Family Study

The ERF study as a part of EUROSPAN (European Special Populations Research Network) was supported by European Commission FP6 STRP grant number 018947 (LSHG-CT-2006-01947) and also received funding from the European Community’s Seventh Framework Programme (FP7/2007- 2013)/grant agreement HEALTH-F4-2007-201413 by the European Commission under the programme “Quality of Life and Management of the Living Resources” of 5th Framework Programme (no. QLG2-CT-2002-01254). The ERF study was further supported by ENGAGE consortium and CMSB. High-throughput analysis of the ERF data was supported by joint grant from Netherlands Organisation for Scientific Research and the Russian Foundation for Basic Research (NWO-RFBR 047.017.043). Ulrich Schotten received funding from the Netherlands Heart Foundation (CVON2014-09, RACE V Reappraisal of Atrial Fibrillation: Interaction between hyperCoagulability, Electrical remodeling, and Vascular Destabilization in the Progression of AF), the European Union (ITN Network Personalize AF: Personalized Therapies for Atrial Fibrillation: a translational network, grant number 860974; MAESTRIA: Machine Learning Artificial Intelligence Early Detection Stroke Atrial Fibrillation, grant number 965286).

### FINCAVAS - Finnish Cardiovascular Study

The Finnish Cardiovascular Study (FINCAVAS) has been financially supported by the Competitive Research Funding of the Tampere University Hospital (Grant 9M048 and 9N035), the Finnish Cultural Foundation, the Finnish Foundation for Cardiovascular Research, the Emil Aaltonen Foundation, Finland, the Tampere Tuberculosis Foundation, EU Horizon 2020 (grant 755320 for TAXINOMISIS and grant 848146 for To Aition), and the Academy of Finland grant 322098.

### FinGesture – Finnish Genetic Study for Arrhythmic Events

This work was supported by the Juselius Foundation (Helsinki, Finland); the Council of Health of the Academy of Finland (Helsinki, Finland); the Montreal Heart Institute Foundation; Finnish Foundation for Cardiovascular Research (Helsinki, Finland); and Erkko Foundation (Helsinki, Finland).

### GAPP - Genetic and phenotypic determinants of blood pressure and other cardiovascular risk factors

The GAPP study was supported by the Liechtenstein Government, the Swiss Heart Foundation, the Swiss Society of Hypertension, the University of Basel, the University Hospital Basel, the Hanela Foundation, the Mach-Gaensslen Foundation, Schiller AG, and Novartis. Sébastien Thériault is supported by a Junior 1 Clinical Research Scholar award from the Fonds de Recherche du Québec-Santé (FRQS).

### GESUS - The Danish General Suburban Population Study

Jonas L Isaksen is supported by CACHET.

### GS20 - Generation Scotland: Scottish Family Health Study

Generation Scotland received core support from the Chief Scientist Office of the Scottish Government Health Directorates [CZD/16/6] and the Scottish Funding Council [HR03006]. Genotyping of the GS:SFHS samples was carried out by the Genetics Core Laboratory at the t Clinical Research Facility, University of Edinburgh, Scotland and was funded by the Medical Research Council UK and the Wellcome Trust (Wellcome Trust Strategic Award “STratifying Resilience and Depression Longitudinally” (STRADL) Reference 104036/Z/14/Z). C.H is supported by an MRC University Unit Programme Grant MC_UU_00007/10 (QTL in Health and Disease)

### HCHS/SOL - Hispanic Community Health Survey/Study of Latinos

This work was supported by funding from the National Heart, Lunch and Blood Institute (N01-HC65233, N01-HC65234, N01-HC65235, N01-HC65236, N01-HC65237, and T32HL7055). Christy Avery and Antoine Baldassari were supported by NIH grants R01HL142825, and U01HG007416. Dawood Darbar was supported by NIH grants R01HL138737 and T32HL139439.

### Health ABC - Health, Aging, and Body Composition Study

This research was supported by NIA contracts N01AG62101, N01AG62103, and N01AG62106. The genome-wide association study was funded by NIA grant 1R01AG032098-01A1 to Wake Forest University Health Sciences and genotyping services were provided by the Center for Inherited Disease Research (CIDR). CIDR is fully funded through a federal contract from the National Institutes of Health to The Johns Hopkins University, contract number HHSN268200782096C. This research was supported in part by the Intramural Research Program of the NIH, National Institute on Aging. Daniel S Evans is supported by NIH grant U24AG051129.

### INGI-CAR - INGI-CARLANTINO / INGI-FVG - INGI-Friuli Venezia Giulia

Italian Ministry of Health - RC 35/17; Italian Ministry of Education, University and Research, D70- RESRICGIROTTO to GG

### INTER99 - Inter99

The study was financially supported by research grants from the Danish Research Council, the Danish Centre for Health Technology Assessment, Novo Nordisk Inc., Research Foundation of Copenhagen County, Ministry of Internal Affairs and Health, the Danish Heart Foundation, the Danish Pharmaceutical Association, the Augustinus Foundation, the Ib Henriksen Foundation, the Becket Foundation, and the Danish Diabetes Association. Niels Grarup is supported by the Novo Nordisk Foundation (Grant number NNF18CC0034900).

### JHS - Jackson Heart Study

The JHS is supported by contracts HHSN268201800010, HHSN268201800011, HHSN268201800012, HHSN268201800013, HHSN268201800014, and HHSN268201800015 from the National Heart, Lung, and Blood Institute and the National Institute on Minority Health and Health Disparities. Dr. Wilson is supported by U54GM115428 from the National Institute of General Medical Sciences. Dr. Hao Mei is also supported by the CHARGE infrastructure grant (HL105756).

### LIFELINES - LifeLines, a three-generation cohort study and biobank

The LifeLines Cohort Study, and generation and management of GWAS genotype data for the LifeLines Cohort Study is supported by the Netherlands Organization of Scientific Research NWO (grant 175.010.2007.006), the Economic Structure Enhancing Fund (FES) of the Dutch government, the Ministry of Economic Affairs, the Ministry of Education, Culture and Science, the Ministry for Health, Welfare and Sports, the Northern Netherlands Collaboration of Provinces (SNN), the Province of Groningen, University Medical Center Groningen, the University of Groningen, Dutch Kidney Foundation and Dutch Diabetes Research Foundation. Jan-Walter Benjamins is funded by the Research Project CVON-AI (2018B017) financed by the PPP Allowance made available by Top Sector Life Sciences & Health to the Dutch Heart Foundation to stimulate public-private partnerships.

### MESA - Multi-Ethnic Study of Atherosclerosis

MESA and the MESA SHARe project are conducted and supported by the National Heart, Lung, and Blood Institute (NHLBI) in collaboration with MESA investigators. Support for MESA is provided by contracts 75N92020D00001, HHSN268201500003I, N01-HC-95159, 75N92020D00005, N01-HC-95160, 75N92020D00002, N01-HC-95161, 75N92020D00003, N01-HC-95162, 75N92020D00006, N01-HC-95163, 75N92020D00004, N01-HC-95164, 75N92020D00007, N01-HC-95165, N01-HC-95166, N01-HC-95167, N01-HC-95168, N01-HC-95169, UL1-TR-000040, UL1-TR-001079, UL1-TR-001420. Funding for SHARe genotyping was provided by NHLBI Contract N02-HL-64278. Genotyping was performed at Affymetrix (Santa Clara, California, USA) and the Broad Institute of Harvard and MIT (Boston, Massachusetts, USA) using the Affymetrix Genome-Wide Human SNP Array 6.0. Also supported in part by the NHLBI contracts R01HL151855 and R01HL146860. Also supported in part by the National Center for Advancing Translational Sciences, CTSI grant UL1TR001881, and the National Institute of Diabetes and Digestive and Kidney Disease Diabetes Research Center (DRC) grant DK063491 to the Southern California Diabetes Endocrinology Research Center. Infrastructure for the CHARGE Consortium is supported in part by the National Heart, Lung, and Blood Institute (NHLBI) grant R01HL105756.

### NEO - Netherlands Epidemiology of Obesity

The genotyping in the NEO study was supported by the Centre National de Génotypage (Paris, France), headed by Jean-Francois Deleuze. The NEO study is supported by the participating Departments, the Division and the Board of Directors of the Leiden University Medical Center, and by the Leiden University, Research Profile Area Vascular and Regenerative Medicine. Dennis Mook-Kanamori is supported by Dutch Science Organization (ZonMW-VENI Grant 916.14.023).

### NFBC1966 – Northern Finland Birth Cohort of 1966

The NFBC1966 Study is conducted and supported by the National Heart, Lung, and Blood Institute (NHLBI [grant number: 5R01HL087679-02] through the STAMPEED program [genotyping, grant number: 1RL1MH083268-01]), in collaboration with the Broad Institute, UCLA, University of Oulu, and the National Institute for Health and Welfare in Finland. The NFBC1966 has received core funding for data generation and curation from the Academy of Finland (project grants 104781, 120315, 129269, 1114194, 24300796, 85547, 285547 (EGEA)), University Hospital Oulu, Finland (75617), ERDF European Regional Development Fund Grant no. 539/2010 A31592 and the EU H2020--PHC-2014 DynaHEALTH action (grant no. 633595).

### OOA - Old Order Amish

The Amish studies are supported by grants and contracts from the NIH, including R01 AG18728, R01 HL088119, U01 GM074518, U01 HL072515, U01 HL84756, U01 HL137181, R01 DK54261,the University of Maryland General Clinical Research Center, grant M01 RR 16500, and the Mid-Atlantic Nutrition Obesity Research Center grant P30 DK72488, the Baltimore Diabetes Research and Training Center grant P60DK79637. May E Montasser receives funding from Regeneron Pharmaceutical unrelated to this work.

### ORCADES - Orkney Complex Disease Study

The Orkney Complex Disease Study (ORCADES) was supported by the Chief Scientist Office of the Scottish Government (CZB/4/276, CZB/4/710), a Royal Society URF to J.F.W., the MRC Human Genetics Unit quinquennial programme “QTL in Health and Disease”, Arthritis Research UK and the European Union framework program 6 EUROSPAN project (contract no. LSHG-CT-2006-018947). James F Wilson acknowledges support from the MRC Human Genetics Unit programme grant, “Quantitative traits in health and disease” (U. MC_UU_00007/10). Linda Repetto is funded by a University of Edinburgh studentship. Pau Navarro is supported by the MRC Human Genetics Unit programme grant, “Quantitative traits in health and disease” (U. MC_UU_00007/10). Xia Shen was in receipt of a Starting Grant (2017-02543) from the Swedish Research Council (Vetenskaprådet).

### PIVUS - Prospective Investigation of the Vasculature of Uppsala Seniors

PIVUS was supported by Knut and Alice Wallenberg Foundation (Wallenberg Academy Fellow), European Research Council (ERC Starting Grant), Swedish Diabetes Foundation (2013-024), Swedish Research Council (2012-1397, 2012-1727, and 2012-2215), Marianne and Marcus Wallenberg Foundation, County Council of Dalarna, Dalarna University, and Swedish Heart-Lung Foundation (20120197). Genotyping was funded by the Wellcome Trust under awards WT064890 and WT086596. Analysis of genetic data was funded by the Wellcome Trust under awards WT098017 and WT090532. Lars Lind is funded by Uppsala University Hospital, Uppsala, Sweden.

### PREVEND - Prevention of REnal and Vascular ENd stage Disease

PREVEND genetics is supported by the Dutch Kidney Foundation (Grant E033), the EU project grant GENECURE (FP-6 LSHM CT 2006 037697), the National Institutes of Health (grant 2R01LM010098), The Netherlands organization for health research and development (NWO-Groot grant 175.010.2007.006, NWO VENI grant 916.761.70, ZonMw grant 90.700.441), and the Dutch Inter University Cardiology Institute Netherlands (ICIN). N Verweij was supported by NWO VENI (016.186.125). Jan-Walter Benjamins is funded by the Research Project CVON-AI (2018B017) financed by the PPP Allowance made available by Top Sector Life Sciences & Health to the Dutch Heart Foundation to stimulate public-private partnerships.

### RS - Rotterdam Study

The Rotterdam Study is supported by the Erasmus Medical Center and Erasmus University Rotterdam; the Netherlands Organization for Scientific Research (NWO); the Netherlands Organization for Health Research and Development (ZonMw); the Research Institute for Diseases in the Elderly (RIDE); the Netherlands Heart Foundation; the Ministry of Education, Culture and Science; the Ministry of Health Welfare and Sports; the European Commission; and the Municipality of Rotterdam. Support for genotyping was provided by the Netherlands Organisation of Scientific Research NWO Investments (nr. 175.010.2005.011, 911-03-012), the Research Institute for Diseases in the Elderly (014-93-015; RIDE2), the Netherlands Genomics Initiative (NGI)/Netherlands Consortium for Healthy Aging (NCHA) project nr. 050-060-810. Jacqueline Witteman wais supported by NWO grant (vici, 918-76-619). Abbas Dehghan is supported by NWO grant (veni, 916.12.154) and the EUR Fellowship. O.H. Franco works in ErasmusAGE, a center for aging research across the life course funded by Nestlé Nutrition (Nestec Ltd.) and Metagenics Inc. Nestlé Nutrition (Nestec Ltd.) and Metagenics Inc. had no role in design and conduct of the study; collection, management, analysis, and interpretation of the data; and preparation, review or approval of the manuscript. The GWA study was funded by the Netherlands Organisation of Scientific Research NWO Investments (nr. 175.010.2005.011, 911-03-012), the Research Institute for Diseases in the Elderly (014-93-015; RIDE2), the Netherlands Genomics Initiative (NGI)/Netherlands Consortium for Healthy Aging (NCHA) project nr. 050-060-810.

### SardiNIA - SardiNIA Project

This study was funded in part by the National Institutes of Health (National Institute on Aging, National Heart Lung and Blood Institute, and National Human Genome Research Institute). This research was supported by the Intramural Research Program of the NIH, National Institute on Aging, with contracts N01-AG-1-2109 and HHSN271201100005C and by Sardinian Autonomous Region (L.R. no. 7/2009) grant cRP3-154.

### SHIP-START and SHIP-TREND - Study of Health in Pomerania

SHIP is supported by the German Federal Ministry of Education and Research (Bundesministerium für Bildung und Forschung (BMBF); grants 01ZZ9603, 01ZZ0103, and 01ZZ0403) and the German Research Foundation (Deutsche Forschungsgemeinschaft (DFG); grant GR 1912/5-1). SHIP-START and SHIP-TREND are part of the Community Medicine Research net (CMR) of the University of Greifswald which is funded by the BMBF as well as the Ministry for Education, Science and Culture and the Ministry of Labor, Equal Opportunities, and Social Affairs of the Federal State of Mecklenburg-West Pomerania. The CMR encompasses several research projects that share data from SHIP. Genome-wide data have been supported by a joint grant from Siemens Healthcare, Erlangen, Germany and the Federal State of Mecklenburg-West Pomerania.

### TWINSUK - TWINSUK

TwinsUK receives funding from the Wellcome Trust (212904/Z/18/Z), Medical Research Council (AIMHY; MR/M016560/1) and European Union (H2020 contract #733100). TwinsUK is supported by the National Institute for Health Research (NIHR)-funded BioResource, Clinical Research Facility and Biomedical Research Centre based at Guy’s and St Thomas’ NHS Foundation Trust in partnership with King’s College London. This work was supported by the British Heart Foundation (grant number PG/12/38/29615). Oliver Hines is supported by a Medical Research Council DTP Studentship. Massimo Mangino is supported by the National Institute for Health Research (NIHR)-funded BioResource, Clinical Research Facility and Biomedical Research Centre based at Guy’s and St Thomas’ NHS Foundation Trust in partnership with King’s College London.

### UK Biobank - UK Biobank Study

This work is supported by Medical Research Council grant MR/N025083/1. William Jon Young is supported by an MRC grant MR/R017468/1. Julia Ramirez is funded by European Union’s Horizon 2020 research and innovation programme under the Marie Sklodowska-Curie grant agreement No.786833. Pier D Lambiase is supported by UCL/UCLH Biomedicine NIHR, Barts Heart Centre Biomedical Research Centre. Patricia B Munroe, Pier D Lambiase and Andrew Tinker acknowledge the NIHR Cardiovascular Biomedical Centre at Barts and The London, Queen Mary University of London

### VIKING - Viking Health Study

The Viking Health Study – Shetland (VIKING) was supported by the MRC Human Genetics Unit quinquennial programme grant “QTL in Health and Disease”. Linda Repetto is supported by a University of Edinburgh studentship. James F Wilson acknowledges support from the MRC Human Genetics Unit programme grant, “Quantitative traits in health and disease” (U. MC_UU_00007/10).

### WHI - Women’s Health Initiative

The WHI program is funded by the National Heart, Lung, and Blood Institute, National Institutes of Health, U.S. Department of Health and Human Services through contracts HHSN268201600018C, HHSN268201600001C, HHSN268201600002C, HHSN268201600003C, and HHSN268201600004C. Scientific Computing Infrastructure at the Fred Hutchinson Cancer Research Center is funded by ORIP grant S10OD028685. The Modification of PM-Mediated Arrhythmogenesis in Populations (WHI-MOPMAP) was funded by the NIEHS (R01-ES017794, Whitsel).

### YFS - Young Finns Study

The Young Finns Study has been financially supported by the Academy of Finland: grants 322098, 286284, 134309 (Eye), 126925, 121584, 124282, 129378 (Salve), 117787 (Gendi), and 41071 (Skidi); the Social Insurance Institution of Finland; Competitive State Research Financing of the Expert Responsibility area of Kuopio, Tampere and Turku University Hospitals (grant X51001); Juho Vainio Foundation; Paavo Nurmi Foundation; Finnish Foundation for Cardiovascular Research ; Finnish Cultural Foundation; The Sigrid Juselius Foundation; Tampere Tuberculosis Foundation; Emil Aaltonen Foundation; Yrjö Jahnsson Foundation; Signe and Ane Gyllenberg Foundation; Diabetes Research Foundation of Finnish Diabetes Association; EU Horizon 2020 (grant 755320 for TAXINOMISIS and grant 848146 for To Aition); European Research Council (grant 742927 for MULTIEPIGEN project); Tampere University Hospital Supporting Foundation and Finnish Society of Clinical Chemistry.

## References

1. Krittayaphong, R. et al. Electrocardiographic predictors of cardiovascular events in patients at high cardiovascular risk: a multicenter study. J Geriatr Cardiol 16, 630–638 (2019).

2. Niemeijer, M.N., van den Berg, M.E., Eijgelsheim, M., Rijnbeek, P.R. & Stricker, B.H. Pharmacogenetics of Drug-Induced QT Interval Prolongation: An Update. Drug Saf 38, 855–67 (2015).

3. Schwartz, P.J., Crotti, L. & Insolia, R. Long-QT syndrome: from genetics to management. Circ Arrhythm Electrophysiol 5, 868–77 (2012).

4. Straus, S.M. et al. Prolonged QTc interval and risk of sudden cardiac death in a population of older adults. J Am Coll Cardiol 47, 362–7 (2006).

5. Zhang, Y. et al. Electrocardiographic QT interval and mortality: a meta-analysis. Epidemiology 22, 660–70 (2011).

6. Tester, D.J. & Ackerman, M.J. Genetics of long QT syndrome. Methodist Debakey Cardiovasc J 10, 29–33 (2014).

7. Lahrouchi, N. et al. Transethnic Genome-Wide Association Study Provides Insights in the Genetic Architecture and Heritability of Long QT Syndrome. Circulation 142, 324–338 (2020).

8. Jamshidi, Y., Nolte, I.M., Spector, T.D. & Snieder, H. Novel genes for QTc interval. How much heritability is explained, and how much is left to find? Genome Med 2, 35 (2010).

9. Wilde, A.A.M., Amin, A.S. & Postema, P.G. Diagnosis, management and therapeutic strategies for congenital long QT syndrome. Heart (2021).

10. Crow, R.S., Hannan, P.J. & Folsom, A.R. Prognostic significance of corrected QT and corrected JT interval for incident coronary heart disease in a general population sample stratified by presence or absence of wide QRS complex: the ARIC Study with 13 years of follow-up. Circulation 108, 1985–9 (2003).

11. Bihlmeyer, N.A. et al. ExomeChip-Wide Analysis of 95 626 Individuals Identifies 10 Novel Loci Associated With QT and JT Intervals. Circ Genom Precis Med 11, e001758 (2018).

12. Arking, D.E. et al. Genetic association study of QT interval highlights role for calcium signaling pathways in myocardial repolarization. Nat Genet 46, 826–36 (2014).

13. Sotoodehnia, N. et al. Common variants in 22 loci are associated with QRS duration and cardiac ventricular conduction. Nat Genet 42, 1068–76 (2010).

14. Desplantez, T., Dupont, E., Severs, N.J. & Weingart, R. Gap junction channels and cardiac impulse propagation. J Membr Biol 218, 13–28 (2007).

15. Silva, C.T. et al. Heritabilities, proportions of heritabilities explained by GWAS findings, and implications of cross-phenotype effects on PR interval. Hum Genet 134, 1211–9 (2015).

16. Duijvenboden, S. et al. Genomic and pleiotropic analyses of resting QT interval identifies novel loci and overlap with atrial electrical disorders. Hum Mol Genet (2021).

17. van der Harst, P. et al. 52 Genetic Loci Influencing Myocardial Mass. J Am Coll Cardiol 68, 1435–1448 (2016).

18. Yang, J., Lee, S.H., Goddard, M.E. & Visscher, P.M. GCTA: a tool for genome-wide complex trait analysis. Am J Hum Genet 88, 76–82 (2011).

19. Eriksson, A.L. et al. Genetic Determinants of Circulating Estrogen Levels and Evidence of a Causal Effect of Estradiol on Bone Density in Men. J Clin Endocrinol Metab 103, 991–1004 (2018).

20. Kemp, J.P. et al. Identification of 153 new loci associated with heel bone mineral density and functional involvement of GPC6 in osteoporosis. Nat Genet 49, 1468–1475 (2017).

21. Yap, C.X. et al. Dissection of genetic variation and evidence for pleiotropy in male pattern baldness. Nat Commun 9, 5407 (2018).

22. Jin, G. et al. Genome-wide association study identifies a new locus JMJD1C at 10q21 that may influence serum androgen levels in men. Hum Mol Genet 21, 5222–8 (2012).

23. Martinez-Garay, I. et al. A new gene family (FAM9) of low-copy repeats in Xp22.3 expressed exclusively in testis: implications for recombinations in this region. Genomics 80, 259–67 (2002).

24. Bulik-Sullivan, B.K. et al. LD Score regression distinguishes confounding from polygenicity in genome-wide association studies. Nat Genet 47, 291–5 (2015).

25. Bulik-Sullivan, B. et al. An atlas of genetic correlations across human diseases and traits. Nat Genet 47, 1236–41 (2015).

26. McLaren, W. et al. The Ensembl Variant Effect Predictor. Genome Biol 17, 122 (2016).

27. Wu, M.C. et al. Rare-variant association testing for sequencing data with the sequence kernel association test. Am J Hum Genet 89, 82–93 (2011).

28. Milano, A., Lodder, E.M. & Bezzina, C.R. TNNI3K in cardiovascular disease and prospects for therapy. J Mol Cell Cardiol 82, 167–73 (2015).

29. Lal, H., Ahmad, F., Parikh, S. & Force, T. Troponin I-interacting protein kinase: a novel cardiac-specific kinase, emerging as a molecular target for the treatment of cardiac disease. Circ J 78, 1514–9 (2014).

30. Nishio, Y. et al. D85N, a KCNE1 polymorphism, is a disease-causing gene variant in long QT syndrome. J Am Coll Cardiol 54, 812–9 (2009).

31. Wang, H. et al. Mutations in NEXN, a Z-disc gene, are associated with hypertrophic cardiomyopathy. Am J Hum Genet 87, 687–93 (2010).

32. Zhang, X.L. et al. Genetic Basis and Genotype-Phenotype Correlations in Han Chinese Patients with Idiopathic Dilated Cardiomyopathy. Sci Rep 10, 2226 (2020).

33. Consortium, G. Human genomics. The Genotype-Tissue Expression (GTEx) pilot analysis: multitissue gene regulation in humans. Science 348, 648–60 (2015).

34. Schmitt, A.D. et al. A Compendium of Chromatin Contact Maps Reveals Spatially Active Regions in the Human Genome. Cell Rep 17, 2042–2059 (2016).

35. Jung, I. et al. A compendium of promoter-centered long-range chromatin interactions in the human genome. Nat Genet 51, 1442–1449 (2019).

36. Hocker, J.D. et al. Cardiac cell type-specific gene regulatory programs and disease risk association. Sci Adv 7(2021).

37. Iotchkova, V. et al. GARFIELD classifies disease-relevant genomic features through integration of functional annotations with association signals. Nat Genet 51, 343–353 (2019).

38. Pers, T.H. et al. Biological interpretation of genome-wide association studies using predicted gene functions. Nat Commun 6, 5890 (2015).

39. Perestrelo, A.R. et al. Multiscale Analysis of Extracellular Matrix Remodeling in the Failing Heart. Circ Res 128, 24–38 (2021).

40. Finan, C. et al. The druggable genome and support for target identification and validation in drug development. Sci Transl Med 9(2017).

41. Lodola, F. et al. Adeno-associated virus-mediated CASQ2 delivery rescues phenotypic alterations in a patient-specific model of recessive catecholaminergic polymorphic ventricular tachycardia. Cell Death Dis 7, e2393 (2016).

42. Li, W. et al. PLK2 modulation of enriched TAp73 affects osteogenic differentiation and prognosis in human osteosarcoma. Cancer Med 9, 4371–4385 (2020).

43. Mitchell, R.N. et al. Effect of Sex and Underlying Disease on the Genetic Association of QT Interval and Sudden Cardiac Death. J Am Heart Assoc 8, e013751 (2019).

44. Ågesen, F.N. et al. Temporal trends and sex differences in sudden cardiac death in the Copenhagen City Heart Study. Heart 107, 1303–1309 (2021).

45. Barth, A.S. & Tomaselli, G.F. Cardiac metabolism and arrhythmias. Circ Arrhythm Electrophysiol 2, 327–35 (2009).

46. Huang, J.P., Huang, S.S., Deng, J.Y. & Hung, L.M. Impairment of insulin-stimulated Akt/GLUT4 signaling is associated with cardiac contractile dysfunction and aggravates I/R injury in STZ- diabetic rats. J Biomed Sci 16, 77 (2009).

47. Slot, J.W., Geuze, H.J., Gigengack, S., James, D.E. & Lienhard, G.E. Translocation of the glucose transporter GLUT4 in cardiac myocytes of the rat. Proc Natl Acad Sci U S A 88, 7815–9 (1991).

48. Maria, Z., Campolo, A.R., Scherlag, B.J., Ritchey, J.W. & Lacombe, V.A. Dysregulation of insulin-sensitive glucose transporters during insulin resistance-induced atrial fibrillation. Biochim Biophys Acta Mol Basis Dis 1864, 987–996 (2018).

49. Bellai-Dussault, K., Nguyen, T.T.M., Baratang, N.V., Jimenez-Cruz, D.A. & Campeau, P.M. Clinical variability in inherited glycosylphosphatidylinositol deficiency disorders. Clin Genet 95, 112–121 (2019).

50. Manea, E. A step closer in defining glycosylphosphatidylinositol anchored proteins role in health and glycosylation disorders. Mol Genet Metab Rep 16, 67–75 (2018).

51. Fedorov, V.V. et al. Effects of KATP channel openers diazoxide and pinacidil in coronary-perfused atria and ventricles from failing and non-failing human hearts. J Mol Cell Cardiol 51, 215–25 (2011).

52. Flagg, T.P. et al. Differential structure of atrial and ventricular KATP: atrial KATP channels require SUR1. Circ Res 103, 1458–65 (2008).

53. Gloyn, A.L., Siddiqui, J. & Ellard, S. Mutations in the genes encoding the pancreatic beta-cell KATP channel subunits Kir6.2 (KCNJ11) and SUR1 (ABCC8) in diabetes mellitus and hyperinsulinism. Hum Mutat 27, 220–31 (2006).

54. Abel, E.D. Insulin signaling in the heart. Am J Physiol Endocrinol Metab 321, E130–E145 (2021).

55. Polina, I. et al. Loss of insulin signaling may contribute to atrial fibrillation and atrial electrical remodeling in type 1 diabetes. Proc Natl Acad Sci U S A 117, 7990–8000 (2020).

56. Lu, Z. et al. Decreased L-type Ca2+ current in cardiac myocytes of type 1 diabetic Akita mice due to reduced phosphatidylinositol 3-kinase signaling. Diabetes 56, 2780–9 (2007).

57. Lu, Z. et al. Increased persistent sodium current due to decreased PI3K signaling contributes to QT prolongation in the diabetic heart. Diabetes 62, 4257–65 (2013).

58. Rautaharju, P.M. et al. Sex differences in the evolution of the electrocardiographic QT interval with age. Can J Cardiol 8, 690–5 (1992).

59. Masuda, K. et al. Testosterone-mediated upregulation of delayed rectifier potassium channel in cardiomyocytes causes abbreviation of QT intervals in rats. J Physiol Sci 68, 759–767 (2018).

60. Marsh, J.D. et al. Androgen receptors mediate hypertrophy in cardiac myocytes. Circulation 98, 256–61 (1998).

61. McGill, H.C., Anselmo, V.C., Buchanan, J.M. & Sheridan, P.J. The heart is a target organ for androgen. Science 207, 775–7 (1980).

62. Gray, A., Feldman, H.A., McKinlay, J.B. & Longcope, C. Age, disease, and changing sex hormone levels in middle-aged men: results of the Massachusetts Male Aging Study. J Clin Endocrinol Metab 73, 1016–25 (1991).

63. Mohamed, R., Forsey, P.R., Davies, M.K. & Neuberger, J.M. Effect of liver transplantation on QT interval prolongation and autonomic dysfunction in end-stage liver disease. Hepatology 23, 1128–34 (1996).

64. Bidoggia, H. et al. Sex differences on the electrocardiographic pattern of cardiac repolarization: possible role of testosterone. Am Heart J 140, 678–83 (2000).

65. Charbit, B. et al. Effects of testosterone on ventricular repolarization in hypogonadic men. Am J Cardiol 103, 887–90 (2009).

66. Schwartz, J.B. et al. Effects of testosterone on the Q-T interval in older men and older women with chronic heart failure. Int J Androl 34, e415–21 (2011).

67. Hönes, G.S. et al. Noncanonical thyroid hormone signaling mediates cardiometabolic effects in vivo. Proc Natl Acad Sci U S A 114, E11323–E11332 (2017).

68. Moskowitz, I.P. et al. A molecular pathway including Id2, Tbx5, and Nkx2-5 required for cardiac conduction system development. Cell 129, 1365–76 (2007).

69. Al Sayed, Z.R. et al. Human model of IRX5 mutations reveals key role for this transcription factor in ventricular conduction. Cardiovasc Res (2020).

70. Wu, L. et al. Bone morphogenetic protein 4 promotes the differentiation of Tbx18-positive epicardial progenitor cells to pacemaker-like cells. Exp Ther Med 17, 2648–2656 (2019).

71. Gewies, A. et al. Prdm6 is essential for cardiovascular development in vivo. PLoS One 8, e81833 (2013).

72. Miller, C.L. et al. Cyclic nucleotide phosphodiesterase 1A: a key regulator of cardiac fibroblast activation and extracellular matrix remodeling in the heart. Basic Res Cardiol 106, 1023–39 (2011).

73. Ten Tusscher, K.H. & Panfilov, A.V. Influence of diffuse fibrosis on wave propagation in human ventricular tissue. Europace 9 **Suppl 6**, vi38-45 (2007).

74. Nauffal, V. et al. Monogenic and Polygenic Contributions to QTc Prolongation in the Population. medRxiv, 2021.06.18.21258578 (2021).

75. Cho, M.S. et al. Clinical Implications of Ventricular Repolarization Parameters on Long-Term Risk of Atrial Fibrillation - Longitudinal Follow-up Data From a General Ambulatory Korean Population. Circ J 84, 1067–1074 (2020).

76. Zhang, N. et al. Prolonged corrected QT interval in predicting atrial fibrillation: A systematic review and meta-analysis. Pacing Clin Electrophysiol 41, 321–327 (2018).

77. Aeschbacher, S. et al. Relationship between QRS duration and incident atrial fibrillation. Int J Cardiol 266, 84–88 (2018).

78. Borggrefe, M. et al. Short QT syndrome. Genotype-phenotype correlations. J Electrocardiol 38, 75–80 (2005).

79. Nielsen, J.B. et al. J-shaped association between QTc interval duration and the risk of atrial fibrillation: results from the Copenhagen ECG study. J Am Coll Cardiol 61, 2557–64 (2013).

80. Feld, G.K. & Cha, Y. Electrophysiologic Effects of the New Class III Antiarrhythmic Drug Dofetilide in an Experimental Canine Model of Pacing-induced Atrial Fibrillation. J Cardiovasc Pharmacol Ther 2, 195–203 (1997).

81. Riera, A.R. et al. Relationship among amiodarone, new class III antiarrhythmics, miscellaneous agents and acquired long QT syndrome. Cardiol J 15, 209–19 (2008).

82. Psaty, B.M. et al. Cohorts for Heart and Aging Research in Genomic Epidemiology (CHARGE) Consortium: Design of prospective meta-analyses of genome-wide association studies from 5 cohorts. Circ Cardiovasc Genet 2, 73–80 (2009).

83. Auton, A. et al. A global reference for human genetic variation. Nature 526, 68–74 (2015).

84. McCarthy, S. et al. A reference panel of 64,976 haplotypes for genotype imputation. Nat Genet 48, 1279–83 (2016).

85. Loh, P.R. et al. Efficient Bayesian mixed-model analysis increases association power in large cohorts. Nat Genet 47, 284–90 (2015).

86. JR, O.C. MMAP User Guide. Available: http://edn.som.umaryland.edu/mmap/index.php. Accessed 7th December 2020.

87. Zhou, W. et al. Efficiently controlling for case-control imbalance and sample relatedness in large-scale genetic association studies. Nat Genet 50, 1335–1341 (2018).

88. Kang, H.M. et al. Variance component model to account for sample structure in genome-wide association studies. Nat Genet 42, 348–54 (2010).

89. Zhan, X., Hu, Y., Li, B., Abecasis, G.R. & Liu, D.J. RVTESTS: an efficient and comprehensive tool for rare variant association analysis using sequence data. Bioinformatics 32, 1423–6 (2016).

90. Winkler, T.W. et al. Quality control and conduct of genome-wide association meta-analyses. Nat Protoc 9, 1192–212 (2014).

91. Devlin, B. & Roeder, K. Genomic control for association studies. Biometrics 55, 997–1004 (1999).

92. Willer, C.J., Li, Y. & Abecasis, G.R. METAL: fast and efficient meta-analysis of genomewide association scans. Bioinformatics 26, 2190–1 (2010).

93. Purcell, S. et al. PLINK: a tool set for whole-genome association and population-based linkage analyses. Am J Hum Genet 81, 559–75 (2007).

94. Pruim, R.J. et al. LocusZoom: regional visualization of genome-wide association scan results. Bioinformatics 26, 2336–7 (2010).

95. Shim, H. et al. A multivariate genome-wide association analysis of 10 LDL subfractions, and their response to statin treatment, in 1868 Caucasians. PLoS One 10, e0120758 (2015).

96. Feng, S., Liu, D., Zhan, X., Wing, M.K. & Abecasis, G.R. RAREMETAL: fast and powerful meta-analysis for rare variants. Bioinformatics 30, 2828–9 (2014).

97. Kumar, P., Henikoff, S. & Ng, P.C. Predicting the effects of coding non-synonymous variants on protein function using the SIFT algorithm. Nat Protoc 4, 1073–81 (2009).

98. Adzhubei, I.A. et al. A method and server for predicting damaging missense mutations. Nat Methods 7, 248–9 (2010).

99. Kircher, M. et al. A general framework for estimating the relative pathogenicity of human genetic variants. Nat Genet 46, 310–5 (2014).

100. Boyle, A.P. et al. Annotation of functional variation in personal genomes using RegulomeDB. Genome Res 22, 1790–7 (2012).

101. Project, T.G.-T.E.G. The Genotype-Tissue Expression (GTEx) Project was supported by the Common Fund of the Office of the Director of the National Institutes of Health, and by NCI, NHGRI, NHLBI, NIDA, NIMH, and NINDS.

102. Battle, A. et al. Genetic effects on gene expression across human tissues. Nature 550, 204–213 (2017).

103. Consortium, G. The GTEx Consortium atlas of genetic regulatory effects across human tissues. Science 369, 1318–1330 (2020).

104. Giambartolomei, C. et al. Bayesian test for colocalisation between pairs of genetic association studies using summary statistics. PLoS Genet 10, e1004383 (2014).

105. Watanabe, K., Taskesen, E., van Bochoven, A. & Posthuma, D. Functional mapping and annotation of genetic associations with FUMA. Nat Commun 8, 1826 (2017).

106. Soskic, B. et al. Chromatin activity at GWAS loci identifies T cell states driving complex immune diseases. Nat Genet 51, 1486–1493 (2019).

107. Shannon, P. et al. Cytoscape: a software environment for integrated models of biomolecular interaction networks. Genome Res 13, 2498–504 (2003).

108. Raudvere, U. et al. g:Profiler: a web server for functional enrichment analysis and conversions of gene lists (2019 update). Nucleic Acids Res 47, W191–W198 (2019).

109. Choi, S.W. & O’Reilly, P.F. PRSice-2: Polygenic Risk Score software for biobank-scale data. Gigascience 8(2019).

110. The Atherosclerosis Risk in Communities (ARIC) Study: design and objectives. The ARIC investigators. Am J Epidemiol 129, 687–702 (1989).

111. Wright, J.D. et al. The ARIC (Atherosclerosis Risk In Communities) Study: JACC Focus Seminar 3/8. J Am Coll Cardiol 77, 2939–2959 (2021).

112. Sotoodehnia, N. et al. Beta2-adrenergic receptor genetic variants and risk of sudden cardiac death. Circulation 113, 1842–8 (2006).

113. Haukilahti, M.A.E. et al. Sudden Cardiac Death in Women. Circulation 139, 1012–1021 (2019).

114. Sabatti, C. et al. Genome-wide association analysis of metabolic traits in a birth cohort from a founder population. Nat Genet 41, 35–46 (2009).

115. Balduzzi, S., Rücker, G. & Schwarzer, G. How to perform a meta-analysis with R: a practical tutorial. Evid Based Ment Health 22, 153–160 (2019).

